# GXwasR: A Toolkit for Investigating Sex-Differentiated Genetic Effects on Complex Traits

**DOI:** 10.1101/2025.06.10.25329327

**Authors:** Banabithi Bose, Freida Blostein, Jeewoo Kim, Jessica Winters, Ky’Era V. Actkins, David Mayer, Harrsha Congivaram, Maria Niarchou, Digna Velez Edwards, Lea K. Davis, Barbara E. Stranger

**Affiliations:** Department of Biomedical Informatics, University of Colorado School of Medicine, Aurora, CO; Vanderbilt Genetics Institute, Vanderbilt University Medical Center, Nashville, TN; Division of Genetic Medicine, Department of Medicine, Vanderbilt University Medical Center, Nashville, TN; Division of Quantitative and Clinical Sciences, Vanderbilt University, Nashville, TN; Medical Scientist Training Program, Vanderbilt University, Nashville, TN; Center for Genetic Medicine, Feinberg School of Medicine, Northwestern University, Chicago, IL; Department of Obstetrics and Gynecology, Department of Medicine, Vanderbilt University Medical Center, Nashville, TN; Department of Biomedical Informatics, Institute for Medicine and Public Health, Vanderbilt Epidemiology Center, Nashville, TN; Mount Sinai Million Health Discoveries Program, Charles Bronfman Institute for Personalized Medicine, New York, NY; Department of AI and Human Health, Icahn School of Medicine at Mount Sinai, New York, NY; Division of Data-Driven and Digital Medicine, Department of Medicine, Icahn School of Medicine at Mount Sinai, New York, NY; New York Genome Center, New York, NY

## Abstract

Many complex traits in humans exhibit sex differences. Recent advances in understanding context-specific genetic effects underscore the imperative for sex-aware genetic analyses to assess the role of sex in the genetic basis of human phenotypes. Despite increasing recognition of the importance of sex as a biological variable, current computational tools for genome-wide association studies (GWAS) lack functionality for performing nuanced sex-aware quality control and analysis. In response, we present GXwasR, a comprehensive R package that implements statistical genetic models for performing both GWAS and X-Chromosome-Wide Association Studies (XWAS). Designed for both sex-combined and sex-stratified analyses, GXwasR can account for unique features of chromosome X, including X-chromosome inactivation. Beyond enabling association analysis and meta-analysis, the tool has functions to a) test for sex differences in SNP-level genetic effects and heritability, b) assess genetic correlation, c) compute sex-aware polygenic risk scores (PRS), and d) support enhanced quality control protocols expressly designed for these purposes. GXwasR thus serves as a computational resource to enable investigations into the role of sex in the genetic basis of complex human phenotypes.

## Introduction

In biomedical research, sex emerged as an important biological variable^1,2^, profoundly shaping human health and disease^3,4^. Advances in genetics and biomedicine are revealing how genetic and physiological sex differences interact to influence complex traits such as autoimmune and cardiometabolic diseases, cancer, and response to treatment. This underscores the importance of incorporating sex as a factor in biomedical research, aligning with precision medicine’s focus on individualized care. As a result, global research organizations now mandate the consideration of sex as a biological variable throughout all stages of biomedical research^4^.

Yet despite these advances, our understanding of how sex affects the genetic architecture of complex traits remains incomplete. Although sex differences can substantially influence disease risk and trait variation^5–7^, most genome-wide association studies (GWAS) do not explicitly test for sex-specific effects—often omitting the sex chromosomes entirely^8,9^. This omission is especially concerning given the unique features of the X and Y chromosomes^10–14^, which likely harbor underexplored genetic variation. Addressing this knowledge gap calls for robust, sex-aware analytical tools capable of integrating both autosomal data and the distinct biology of the sex chromosomes. Such a tool must correctly account for the intricacies of both autosomal and Chromosome X (hereafter, ChrX) genetic data, accommodating unique aspects such as XCI patterns^9,15,16^, and be able to identify sex-differentiated genetic effects.

We present GXwasR, an R-based tool enabling sex-aware genetic analysis across autosomes and chromosome X. Building on our recent guidelines for analyzing sex differences in complex traits^17^, GXwasR provides thirty-three functions organized into six main categories: (A) pre-imputation quality control (QC), (B) post-imputation QC, (C) sex-combined and sex-stratified GWAS and XWAS, (D) tests for sex-differentiated genetic effects, (E) additional analyses (heritability estimation, genetic correlation, gene-based testing, polygenic risk scores (PRS) computation, and meta-analysis), and (F) utility functions.

Beyond QC and association testing, GXwasR supports additional analyses, including SNP × sex interaction tests and sex-stratified association followed by Z-score or t-statistic comparisons. It also addresses unique aspects of ChrX, such as random, skewed, and escaped X-chromosome inactivation (XCI) and pseudoautosomal regions (PAR)—and introduces sex-specific checks for minor allele frequency (MAF) and Hardy–Weinberg equilibrium (HWE).

GXwasR integrates seamlessly with established tools, PLINK^18^,GCTA^19^, REGENIE^20^ and SAIGE^21^. It leverages R’s statistical and graphical capabilities to streamline sex-aware genetic research, offering visualization options, flexible workflows, and extensive documentation. Together these features make GXwasR a versatile resource for investigating sex differences in the genetics of complex traits.

### Sex-aware QC of genotype data with GXwasR

GXwasR integrates a comprehensive QC pipeline for analyzing both autosomal and ChrX genotypes, covering pre- and post-imputation steps (Figure 1). It can accommodate pre- and post-imputation genotype data as well as variants derived from whole-genome sequencing. Importantly, this tool is not limited to sex-aware analysis; it also accommodates combined-sex QC and analysis.

**Figure 1.**
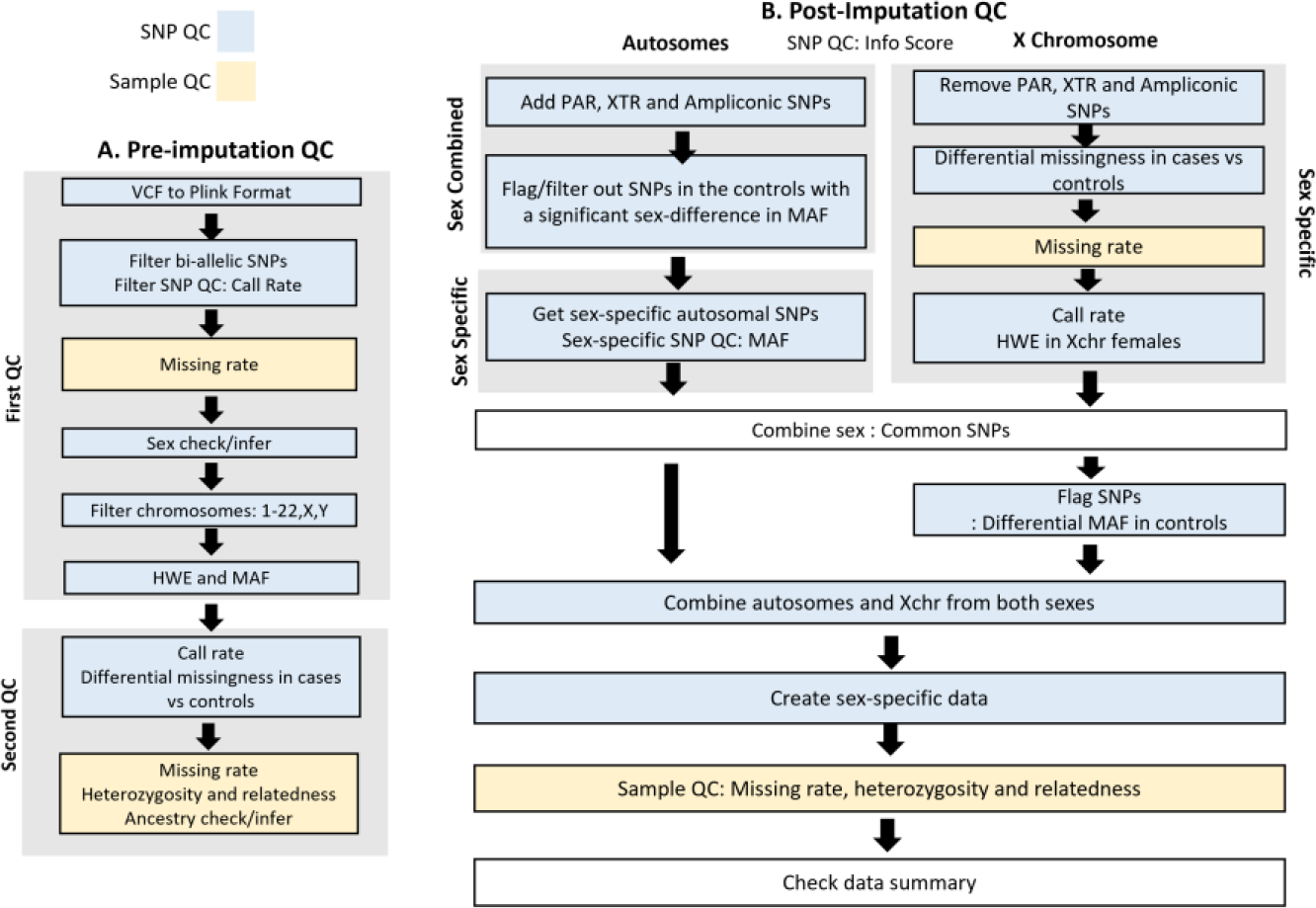
Pipelines for sex-aware QC of A) pre-imputed and B) post-imputed genotypes using GXwasR functions.

**Figure 3:**
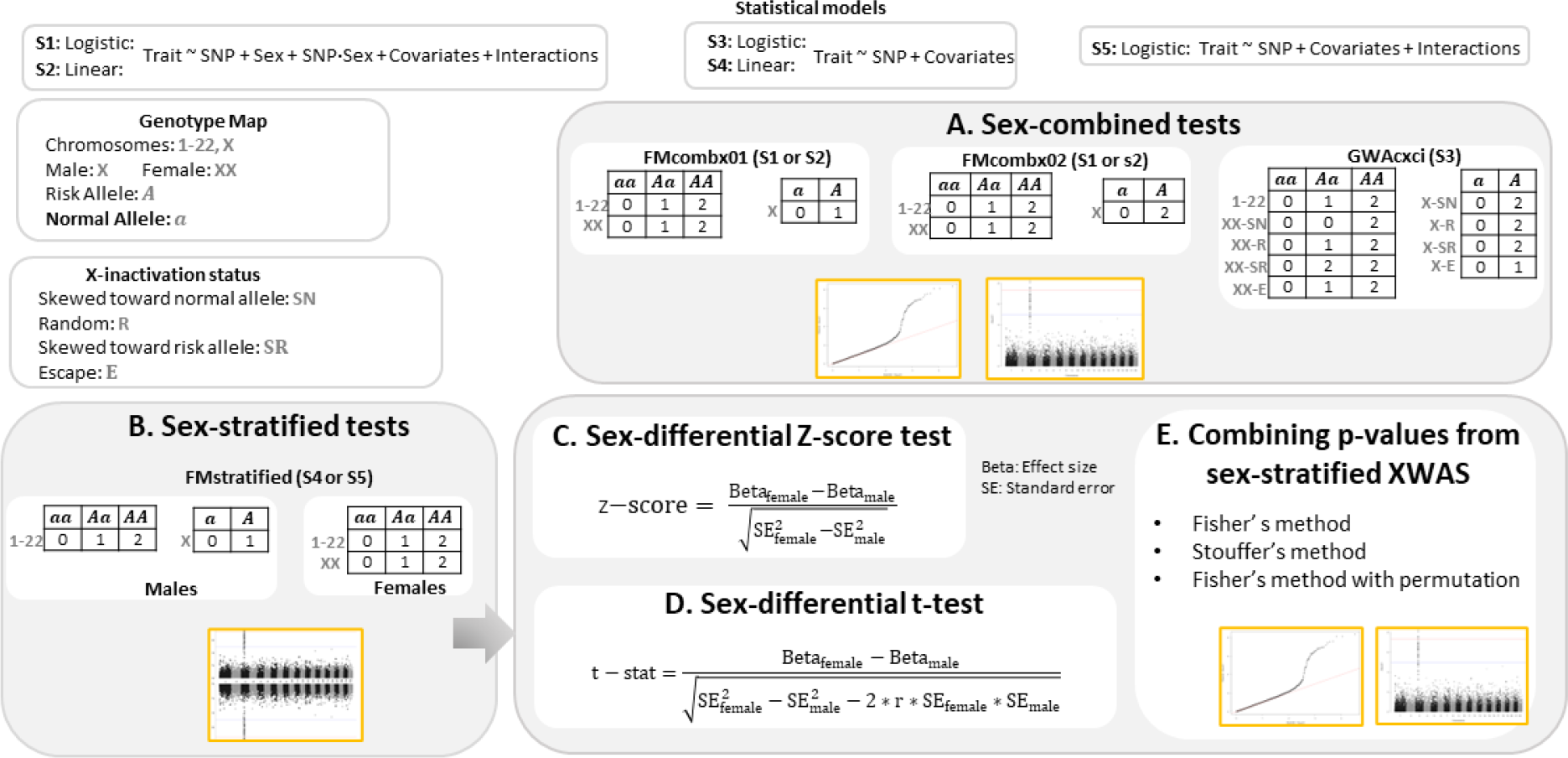
Overview of sex-aware GWAS and XWAS methods implemented in GXwasR: A) sex-combined tests using different ways of handling ChrX; B) sex-stratified tests; C) sex-differential Z-score test comparing effect sizes using standard errors; D) sex-differential t-test accounting for correlation between estimates; and E) methods to combine p-values from sex-stratified analyses using Fisher’s or Stouffer’s approaches.

**Figure 4:**
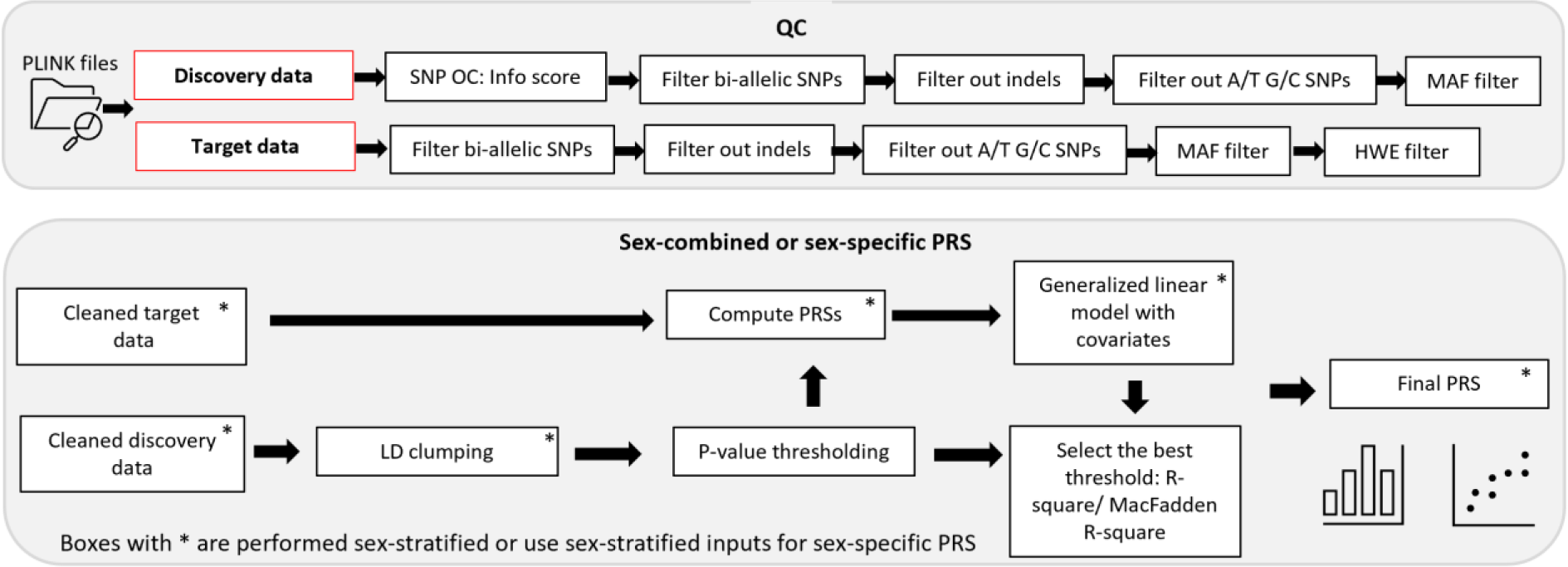
Overview of the PRS workflow, including: QC steps for discovery and target datasets (e.g., SNP info score filtering, removal of indels and ambiguous SNPs); sex-combined or sex-specific PRS construction involving LD clumping, p-value thresholding, PRS computation, and model fitting; and selection of the best-fit threshold using R-square metrics, with sex-stratified analyses indicated by asterisks.

The pre-imputation QC pipeline is supported by the following features:

#### Ancestry and chromosomal sex verification

The tool verifies user-provided ancestry labels or determines them using genetic data from reference populations. It also confirms or imputes sex assignments using genetic data from sex chromosomes.

#### SNP level QC

The tool performs QC of autosomal and ChrX variants. This involves filtering based on quality metrics, including genotyping efficiency, call rate, HWE, MAF and high-LD regions.

#### Sample level QC

The tool can identify and remove samples with a high rate of missing data and or heterozygosity.

#### Flexible thresholds

While default thresholds are provided based on prior literature^22,23^, but users can adjust them as needed.

#### Visual displays

The tool provides visualization, including plots and tables.

Detailed GXwasR pipelines and tutorials for pre-imputation QC, ancestry estimation and post-imputation QC are available in the **Supplementary File**.

The post-imputation QC pipeline is supported by the following features (Figure 1):

#### Filtering SNPs with low imputation accuracy

The tool can remove SNPs based on *r*^2^ values and average certainty of best-guess genotypes.

*The tool can either remove or flag specific regions of ChrX that may confound the analysis, or integrate these regions with autosomes if appropriate*

The tool can exclude or flag specific regions of ChrX that could potentially confound the analysis, (i.e., Pseudoautosomal Regions^24^, X-Transposed Regions^25^, and Ampliconic regions^26^).

#### Sex differences in allele frequencies within controls

GXwasR can test for sex-differentiated allele frequencies among controls which can indicate technical issues or biases within the cohort.

#### MAF thresholds for autosomal SNPs

The tool can filter autosomal SNPs based on MAF, which can be adjusted separately for males and females.

#### Quality control for ChrX

The tool implements sex-stratified QC for ChrX variants while addressing the unique characteristics of ChrX.

#### Post-imputation Sample QC

The tool can remove samples based on missing genotype rate, heterozygosity, and identity-by-descent (IBD; pi-hat)^17^ separately for each sex.

### Sex-aware genetic association analysis

GXwasR performs sex-aware genetic association studies while accounting for unique aspects of sex chromosomes and autosomes. It includes the *GXwas()* function for full genome-wide analysis (GWAS + XWAS), with special modeling of ChrX variants. Unlike autosomes where genotypes are coded as 0, 1, and 2, ChrX variants can be subject to one of several XCI patterns. For XWAS, GXwasR codes female ChrX genotypes as 0, 1, and 2, while male genotypes are coded to reflect dosage compensation nuances, supporting several different XCI patterns (Figure 2).

The models supported by *GXwas()* for running GWAS including XWAS are

#### FMcombx01

Sex-combined analysis of autosomes and sex chromosomes, where female ChrX genotypes are coded as autosomal variants (i.e., 0, 1, and 2), whereas male ChrX genotypes are coded as 0 and 1 to reflect the XCI-Random pattern^27^.

#### FMcombx02

Sex-combined analysis of autosomes and sex chromosomes, where female ChrX genotypes are coded as autosomal variants, whereas male ChrX genotypes are coded as 0 and 2, to reflect the XCI-Escape pattern^28^.

Both models can accommodate covariates and can perform interaction tests, such as genotype-by-sex (i.e, SNP × Sex).

#### Fmstratified

Sex-stratified GWAS and XWAS with covariates.

#### Fmcomb

Combining the p-values of the *FMstratified* XWAS tests to obtain a final sex-combined XWAS p-values for each ChrX SNP using Fisher method^29^, Fisher method with permutation^30^ and Stouffer method^31^ to allow the ChrX SNP tested to have different, even an opposite, effect on disease risk in males and females. This functionality is automated into the *FMstratified* model of *GXwas()* function as well.

#### GWAcxci

This model represents a female-male combined analysis for autosomes, akin to *FMcombx01* or *FMcombx02*. However, for ChrX, it incorporates a continuous XCI pattern^32^, particularly suitable for binary traits.

The *GXwas()* function automatically generates Manhattan and quantile-quantile (QQ) plots to visualize GWAS and XWAS results.

### Test for sex-differentiated genetic architecture

GXwasR can test for sex-differentiated genetic effects using sex-stratified GWAS summary statistics as input. For each SNP, the tool utilizes a t-test and Z-score to test for a sex difference in effect size (Figure 2). For the t-test, the *SexDiff()* function is applied to summary statistics derived from sex-stratified tests and for the Z-score test, the *SexDiffZscore()* calculates the difference in any kind of measured entities such as effect size, SNP heritability, genetic correlation, etc., between sexes using a Z-score and associated p-value. This function also outputs the QQ plot of the Z-score distribution p-values. It is important to note that a study design that performs a sex-stratified analyses followed by a difference test equates to a SNP × Sex interaction test under certain conditions, specifically when there’s no interaction between covariates and sex, and when trait variances are similar across sexes^17^. This approach, while generally conservative, offers robustness against violations of these assumptions^17^, such as covariate interactions or sex differences in phenotypic variance.

Another function, *DiffZeroOne()*, evaluates a Z-score to assess deviations from one and zero, testing the null hypothesis that a statistic (e.g., genetic correlation between males and females for a given trait) is less than one using a one-tailed test against a normal distribution.

To apply these tests to a set of independent SNPs, GXwasR performs LD clumping using the *LDclump()* function

### Polygenic risk scores

A polygenic risk score (PRS) is a quantitative estimate of an individual’s cumulative genetic predisposition to a specific trait. GXwasR provides several functions for working with PRSs in the context of sex differences:

The *ComputePRS()* function generates polygenic risk scores, which are derived by summing the effects of multiple variants across the genome, each weighted by its effect size derived from GWAS. The function includes customizable Clumping + Thresholding (C + T) procedure to ensure that only independent trait-associated variants are used for PRS estimation.

The function conducts an implicit model fitting process, wherein a number of p-value thresholds are tested to maximize the predictive power of the resulting polygenic scores. The function evaluates model performance using R^2^ for quantitative traits and McFadden R^2^ for binary traits^33^. Multiple plots are generated, including visualizations of the PRS model fit as a function of p-value thresholds, the distributions of computed PRS in males and females, and among cases and controls. For binary traits, ROC curves are produced (see Supplementary File).

The *ComputePRS()* function, along with the *SexRegress()* function enable assessment of a genome-wide impact of sex differences on the genetic architecture of a trait which can manifest as sex differences in predictive performance of the PRS. For example, a PRS derived from one sex may predict trait status less accurately in the other sex^17,34,35^. In this context, *SexRegress()* facilitates the investigation of associations between a phenotype in the target sex using the PRS derived from effect sizes estimated in the other (discovery) sex. Differences may suggest a sex difference in the genetic burden of the trait, however, careful interpretation is warranted, as differences in sample size between sexes can impact prediction accuracy.

### Heritability and genetic correlation

GXwasR performs two types of heritability estimation, (i) GREML: Genetic relatedness matrix (GRM) restricted maximum likelihood as implemented in GCTA^19^ and (ii) LDSC: LD score regression^36,37^ using the *EstimateHerit()* function.

*For the GREML model, EstimateHerit()* fits the effects of all the SNPs as random effects in a mixed linear model. The function then estimates the variance explained by the SNPs using RELM (i.e., Restricted Maximum Likelihood) relying on the GRM. For ChrX, it adjusts for dosage compensation by modifying the GRM.

For the LDSC model, *EstimateHerit()* uses GWAS summary statistics and recomputed LD scores^37^. *EstimateHerit()* also matches genetic variants across datasets by flipping and strand reversing and provides the option to use LD-clumped SNPs in the computation of LD scores.

For both models, *EstimateHerit()* provides the trait heritability estimate in both observed and liability scales, and can partition heritability by MAF, chromosome, etc. It plots the estimated heritability vs. chromosome length, number of genes, and proportion of the SNPs per chromosome.

The functions *GeneticCorrBT()* and *SumstatGenCorr()* estimate genetic correlation between two quantitative traits, two binary traits, and between a quantitative trait and a binary trait. *GeneticCorrBT()* performs a bivariate GREML^19,38^ analysis utilizing genotype data and provides the option to compute genetic correlation using user defined MAF thresholds. *SumstatGenCorr()* applies High Definition Likelihood (HDL)^39^ method to two sets of GWAS summary statistics, e.g., sex-stratified summary statistics. Both functions provide the option to compute the genetic correlation separately for each chromosome.

### Meta-analysis

The *MetaGwas()* function combines summary statistics from separate GWASes. It utilizes an inverse variance-based approach to enable fixed-effect and random-effect models, and quantifies effect size heterogeneity using Cochran’s Q^40^ and the I^2^ statistics^41^. This function also calculates weighted Z-score-based p-values^42^. The function can also apply correction for genomic inflation^43^.

The *MetaGwas()* function must be applied to independent trait-associated variants or those in linkage equilibrium. The *ClumpLD()* function and the distinct loci provided as the *SNPfile* input can be used for this purpose. The *MetaGwas()* function also generates Manhattan, QQ, and forest plots.

### Gene-based tests

The *TestXGene()* function enables gene-based association testing, utilizing GWAS/XWAS summary statistics and SNP-SNP correlation matrices using either user-provided genotype data or 1000 Genomes Phase 3 reference data^44^. While *TestXGene()* is by default suited for X-linked gene-based tests, its design also allows for its application across the autosomes. The function provides flexibility to use pre-defined or custom genomic gene coordinates.

Central to *TestXGene()* is its capability to compute gene-based SNP-SNP correlation matrices. The function is able to perform a variety of gene-based tests, including the burden test (BT^45^), sequence kernel association test (SKAT^46^), SKAT-O^47^ (a combination of BT and SKAT), sum of *χ*^2^-statistics, aggregated Cauchy association test (ACAT^48^), principal component approach (PCA^49^), functional multiple linear regression model (FLM^50^), Bonferroni correction test (simpleM^51^), and minimum P-value (minp^51^). These tests are executed by leveraging the functionality of PLINK1.9 and sumFREGAT^51^ tools.

Tutorials for the functions above as well as more detailed descriptions of the statistical models are provided in Supplementary File.

### Application to human height

We illustrate the use of GXwasR to assess sex differences in the genetic basis of height using data from the Vanderbilt University Medical Center’s (VUMC) genetic biobank (BioVU) and the linked de-identified mirror of the VUMC electronic health record, known as the Synthetic Derivative (SD). Briefly, we used genotype data from 50,294 individuals (males = 21,881; females = 28,421) whose genetic data clustered with European reference populations and who had clinical data on height. After performing sex-aware genomic quality control using GXwasR, 3,359,461 variants were available for running sex-stratified GWAS using *GXwas()* function (see **Supplemental File**).

We identified 1,549 SNPs associated with height across both males and females (p < 5×10^−8^) from 71 independent loci (**Supplemental File**) using *ClumpLD()* function (**Supplemental File**). These included five loci identified only in females on chromosomes 9 (rs4385527; rs2297086) and 15 (rs16869; rs4843154; rs4414460), and one locus identified only in males, on chromosome 2 (rs12615742). Fifteen of these loci were novel (Table 2); for the other 56 loci, the lead or one of the clumped variants was previously reported to be associated with height in the NHGRI-EBI GWAS Catalog^52^. Loci associated in only one sex were more likely to be novel (*p-value* from Fisher’s exact test = 0.006). One novel locus on ChrX was identified near pseudogene RP1-164L12.1/CORO1C pseudogene 1.

**Table 1:** An overview of GXwasR’s functions ranging from quality control to statistical analysis. Note that these functions can be used across multiple categories. For example, *QCsnp* and *QCsample* can be used in both pre- and post-imputation. Detailed descriptions of these functions and example code are provided (see Supplementary File).

| **A. Pre-imputation Quality Control** |  |
| --- | --- |
| **Function Name** | **Functionality** |
| QCsnp | Performs quality control for SNPs. |
| QCsample | Performs quality control for samples. |
| SexCheck | Compares user-defined sex assignments with predictions based on X chromosome inbreeding coefficients or imputes sex. |
| AncestryCheck | Evaluates samples' ancestry and flags outliers. |
| **B. Post-imputation Quality Control** |  |
| **Function Name** | **Functionality** |
| FilterRegion | Removes user-defined chromosomal regions. |
| MAFdiffSexControl | Tests for significant sex differences in minor allele frequency (MAF) in controls |
| Xhwe | Removes X-chromosome variants violating Hardy-Weinberg Equilibrium in females. |
| **C. Sex-combined and sex-stratified GWAS with XWAS** |  |
| **Function Name** | **Functionality** |
| GXwas | Runs additive GWAS models for autosomes, and XWAS models for binary and quantitative traits. , and can incorporate covariates and their interactions. |
| PvalComb | Combines p-values from male and female-specific XWAS for each SNP. |
| **D. Test of sex-differentiated genetic effects** |  |
| **Function Name** | **Functionality** |
| DiffZeroOne | Assesses Z-scores for deviation from one and zero for metrics such as genetic correlation and heritability. |
| SexDiff | Evaluates sex differences in the genetic effect size for each SNP using a t-test. |
| SexDiffZscore | Evaluates sex differences in the genetic effect size for each SNP using a Z-score. |
| GXwas | Runs SNP x Sex interaction model. |

E. Additional analyses
| Function Name | Functionality |
| --- | --- |
| ComputeCorrB | Computes genetic correlation between two traits (or single trait in two subgroups, e.g. males and females) using germline genotypes. |
| SumstatGenCorr | Computes genetic correlation between two traits (or single trait in two subgroups, e.g. males and females) using GWAS summary statistics. |
| ComputePRS | Computes PRS using clumping and thresholding method. |
| SexRegress | Computes the association of a phenotype in the target sex using a PRS derived from the other (discovery) sex. |
| EstimateHerit | Computes SNP heritability using either genotype data or GWAS summary statistics. |
| MetaGXwas | Performs fixed-effect and random-effect meta-analysis across sets of GWAS summary statistics. |
| TestXGene | Performs gene-based association tests using GWAS/XWAS summary statistics. |

F. Utility Functions
| Function Name | Functionality |
| --- | --- |
| ClumpLD | Performs linkage disequilibrium (LD) clumping of SNPs. |
| LDPrune | Performs LD pruning of SNP data. |
| ComputeGeneticPC | Computes principal components from a genetic relationship matrix adjusting for high LD regions. |
| DummyCovar | Recodes a categorical covariate into binary dummy variables. |
| FilterAllele | Removes multi-allelic variants from genotype data. Note: GXwasR cannot handle analysis of multi-allelic variants. |
| filterPlinkSample | Prepares PLINK binary files with desired samples based on user-defined criteria. |
| FilterSNP | Removes specific SNPs based on user-defined criteria. |
| GetMFPlink | Prepares separate male and female PLINK binary files from combined-sex files. |
| MergeRegion | Combines two genotype datasets based on common or all SNPs. |
| PlinkSummary | Provides a summary of samples, variants and phenotypes in a PLINK formatted genotype dataset. |
| plinkVCF | Converts VCF files to PLINK binary formats and vice versa, including creation of dummy FAM files. |
| executePlinkMAF | Execute PLINK to Calculate Minor Allele Frequencies (MAF) |

**Table 2:**
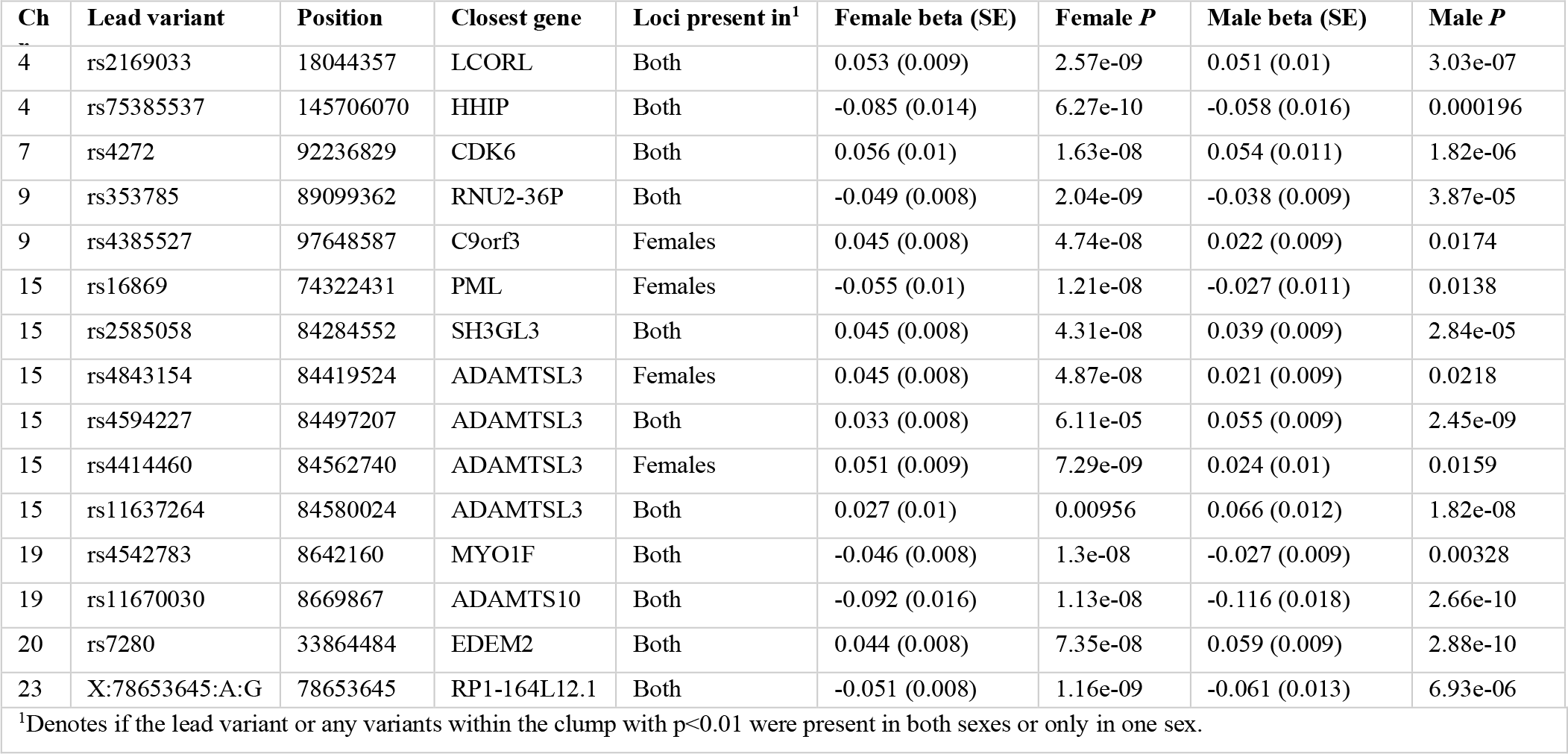
Novel significant lead variants (p < 5×10^−8^) from genome-wide association testing with height in BioVU data.

| Ch | Lead variant | Position | Closest gene | Loci present in<sup>1</sup> | Female beta (SE) | Female *P* | Male beta (SE) | Male *P* |
| --- | --- | --- | --- | --- | --- | --- | --- | --- |
| 4 | rs2169033 | 18044357 | LCORL | Both | 0.053 (0.009) | 2.57e-09 | 0.051 (0.01) | 3.03e-07 |
| 4 | rs75385537 | 145706070 | HHIP | Both | -0.085 (0.014) | 6.27e-10 | -0.058 (0.016) | 0.000196 |
| 7 | rs4272 | 92236829 | CDK6 | Both | 0.056 (0.01) | 1.63e-08 | 0.054 (0.011) | 1.82e-06 |
| 9 | rs353785 | 89099362 | RNU2-36P | Both | -0.049 (0.008) | 2.04e-09 | -0.038 (0.009) | 3.87e-05 |
| 9 | rs4385527 | 97648587 | C9orf3 | Females | 0.045 (0.008) | 4.74e-08 | 0.022 (0.009) | 0.0174 |
| 15 | rs16869 | 74322431 | PML | Females | -0.055 (0.01) | 1.21e-08 | -0.027 (0.011) | 0.0138 |
| 15 | rs2585058 | 84284552 | SH3GL3 | Both | 0.045 (0.008) | 4.31e-08 | 0.039 (0.009) | 2.84e-05 |
| 15 | rs4843154 | 84419524 | ADAMTSL3 | Females | 0.045 (0.008) | 4.87e-08 | 0.021 (0.009) | 0.0218 |
| 15 | rs4594227 | 84497207 | ADAMTSL3 | Both | 0.033 (0.008) | 6.11e-05 | 0.055 (0.009) | 2.45e-09 |
| 15 | rs4414460 | 84562740 | ADAMTSL3 | Females | 0.051 (0.009) | 7.29e-09 | 0.024 (0.01) | 0.0159 |
| 15 | rs11637264 | 84580024 | ADAMTSL3 | Both | 0.027 (0.01) | 0.00956 | 0.066 (0.012) | 1.82e-08 |
| 19 | rs4542783 | 8642160 | MYO1F | Both | -0.046 (0.008) | 1.3e-08 | -0.027 (0.009) | 0.00328 |
| 19 | rs11670030 | 8669867 | ADAMTS10 | Both | -0.092 (0.016) | 1.13e-08 | -0.116 (0.018) | 2.66e-10 |
| 20 | rs7280 | 33864484 | EDEM2 | Both | 0.044 (0.008) | 7.35e-08 | 0.059 (0.009) | 2.88e-10 |
| 23 | X:78653645:A:G | 78653645 | RP1-164L12.1 | Both | -0.051 (0.008) | 1.16e-09 | -0.061 (0.013) | 6.93e-06 |

<sup>1</sup>Denotes if the lead variant or any variants within the clump with $p < 0.01$ were present in both sexes or only in one sex.

While this locus is novel, other variants near RP1-164L12.1/CORO1C pseudogene 1 have previously been reported to be associated with height^53,54^.

The t-test for sex differences was performed via *SexDiff()* on variants (see Supplementary File) with a nominal association (p < 0.05) in at least one sex. Thirty-four variants from four independent loci were identified as having different effects by sex (Benjamini-Hochberg adjusted p < 0.05). None of these variants were reported in the NHGRI-EBI GWAS Catalog as being associated with height. However, other variants mapping to three of the four annotated genes (Table 3) have previously been reported to be associated with height^55–57^.

**Table 3:** Independent loci with sex-differentiated genetic effects on height (p < 0.05) after running the SexDiff() function with BioVU data.

| CHR | SNP | BP | Closest gene | Female beta (SE) | Female *P* | Male beta (SE) | Male *P* |
| --- | --- | --- | --- | --- | --- | --- | --- |
| 9 | rs324537 | 9047726 | PTPRD* | 0.06 (0.017) | 0.00033 | -0.07 (0.019) | 0.000578 |
| 11 | rs142535863 | 1.02E+08 | TRPC6* | -0.06 (0.01) | 0.000199 | 0.05 (0.017) | 0.002524 |
| 14 | rs55638307 | 77429613 | RP11-7F17.7 | 0.06 (0.018) | 0.001086 | -0.07 (0.020) | 0.000875 |
| 1 | rs4908192 | 1.02E+08 | OLFM3* | 0.03 (0.008) | 0.000294 | -0.03 (0.009) | 0.003164 |

\*Denotes variants mapped to this gene in the GWAS catalog have previously been reported as associated with height

Heritability was estimated using the *EstimateHerit()* function, which implements both GREML and LDSC models. For GREML, the function computed the GRM from BioVU genotype data across chromosomes 1–23 and estimated chromosome-specific (Figure 6–7) as well as genome-wide heritability in each sex (see Supplemental File).

**Figure 6.**
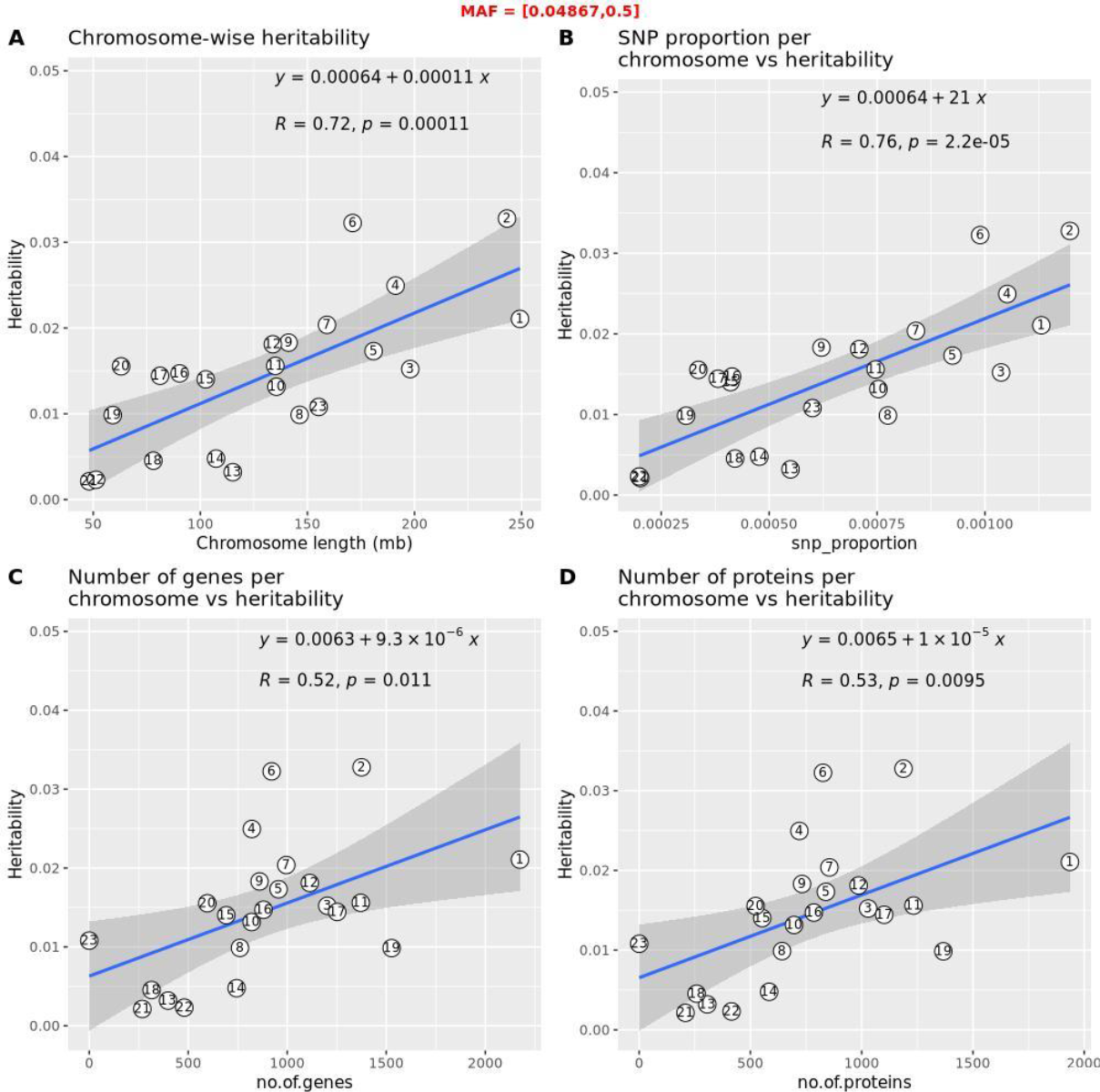
Male heritability per chromosome for height as estimated using *EstimateHerit()* function with BioVU data and GREML models.

**Figure 7.**
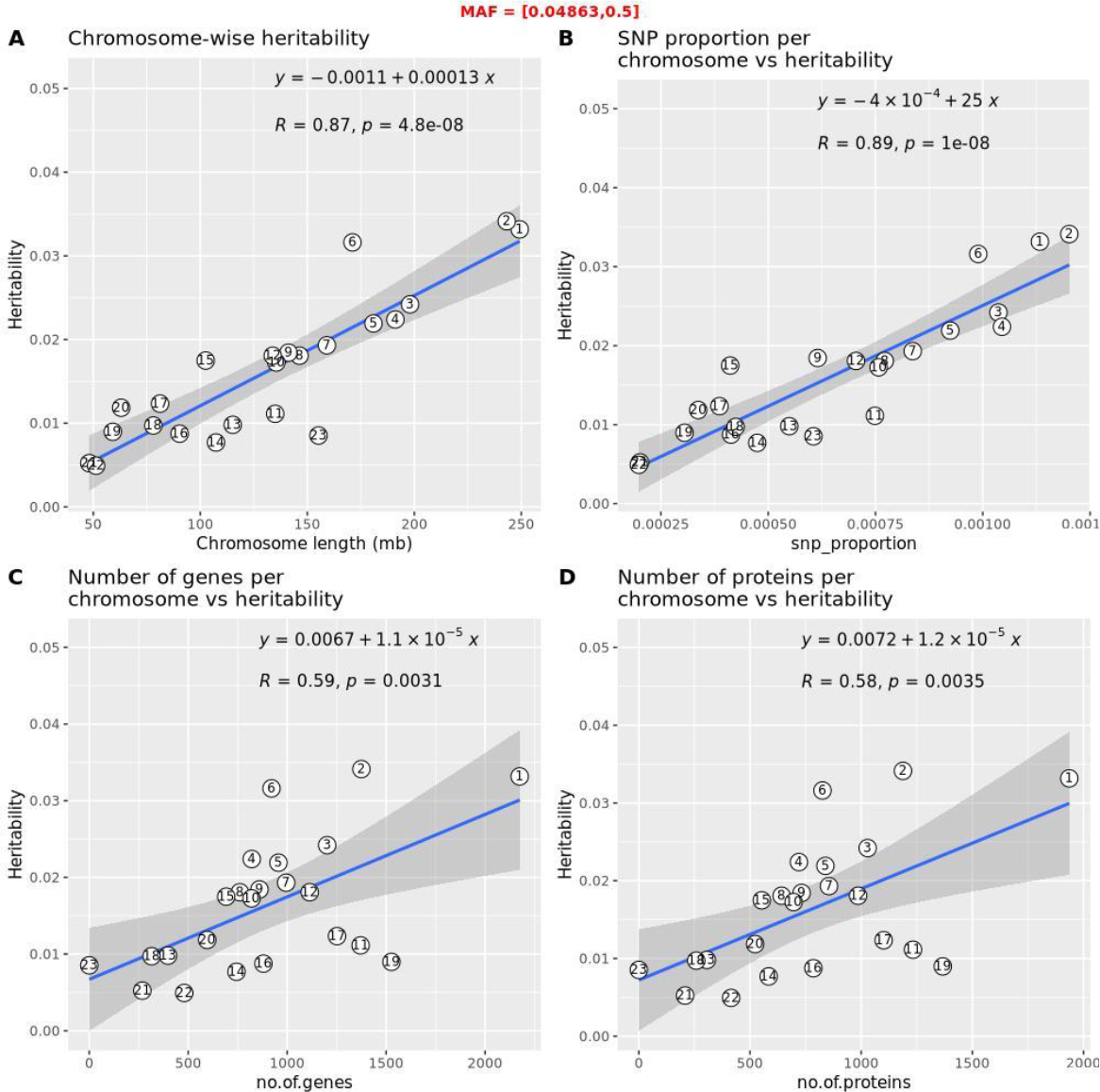
Female heritability per chromosome for height as estimated using *EstimateHerit()* function with BioVU data and GREML models.

For LDSC, the function applied the LDSC model using sex-stratified GWAS and XWAS summary statistics, together with precomputed LD scores derived from UK Biobank genotype data^36^, to compute genome-wide heritability (see Supplemental File). Genome-wide height heritability estimates were similar between the two sexes and recapitulated previously reported heritability estimates (Table 4). Male height heritability ranged between 0.21-0.34, and female height heritability between 0.24-0.36. The GREML model that utilized GRMs estimated a higher heritability compared to the LDSC model that used GWAS summary statistics and precomputed scores.

**Table 4:**
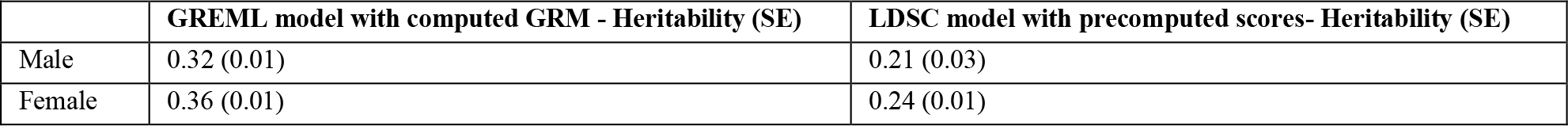
Sex-stratified heritability estimates with associated standard error (SE) for height as generated via the *EstimateHerit()*using both GREML and LDSC models and BioVU data.

|  | GREML model with computed GRM - Heritability (SE) | LDSC model with precomputed scores- Heritability (SE) |
| --- | --- | --- |
| Male | 0.32 (0.01) | 0.21 (0.03) |
| Female | 0.36 (0.01) | 0.24 (0.01) |

We computed the genetic correlation between male and female BioVU summary statistics for height to be 0.748 (p = 5.10 × 10^−52^), as shown in Table 5, using the *SumstatGenCorr()* function. This analysis leveraged the function’s option to use a reference LD matrix computed from 1,029,876 imputed and quality-controlled SNPs from the UK Biobank dataset^39^ (see Supplemental File). We also used *SumstatGenCorr()* to assess the genetic correlation between the BioVU sex-stratified summary statistics and publicly available sex-stratified summary statistics from the UK Biobank^58^ for the raw standing height phenotype. These results are also presented in Table 5. High genetic correlations were observed, consistent with previous findings^59^.

**Table 5:** Heritability and Genetic Correlation estimated using *SumstatGenCorr()* using BioVU and UKBB summary statistics of standing height.

|  | Heritability (SE) | Genetic Correlation (SE) | P value |
| --- | --- | --- | --- |
| BioVU male | 0.207 (0.016) | 0.748 (0.049) | $5.10 \times 10^{-52}$ |
| BioVU female | 0.215 (0.014) |  |  |
|  | **Heritability** | **Genetic Correlation** | **P value** |
| UKBB male | 0.478 (0.024) | 0.890 (0.049) | $1.91 \times 10^{-73}$ |
| BioVU male | 0.207 (0.016) |  |  |
|  | **Heritability** | **Genetic Correlation** | **P value** |
| UKBB female | 0.558 (0.028) | 0.908 (0.042) | $1.20 \times 10^{-102}$ |
| BioVU female | 0.214 (0.014) |  |  |
|  | **Heritability** | **Genetic Correlation** | **P value** |
| UKBB male | 0.516 (0.024) | 0.962 (0.043) | $7.01 \times 10^{-112}$ |
| UKBB female | 0.609 (0.027) |  |  |

## Discussion

The exploration of sex-dependent genetic effects in the analysis of complex traits has gained momentum^60^, in part due to increasing awareness of sex differences in health and disease, but also due to recognition of the need to study diverse cohorts in biomedical research to reduce health disparities. As previously described^17^, a variety of approaches can be employed to assess different aspects of sex differences in genetic effects. We recommend the use of multiple approaches, particularly when the specific nature of allelic effects is unknown. The chosen approach might vary depending on the allelic effects, the size of the sample, and specific characteristics of the dataset or trait being studied.

To facilitate these analyses, we have created GXwasR, a comprehensive R package designed to enable these complex analyses. It enables sex-aware and sex-agnostic GWAS and XWAS, equipped with functionalities that span the entire analytical spectrum, from preliminary QC to association analysis strategies. It facilitates sex-combined and sex-stratified analysis methods across the genome, managing XCI intricacies. Importantly, it can integrate with other workflows at multiple stages. To demonstrate the capabilities of GXwasR, we provided several tutorials using simulated datasets, along with an applied analysis of height GWAS data from our BioVU, the VUMC biobank, showcasing its utility in investigating sex-specific genetic architecture of a complex trait. Our ultimate goal with GXwasR is to increase accessibility of these types of analyses to a broad scientific audience.

*GXwasR* is developed as an R package. To install the development version, use the command pak::pak(“boseb/GXwasR”). The source code is publicly available on GitHub at https://github.com/boseb/GXwasRunder the Creative Commons Attribution-NonCommercial 4.0 International Public License. The scripts for running all functions and accompanying tutorials are provided in the Supplementary File.

## Data Availability

The data that support the findings of this study are available from Vanderbilt University Medical Center but restrictions apply to the availability of these data, which were used under license for the current study, and so are not publicly available. Data are, however, available from the authors upon reasonable request and with permission of Vanderbilt University Medical Center. The data in question must first be reviewed by the Integrated Data Access and Services Core to ensure that the de-identification is complete and no potentially identifying information remains. Please contact the Vanderbilt Institute for Clinical and Translational Research for more information.

## Acknowledgements

We would like to thank Ekaterina Khramtsova and Catherine Stanhope for helpful comments during the development of the package.

This research was supported in part through the computational resources and staff contributions provided by the Genomics Compute Cluster which is jointly supported by the Feinberg School of Medicine, the Center for Genetic Medicine, and Feinberg’s Department of Biochemistry and Molecular Genetics, the Office of the Provost, the Office for Research, and Northwestern Information Technology. The Genomics Compute Cluster is part of Quest, Northwestern University’s high performance computing facility, with the purpose of advancing research in genomics.

