## Supplementary File for "GXwasR: A Toolkit for Investigating Sex-Differentiated Genetic Effects on Complex Traits"

LD Pruning: 57
Filter out regions of known high-LD structure: 58
Genotype Data Processing for Merge: 58
Correcting Mismatches: 58
Handling Allele Flips: 58
Handling Sex Chromosomes: 58
Removing Mismatches Post-Flipping: 58
Principal Component Analysis (PCA): 58
Outlier Detection and Analysis: 58
Setting Outlier Thresholds: 59
Visualization and Reporting: 59
AncestryCheck Usage 59
Tutorial: Running post-imputation QC using GXwasR 61
Post-imputation QC steps 61
Post-imputation QC pipeline 62
Filtering Ambiguous SNPs and indels 62
Filter PAR, XTR and Ampliconic Regions 62
Preparing autosomal and X-chromosomal plink binary files 65
Merging the autosomal regions with PAR, XTR and Ampliconic regions 65
Flag the SNPs in the controls with a sex difference in MAF 66
Prepare separate male and female subsets of the genotypes 66
Remove SNPs with MAF < 0.05 from each sex from the sex-specific autosomal PLINK files 67
Combine sex-specific autosomal QC-ed files using common SNPs: 69
Obtain sex-specific X chromosomal PLINK files 69
QC of sex-specific X-chromosomal SNPs 70
Combine sex-specific QC-ed X chromosomal dataset using common SNPs 73
Test for HWE across the X chromosome in females 73
Check for Xchromosome SNPs with sex difference in MAF in controls 74
Combine autosomal and X chromosomal QC-ed datasets 75
Prepare separate male and female genotype datasets for sample level QC 76
Sex-specific sample level QC 76
Combine male and female-specific QC-ed genotype datasets 79
Summary of the final QC-ed genotype dataset 79

### Description of GXwasR functions

The GXwasR tool consists of thirty-three distinct functions organized into six main categories: A) Pre-imputation Quality Control (QC), B) Post-imputation QC, C) Sex-combined and sex-stratified GWAS and XWAS, tests for sex-differentiated genetic effects, (E) additional analyses (heritability estimation, genetic correlation, gene-based testing, polygenic risk scores (PRS) computation, and meta-analysis), and (F) utility functions.

#### QCsnp: Quality Control (QC) for SNPs

This function performs QC of genotype data from PLINK binary files. It can filter based on minor allele frequency, Hardy-Weinberg equilibrium, call rate, and differential missingness between cases and controls. It can also perform linkage disequilibrium-based filtering.

Table 1: Description of the input arguments of QCsnp() function

| **Arguments** | **Description** |
|---|---|
| DataDir | A character string for the file path of the input PLINK binary files. |
| ResultDir | A character string for the file path where all output files will be stored. The default is tempdir(). |
| finput | Character string, specifying the prefix of the input PLINK binary files with both male and female samples. This file needs to be in DataDir. |
| foutput | Character string, specifying the prefix of the output PLINK binary files if the filtering option for the SNPs is chosen. The default is "FALSE". |
| DataDir | A character string for the file path of the input PLINK binary files. |
| casecontrol | Boolean value, 'TRUE' or 'FALSE' indicating if the input plink files has cases-control status or not. The default is FALSE. |
| hweCase | Numeric value between 0 to 1 or NULL for removing SNPs which fail Hardy-Weinberg equilibrium for cases. The default is NULL. |
| hweControl | Numeric value between 0 to 1 or NULL for removing SNPs which fail Hardy-Weinberg equilibrium for controls. The default is NULL. |
| hwe | Numeric value between 0 to 1 or NULL for removing SNPs which fail Hardy-Weinberg equilibrium for entire dataset. The default is NULL. |
| maf | Numeric value between 0 to 1 for removing SNPs with minor allele frequency less than the specified threshold. The default is 0.05. |
| geno | Numeric value between 0 to 1 for removing SNPs that have less than the specified call rate. The default is 0.05. Users can set this as NULL for not applying this filter. |
| monomorphicSNPs | Boolean value, 'TRUE' or 'FALSE' for filtering out monomorphic SNP. The default is "TRUE". |
| caldiffmiss | Boolean value, 'TRUE' or 'FALSE', specifying whether to compute differential missingness between cases and controls for each SNP (threshold is 0.05/length(unique(No. of. SNPs in the test))). The default is TRUE. |
| diffmissFilter | Boolean value, 'TRUE' or 'FALSE', specifying whether to filter out the SNPs or only flagged them for differential missingness in cases vs contols. The deafailt is "TRUE". |
| dmissX | Boolean value, 'TRUE' or 'FALSE' for computing differential missingness between cases and controls for X chromosome SNPs only. The default is "FALSE". The diffmissFilter will work for all these SNPs. |

| | |
|---|---|
| dmissAutoY | Boolean value, 'TRUE' or 'FALSE' for computing differential missingness between cases and controls for SNPs on autosomes and Y chromosome only. The default is "FALSE". If dmissX and dmissAutoY are both FALSE, then this will be computed genome-wide. The diffmissFilter will work for all these SNPs. |
| highLD_regions | A dataframe with known high LD regions. |
| ld_prunning | Boolean value, 'TRUE' or 'FALSE' for applying linkage disequilibrium (LD)-based filtering. |
| window_size | Integer value, specifying a window size in the variant counts for LD-based filtering. The default is 50. |
| step_size | Integer value, specifying a variant count to shift the window at the end of each step for LD filtering. The default is 5. |
| r2_threshold | Numeric value between 0 to 1 of pairwise r^2 threshold for LD-based filtering. The default is 0.02. |

Usage of *QCsnp()*

QCsnp(DataDir, ResultDir = tempdir(), finput, foutput, casecontrol = TRUE, hweCase = NULL, hweControl = NULL, hwe = NULL, maf = 0.05, geno = 0.1, monomorphicSNPs = FALSE, caldiffmiss = FALSE, diffmissFilter = FALSE, dmissX = FALSE, dmissAutoY = FALSE, highLD_regions = NULL, ld_prunning = FALSE, window_size = 50, step_size = 5, r2_threshold = 0.02)

Output of *QCsnp()*

A list of two objects, namely, *MonomorSNPs* and *DiffMissSNPs* containing monomorphic SNPs and SNPs with differential missingness in cases vs controls, respectively. Output plink binary files will be stored in the working directory.

#### QCsample: QC of samples

This function identifies outlier individuals based on heterozygosity and/or missing genotype rates, which aids in the detection of samples with subpar DNA quality and/or concentration that should be excluded.

Table 2: Description of the input arguments of QCsample() function.

| **Arguments** | **Description** |
|---|---|
| DataDir | A character string for the file path of the input PLINK binary files. |
| ResultDir | A character string for the file path where all output files will be stored. The default is tempdir(). |
| finput | Character string, specifying the prefix of the input PLINK binary files with both male and female samples. This file needs to be in DataDir. |
| foutput | Character string, specifying the prefix of the output PLINK binary files if the filtering option for the SNPs is chosen. The default is "FALSE". |
| imiss | Numeric value between 0 to 1 for removing samples that have more than the specified missingness. The default is 0.03. |
| het | Positive numeric value, specifying the standard deviation from the mean heterozygosity rate. The samples whose rates are more than the specified sd from the mean heterozygosity rate are removed. The default is 3. With this default value, outlying heterozygosity rates would remove individuals who are three sd away from the mean rate. |
| small_sample_mod | Boolean value indicating whether to apply modifications for small sample sizes. Default is FALSE. |
| IBD | Numeric value for setting the threshold for Identity by Descent (IBD) analysis. Default is |

| | |
|---|---|
| | NULL. |
| IBDmatrix | Boolean value indicating whether to generate an entire IBD matrix. Default is FALSE. In this case filtered IBD matrix will be stored. |
| ambi_out | Boolean value indicating whether to process ambiguous samples. Default |
| legend_text_size | Integer, specifying the size for legend text in the plot. |
| legend_title_size | Integer, specifying the size for the legend title in the plot. |
| axis_text_size | Integer, specifying the size for axis text in the plot. |
| axis_title_size | Integer, specifying the size for the axis title in the plot. |
| title_size | Integer, specifying the size of the title of the plot heterozygosity estimate vs missingness across samples. |
| filterSample | Boolean value, 'TRUE' or 'FALSE' for filtering out the samples or not (i.e., only flagged). The default is "TRUE". |

Usage of QCsample ():

QCsample(DataDir, ResultDir, finput, foutput = NULL, imiss, het, small_sample_mod = FALSE, IBD, IBDmatrix = FALSE, ambi_out = TRUE, legend_text_size = 8, legend_title_size = 7, axis_text_size = 5, axis_title_size = 7, title_size = 9, filterSample = TRUE)

Output of QCsample():

A plot of heterozygosity vs missingness across samples and a list containing five R dataframe objects, namely, *HM* (samples with excess heterozygosity and/or missing genotype rates), *Failed_Missingness* (samples with excess missing genotypes), *Failed_heterozygosity* (samples with excess heterozygosity), Missingness_results (missingness results) and *Heterozygosity_results* (heterozygosity results) with output plink files in ResultDir if the sample filtering option is specified. *Missingness_results* contains missingness results per individual, with six columns as FID, IID, MISS_PHENO, N_MISS, N_GENO and F_MISS for Family ID, Within-family ID, Phenotype missing? (Y/N), Number of missing genotype call(s), not including obligatory missings or heterozygous haploids, number of potentially valid call(s), and missing call rate, respectively.

*Heterozygosity_results* contains heterozygosity results per individual, with six columns as FID, IID, O(HOM), E(HOM), N(NM), and F for Family ID, Within-family ID, Observed number of homozygotes, Expected number of homozygotes, Number of (non-missing, non-monomorphic) autosomal genotype observations and, Method-of moments F coefficient estimate, respectively.

#### AncestryCheck: Evaluation of the samples' ancestry label.

This function displays the result of the ancestry analysis in a color-coded scatter plot of the first two genotype principal components for study samples and reference populations. Specifically, it compares the provided study samples' ancestry labels to a panel representing a reference population, and it also flags outlier samples with respect to a chosen reference population. The function first filters the reference and study data for non-A-T or G-C SNPs. It then performs LD pruning, corrects chromosome mismatches such as incorrect chromosome numbers or base-pair positions relative to the reference dataset, checks for allele flips, updates genomic coordinates, and flips alleles as needed.. The two datasets are then joined, and then Principal Component Analysis (PCA) is performed on the resulting genotype dataset. The detection of population structure down to the level of the reference dataset can then be accomplished using PCA on this combined genotyping panel. This function uses the HapMap phase 3 data in NCBI 36 and 1000GenomeIII in CGRCh37. Both study and reference datasets need to be on the same genome build. If not, users need to lift over one of the datasets to the same build. The details on using this function are provided in the next section “Decoding Ancestry”.

Table 3: Description of the input arguments of AncestryCheck() function.

| Arguments | Description |
|---|---|
| DataDir | A character string for the file path of the input PLINK binary files. |
| ResultDir | A character string for the file path where all output files will be stored. The default is tempdir(). |
| finput | Character string, specifying the prefix of the input PLINK binary files with both male and female samples. This file needs to be in DataDir. |
| reference | Boolean value,'HapMapIII_NCBI36' and 'ThousandGenome', specifying Hapmap Phase3[1] and 1000 Genomes phase III[2] reference population, respectively. The default is 'HapMapIII_NCBI36'. |
| filterSNP | Boolean value, 'TRUE' or 'FALSE' for filtering out the SNPs. The default is TRUE. |
| studyLD | Boolean value, 'TRUE' or 'FALSE' for applying linkage disequilibrium (LD)-based filtering on study genotype data. |
| studyLD_window_size | Integer value, specifying a window size in variant count or kilobase for LD-based filtering of the the study data. |
| studyLD_step_size | Integer value, specifying a variant count to shift the window at the end of each step for LD filtering of the study data. |
| studyLD_r2_threshold | Numeric value between 0 to 1 of pairwise r^2 threshold for LD-based filtering of the study data. |
| referLD | Boolean value, 'TRUE' or 'FALSE' for applying linkage disequilibrium (LD)-based filtering of reference genotype data. |
| referLD_window_size | Integer value, specifying a window size in variant count or kilobase for LD-based filtering of the reference data. |
| referLD_step_size | Integer value, specifying a variant count to shift the window at the end of each step for LD filtering for the reference data. |
| referLD_r2_threshold | Numeric value between 0 to 1 of pairwise r^2 threshold for LD-based filtering of the reference data. |
| highLD_regions | A dataframe with known high LD regions. |
| study_pop | A dataframe containing two columns for study in first column, sample ID (i.e., IID) and in second column, the ancestry label. |
| outlier | Boolean value, 'TRUE' or 'FALSE', specifying to perform outlier detection or not. |
| outlierOf | Chracter string, specifying the reference ancestry name for detecting outlier samples. The default is "outlierOf = "CEU". |
| outlier_threshold | Numeric value, specifying the threshold to detect outlier samples. This threshold will be multiplied with the Eucledean distance from the center of the PC1 and PC2 to the maximum Euclidean distance of the reference samples. Study samples exceeding this distance will be considered outliers. The default is 3. |

#### Usage of AncestryCheck():

AncestryCheck (DataDir, ResultDir = tempdir(), finput, reference = c("HapMapIII_NCBI36", "ThousandGenome"), filterSNP = TRUE, studyLD = TRUE, studyLD_window_size = 50, studyLD_step_size = 5, studyLD_r2_threshold = 0.02, referLD = FALSE, referLD_window_size = 50, referLD_step_size = 5, referLD_r2_threshold = 0.02, highLD_regions, study_pop, outlier = FALSE, outlierOf = "CEU", outlier_threshold = 3)

#### Output of AncestryCheck():

A dataframe with the IDs of the outlier samples.

##### SexCheck: Compare sex assignments in the input plink files with those imputed from X chromosome inbreeding coefficients

This function compares provided sample sex assignments with those predicted from X chromosome inbreeding coefficients following Purcell et. al.,(2007)[3], and provides the option to convert sex assignments to the predicted valuesThis function implicitly computes the observed and expected counts of autosomal homozygous genotypes for each sample and reports method-of-moments estimates of the inbreeding coefficient (F), calculated as (observed homozygous count – expected count) divided by (total observations – expected count). The expected counts will be based on imputed minor allele frequencies. Since imputed MAFs are highly inaccurate when there are few samples, the 'compute freq' parameter should be set to TRUE to compute MAF implicitly. Due to the use of allele frequencies, if a cohort is comprised of individuals of different ancestries, users may need to process any samples with rare ancestry individually if the dataset has a very unbalanced ancestry distribution. It is advised to run this function with all the parameters set to zero, then examine the distribution of the F estimates (there should be a clear gap between a very tight male clump on the right side of the distribution and the females everywhere else). Then, rerun the function with the parameters that correspond to this gap.

Table 4: Description of the input arguments of SexCheck() function.

| **Arguments** | **Description** |
|---|---|
| DataDir | A character string for the file path of the input PLINK binary files. |
| ResultDir | A character string for the file path where all output files will be stored. The default is tempdir(). |
| finput | Character string, specifying the prefix of the input PLINK binary files with both male and female samples. This file needs to be in DataDir. |
| impute_sex | Boolean value, 'TRUE' or 'FALSE', specifying sex to be imputed or not. If 'TRUE' then seximputed PLINK files, prefixed, 'seximputed_plink', will be produced in DataDir. |
| compute_freq | Boolean value, 'TRUE' or 'FALSE', specifying minor allele frequency (MAF). This function requires MAF estimates, so it is essential to use compute_freq = TRUE for computing MAF from an input PLINK file if there are few samples in the input dataset. The default is FALSE. |
| LD | Boolean value, 'TRUE' or 'FALSE' for applying linkage disequilibrium (LD)-based filtering. The default is TRUE. |
| LD_window_size | Integer value, specifying a window size in variant count for LD-based filtering. The default is 50. |
| LD_step_size | Integer value, specifying a variant count to shift the window at the end of each step for LD filtering. The default is 5. |
| LD_r2_threshold | Numeric value between 0 to 1 of pairwise r^2 threshold for LD-based filtering. The default is 0.02. |
| fmax_F | Numeric value between 0 to 1. Samples with F estimates smaller than this value will be labeled as females. The default is 0.2. |
| mmin_F | Numeric value between 0 to 1. Samples with F estimates larger than this value will be labeled as males. The default is 0.8. |

Usage of SexCheck():

SexCheck(DataDir, ResultDir = tempdir(), finput, impute_sex = FALSE, compute_freq = FALSE, LD = TRUE, LD_window_size = 50, LD_step_size = 5, LD_r2_threshold = 0.02, fmax_F = 0.2, mmin_F = 0.8)

Output of SexCheck():

A dataframe with six columns:FID(Family ID), IID(Individual ID), PEDSEX(Sex as determined in pedigree file (1=male, 2=female)), SNPSEX(Sex as determined by X chromosome), STATUS(Displays "PROBLEM" or "OK" for each individual) and F(The actual X chromosome inbreeding (homozygosity) estimate). A PROBLEM arises if the provided and inferred sexes differ, or if the SNP data or pedigree data are ambiguous with respect to sex.

##### Xhwe: Filter X-chromosome variants for HWE in females.

This function is a part of the post-imputation quality control. This tests for Hardy-Weinberg Equilibrium (HWE) for X-chromosome variants in females. Males' hemizygous X chromosome prevents testing for HWE on their haploid X calls, and testing for HWE across all samples would have a high failure rate. This function will check for HWE across the X in females (cases and controls combined), following the recommendation in Khramtsova et al., 2023, and can remove these regions from analysis in all samples. The p-value threshold for filtering SNPs is 0.05/no.of. X-chromosome variants.

Table 5: Description of the input arguments of Xhwe() function.

| **Arguments** | **Description** |
|---|---|
| DataDir | A character string for the file path of the input PLINK binary files. |
| ResultDir | A character string for the file path where all output files will be stored. The default is tempdir(). |
| finput | Character string, specifying the prefix of the input PLINK binary files with both male and female samples. This file needs to be in DataDir. |
| foutput | Character string, specifying the prefix of the output PLINK binary files if the SNP filtering option is chosen. The default is "FALSE". |
| filterSNP | Boolean value, 'TRUE' or 'FALSE' for filtering X-chromosome variants i.e., SNPs from the input file or not (i.e., only flagged). The default is "FALSE". |

Usage of Xhwe():

Xhwe(DataDir, ResultDir = tempdir(), finput, filterSNP = TRUE, foutput)

Output of Xhwe():

A list object containing SNPs. If filterSNP = TRUE, the output filtered PLINK binary files will be produced inside DataDir.

##### MAFdiffSexControl: Test for significantly different minor allele frequency (MAF) between sexes in control samples.

With parameters to filter SNPs and/or flag SNPs, this function tests for significantly different MAF (p-value < 0.05/no. of SNPs) between sexes in control samples solely for binary phenotypes. Since the disparities may be caused by technical confounding or sample biases in the cohorts, it is advised that any SNPs in the controls with a sex difference in MAF be identified and carefully evaluated[4]. Sex differences in MAF are not anticipated for autosomal variants.

Table 6: Description of the input arguments of MAFdiffSexControl() function.

| **Arguments** | **Description** |
|---|---|
| DataDir | A character string for the file path of the input PLINK binary files. |

| | |
|---|---|
| ResultDir | A character string for the file path where all output files will be stored. The default is tempdir(). |
| finput | Character string, specifying the prefix of the input PLINK binary files with both male and female samples. This file needs to be in DataDir. |
| foutput | Character string, specifying the prefix of the output PLINK binary files if the SNP filtering option is chosen. The default is NULL. |
| filterSNP | Boolean value, 'TRUE' or 'FALSE' for removing X-chromosome variants i.e., SNPs from the input file or not (i.e., only flagged). The default is "FALSE". |

Usage of MAFdiffSexControl():

MAFdiffSexControl(DataDir, ResultDir = tempdir(), finput, foutput, filterSNP = NULL)

Output of Xhwe():

A list object containing excluded or flagged SNPs. If filterSNP = TRUE, the output filtered PLINK binary files will be produced inside DataDir.

##### FilterRegion: Filter chromosomal regions.

This function removes variants from the chromosome X Pseudo-Autosomal Region (PAR), X-transposed region (XTR), Ampliconic regions, or other user-defined regions from input plink files. Only one filtering approach can be applied at a time: either by region (using regionfile = TRUE), by chromosome (using filterCHR), or by predefined genomic categories such as filterPAR, XTR, or Ampliconic.

Table 7: Description of the input arguments of FilterRegion() function.

| **Arguments** | **Description** |
|---|---|
| DataDir | A character string for the file path of the input PLINK binary files. |
| ResultDir | A character string for the file path where all output files will be stored. The default is tempdir(). |
| finput | Character string, specifying the prefix of the input PLINK binary files with both male and female samples. This file needs to be in DataDir. |
| foutput | Character string, specifying the prefix of the output PLINK binary files if the SNP filtering option is chosen. The default is NULL. |
| CHRX | Boolean value, 'TRUE' or 'FALSE' to filter/flag regions from chromosome X. The default is TRUE. Note: CHRX only in effect if one of PAR, XTR or Ampliconic filter is in effect. |
| CHRY | Boolean value, 'TRUE' or 'FALSE' to filter/flag regions from chromosome Y The default is FALSE. Note: CHRY only in effect if one of the PAR, XTR or Ampliconic filters is in effect. |
| filterPAR | Boolean value, 'TRUE' or 'FALSE' to filter out PARs from input plink file. The default is TRUE. |
| filterXTR | Boolean value, 'TRUE' or 'FALSE' to filter out XTRs from input plink file. The default is TRUE. |
| filterAmpliconic | Boolean value, 'TRUE' or 'FALSE' to filter out Ampliconic regions from input plink file. The default is TRUE. |
| regionfile | Character string, specifying the name of the .txt file containing the user-defined regions to be filtered out from input plink file in bed format. The default is FALSE. If regionfile = TRUE, only this filtering will be in effect. Also, PAR, XTR and Ampliconic SNPs from X-chromosome will be flagged and returned. |
| filterCHR | Vector value with positive integer, specifying the chromosome number to filter/flag the SNPs. The default is 0, corresponding to no filtering based on chromosome. For non-zero values of this argument, the function will |

| | |
|---|---|
| | only consider the chromosome code to filter or flag. All other filtering will not work. If filterCHR = TRUE, only this filtering will be in effect. Also, PAR, XTR and Ampliconic SNPs from X-chromosome will be flagged and returned. |
| Hg | Character value, '19', or '38', specifying which genome build to use for PAR, XTR and Ampliconic regions. The default is Hg = "19". |
| exclude | Boolean value, 'TRUE' or 'FALSE' to filter and flag, or only flag the SNPs. The default is TRUE. |

Usage of FilterRegion():

FilterRegion(DataDir, ResultDir, finput, foutput, CHRX = TRUE, CHRY = FALSE, filterPAR = TRUE, filterXTR = TRUE, filterAmpliconic = TRUE, regionfile = FALSE, filterCHR = NULL, Hg = "19", exclude = TRUE)

Output of FilterRegion():

A list of three dataframes: *PAR* containing SNPs from PAR regions; *XTR* containing SNPs from XTR region and *Ampliconic* containing SNPs from Ampliconic region.

For non-zero value of filterCHR, a dataframe containing the excluded/flagged SNPs will be returned.

For exclude = TRUE, two sets of plink binary files will be produced in ResultDir. One set will contain the remaining SNPs after filtering, with the other containing the discarded SNPs.

##### GXwas: Running genome-wide association study (GWAS) and Xchromosome-wide association study (XWAS) models.

This function runs GWAS models for autosomes with several alternative XWAS models, such as "FMcombx01","FMcombx02", and "FMstratified" that can be applied to both binary and quantitative traits, while "GWAcxci" can only be applied to a binary trait. For binary and quantitative traits, this function uses logistic and linear regression respectively, allowing for inclusion of covariates and interactions with those covariates in a multiple regression framework. These models are test additive effects, with each additional minor allele's influence represented by the magnitude and direction of the regression coefficient. This function attempts to identify multi-collinearity among predictors by displaying NA for the test statistic and a p-value for all terms in the model. As more terms are included in the model, the likelihood of these issues increases. For details about the different XWAS models, see REF.

Table 8: Description of the input arguments of GXwas() function.

| **Arguments** | **Description** |
|---|---|
| DataDir | Character string for the file path of the input PLINK binary files. |
| ResultDir | Character string for the folder path where the outputs will be saved. |
| finput | Character string, specifying the prefix of the input PLINK binary files with both male and female samples. This file needs to be in DataDir. Note: Case/control phenotypes are expected to be encoded as 1=unaffected (control), 2=affected (case); 0 is accepted as an alternate missing value encoding. The missing case/control or quantitative phenotypes are expected to be encoded as 'NA'/'nan' (any capitalization) or -9. |
| trait | Boolean value, 'binary' or 'quantitative' for the trait. |
| standard_beta | Boolean value, 'TRUE' or 'FALSE' in case of quantitative trait for standardizing the trait or phenotype values (mean 0, unit variance), so the resulting coefficients will be standardized. The default is TRUE. |

| | |
|---|---|
| xmodel | Models "FMcombx01","FMcombx02",and "FMstatrified" can be specified for both binary and quantitative traits while "GWAcxci" can only apply to a binary trait. These models differ in how the X-chromosomal variants are handled. Three female genotypes are coded by 0, 1, and 2 in FMcombx01and FMcombx02. In the FMcombX01 model, which assumes random X-chromosome inactivation (XCI-R), the two genotypes in males are coded as 0 and 1. Similarly, in the FMcombX02 model, which assumes the X-chromosome escapes inactivation (XCI-E), male genotypes are also coded as 0 and 1. While males do not undergo XCI, these models apply different assumptions about how female genotypes are modeled and compared to males in combined analyses. To reflect dosage compensation between males and females, FMcombx02 treats men as homozygous females. In the FMcombx01 and FMcombx02 methods, associations are tested separately for males and females, and then a per SNP combined p value is computed using the Fisher's method, Fisher's method with permutation, or Stouffer's method[5–8]. Specification of an X-chromosome inactivation (XCI) pattern, or coding technique for X-chromosome variants in each sex, is not required for the GWAcxci. By simultaneously accounting for four distinct X-chromosome inactivation patterns, including random inactivation (XCI-R), escape from inactivation (XCI-E), skewed inactivation toward the reference allele (XCI-SN), and skewed inactivation toward the alternate risk allele (XCI-SR), this model may maintain reasonably high statistical power[9]. Note that sex should not be provided as a covariate in the GWAcxci model. |
| sex | Boolean value, TRUE or FALSE, indicating whether sex should be included as a covariate in the association test. This option is applicable genome-wide and is particularly relevant for combined-sex analyses, including autosomal GWAS. The default is FALSE. |
| xsex | Boolean value, 'TRUE' or 'FALSE' for using sex as covariate in association test for X chromosome SNPs. The default is FALSE. This will overwrite the 'sex' argument for Xchromosome. |
| covarfile | Character string for the full name of the covariate file in .txt format. This file should be placed in DataDir. Note about the covariate file: The first column of this file should be FID, the second column should be IID and the other columns should be covariates. The primary header line should be there starting with “FID”, and “IID” followed by covariate names. If an individual is not present in the covariate file, or if the individual has a missing phenotype value (i.e. -9 by default) for the covariate, then that individual is set to missing (i.e. will be excluded from association analysis). Use the function "DummyCovar()" to generate a new covariate file with categorical variables down-coded as binary dummy variables for the covariate file with categorical variables. For instance, if a variable has K categories, K-1 new dummy variables are constructed, and the original covariate is now estimated with a coefficient for each category. |
| interaction | Boolean value, 'TRUE' or 'FALSE' for including SNP × covariate interaction term/terms from the association analysis. The default is FALSE. If a permutation procedure is chosen, then the interaction will be automatically FALSE. For specifying an interaction with the two covariates COV1 and COV2, the model will be: Y = b0 + b1.ADD + b2.COV1 + b3.COV2 + b4.ADD x COV1 + b5.ADD x COV2 + e. When interaction factors are incorporated into the model, the main effects' significance is not always determined simply; rather, it depends on the arbitrary coding of the variables. In other words, only the interaction p-value should be used for interpretion. Also, the p-values for the covariates do not represent the test for the SNP-phenotype association after controlling for the covariate. That is the first row (ADD). Rather, the covariate term is the test associated with the covariate-phenotype association. These p-values might be extremely significant (e.g. if one includes smoking as a covariate in an analysis of heart disease, etc) ,but this does not necessarily mean that the SNP has a highly significant effect. Note that, this feature is not applicable to XCGA model. |
| covartest | Vector value with "NULL","ALL" or covariate name/names to be included in the test. The default is NULL. For instance, the user can specify “AGE” and “SEX” as covartest = c(“AGE”, “SEX”) or all the covariates as covartest = c(“ALL”). |
| Inphenocov | Vector of integer values starting from 1 to extract the terms which the user wants from the above model: Y = b0 + b1.ADD + b2.COV1 + b3.COV2 + b4.ADDxCOV1 + b5.ADDxCOV2 + e. The terms will appear in order as (1) for ADD, (2) for COV1, (4) for ADD x COV1, and (5) for ADD x COV2. If the user wants to extract the terms for COV1 and ADD x COV1, they specify it as c(2,4). The default is c(“ALL”). Note that this feature is not valid for the XCGA model for the XWAS part. |

| | |
|---|---|
| combtest | Character vector specifying the method for combining male and female p-values for stratified GWAS in FM01comb and FM02comb models. The options are: "stouffer.method", "fisher.method" and "fisher.method.perm". For the fisher.method, the function for combining p-values uses a statistic, $S = -2 \sum_{i=1}^{k} \log(p_i)$ (where k is the number of summary statistics to be combined), which follows a $\chi^2$ distribution with 2k degrees of freedom[5]. For the fisher.method.perm, using p-values from sex-stratified tests, the summary statistic for combining p-values is $S = -2 \Sigma \log(p)$. A p-value for this statistic can be derived by randomly generating summary statistics[6]. Therefore, a p-value is randomly sampled from each contributing study, and a random statistic is calculated. The fraction of random statistics greater or equal to S corresponds to the final p-value.<br><br>For the stouffer.method, the function applies Stouffer's method[7] to the p-values assuming that the p-values to be combined are independent. Letting $p_1, p_2, ..., p_k$ denote the individual (one- or two-sided) p-values of the k summary statistics to be combined, the test statistic is then computed with $Z = (1/\sqrt{k}) \Sigma Z_i$ where $Z_i = \Phi^{-1}(1 - p_i)$ and $\Phi^{-1}(\cdot)$ denotes the inverse of the cumulative distribution function of a standard normal distribution. Under the joint null hypothesis, the test statistic follows a standard normal distribution which is used to compute the combined p-value. This functionality is taken from the R package *poolr*[8]. Note that only p-values between 0 and 1 are allowed to be passed to these methods.<br><br>Note: Though this parameter is enabled for both autosome GWAS and XWAS, the combining p-value after sex-stratified test is recommended to ChrX only. |
| MF.zero.sub | Small numeric value for substituting p-values of 0 in in stratified GWAS with FM01comb and FM02comb XWAS models. The default is 0.00001. As log(0) results in Inf this replaces pvalues of 0 by default with a small float. |
| B | Integer value specifying the number of permutations to be performed when applying the fisher.method.perm method in stratified GWAS with FM01comb and FM02comb XWAS models. The default is 10000. |
| MF.mc.cores | Number of cores used for fisher.method.perm in stratified GWAS with FM01comb and FM02comb XWAS models. |
| MF.na.rm | Boolean value, 'TRUE' or 'FALSE' for removing p-values of NA in stratified GWAS with FM01comb and FM02comb XWAS when using Fisher's and Stouffer's methods. The default is FALSE. |
| MF.p.corr | Character vector specifying the method for correcting the summary p-values for FMfcomb and FMscomb models. The options are "bonferroni", "BH" and "none" for Bonferroni, BenjaminiHochberg and none, respectively. The default is "none". |
| plot.jpeg | Boolean value, 'TRUE' or 'FALSE' for saving the plots in .jpeg file. The default is TRUE. |
| plotname | A character string specifying the prefix of the file for plots. This file will be saved in DataDir. The default is "GXwas.plot". |
| snp_pval | Numeric value as p-value threshold for annotation. SNPs below this p-value will be annotated on the plot. The default is 1e-08. |
| annotateTopSnp | Boolean value, 'TRUE' or 'FALSE. If TRUE, it annotates the top hit on each chromosome that is below the snp_pval threshold. The default is FALSE. |
| suggestiveline | The default is 5 (for p-value 1e-05). |
| genomewideline | The default is 7.3 (for p-value 5e-08). |
| ncores | Integer value, specifying the number of cores for parallel processing. The default is 0 (no parallel computation). |

Usage of GXwas():

GXwas(DataDir, ResultDir, finput, trait = c("binary", "quantitative"), standard_beta = TRUE, xmodel = c("FMcomb01", "FMcomb02", "FMstatrified", "GWAScxci"), sex = FALSE, xsex = FALSE, covarfile = NULL, interaction = FALSE, covartest = c("ALL"), Inphenocov = c("ALL"), combtest = c("fisher.method", "fisher.method.perm", "stouffer.method"), MF.zero.sub = 1e-05, B = 10000, MF.mc.cores = 1, MF.na.rm = FALSE, MF.p.corr = "none", plot.jpeg = FALSE, plotname = "GXwas.plot", snp_pval = 1e-08, annotateTopSnp = FALSE, suggestiveline = 5, genomewideline = 7.3, ncores = 0)

Output of GXwas():

A dataframe containing GWAS summary statistics (with XWAS for X-chromosomal variants) along with Manhattan and Q-Q plots. For sex-stratified analysis, the return is a list containing three dataframes, namely, FWAS, MWAS, and MFWAS with association results from only female, only male, and combination p-values from both cohorts, respectively. These will be accompanied by Miami and Q-Q plots. The individual Manhattan and Q-Q-plots for sex-stratified tests prefixed with xmodel type will be in the DataDir.

When exclude = TRUE, two sets of PLINK binary files will be generated in the ResultDir. One set will contain the SNPs that remain after filtering, and the other will contain the SNPs that were excluded during filtering.

#### PvalComb: Combining p-values from stratified XWAS.

This function combines the p-values of two XWAS summary statistics (for instance male and female populations) applying various statistical methods (like Stouffer's method, Fisher's method) to integrate the p-values. It also generates Manhattan and Q-Q plots.

Table 9: Description of the input arguments of PvalComb() function.

| Arguments | Description |
|---|---|
| SumstatMale | R dataframe object of summary statistics of male XWAS with five mandatory columns: "CHR" (numeric chromosome code),"SNP" (variant id), "A1" (allele) ,"POS" (base-pair position) and "P" (p-value). Other columns may be present. |
| SumstatFemale | R dataframe object of summary statistics of female XWAS with five mandatory columns: "SNP", "A1", "TEST" , "POS" and "P". Other columns may be present. |
| combtest | Character vector specifying the method for combining p-values from sex-stratified GWAS models. Choices are “stouffer.method”, "fisher.method" and "fisher.method.perm". For fisher.method the function for combining p-values uses a statistic, S = -2 ∑^k /log p, which follows a χ^2 distribution with 2k degrees of freedom[5]. For fisher.method.perm, using p-values from sex-stratified tests, the summary statistic for combining p-values is S = -2 ∑ /log p. A p-value for this statistic can be derived by randomly generating summary statistics[6]. Therefore, a p-value is randomly sampled from each contributing study, and a random statistic is calculated. The fraction of random statistics greater or equal to S then gives the final p-value. |
| MF.p.corr | Character vector specifying the method for correcting the summary p-values for FMfcomb and FMscomb models. Choices are "bonferroni", "BH" and "none" for Bonferroni, BenjaminiHochberg and none, respectively. The default is "none". |
| MF.zero.sub | Small numeric value for substituting p-values of 0 in GWAS summary statistics. The default is 0.00001. As log(0) results in Inf this replaces p-values of 0 by default with a small float. |
| MF.na.rm | Boolean value, 'TRUE' or 'FALSE' for removing p-values of NA in stratified GWAS summary statistics when using Fisher’s and Stouffer’s methods. The default is TRUE. |
| MF.mc.cores | Number of cores used for fisher.method.perm for combining p-values. The default is 1. |
| B | Integer value specifying the number of permutations to be performed when using fisher.method.perm method. The default is 10000. |
| plot.jpeg | Boolean value, 'TRUE' or 'FALSE' for saving the plots in .jpeg file. The default is TRUE. |
| plotname | A character string specifying the prefix of the file for plots. This file will be saved in DataDir. The default is "GXwas.plot". |

| | |
|---|---|
| suggestiveline | Numeric value for suggestive significance threshold in the GWAS Manhattan plot. The default is 5 (corresponding to p-value 1e-05). |
| genomewideline | Numeric value for genome-wide significance threshold in GWAS Manhattan plot. The default is 7.3 (corresponding to p-value 5e-08). |
| ncores | Integer value, specifying the number of cores for parallel processing. The default is 0 (no parallel computation). |
| PlotDir | A character string specifying the path of the directory where the plots will be saved. The default is tempdir(). |
| snp_pval | Numeric value as p-value threshold for annotation. SNPs with p-value ≤ snp_pval will be annotated on the plot. The default is 1e-08. |
| annotateTopSnp | Boolean value, 'TRUE' or 'FALSE. If TRUE, it only annotates the top hit per chromosome where p-value ≤ snp_pval. The default is FALSE. |

Usage of PvalComb():
PvalComb(SumstatMale, SumstatFemale, combtest, MF.p.corr = "none", MF.zero.sub = 1e-05, MF.na.rm = TRUE, MF.mc.cores = 1, B = 1000, plot.jpeg = TRUE, plotname = "GXwas.plot", PlotDir = tempdir(), snp_pval, annotateTopSnp = FALSE, suggestiveline = 5, genomewideline = 7.3, ncores = 0 )

Output of PvalComb():

A dataframe with GWAS summary statistics (with XWAS for X-chromosomal variants) along with Manhattan and QQ plots.

##### SexDiff: Sex difference in effect size for each SNP using t-test.

This function uses the GWAS summary statistics from sex-stratified tests like FM01comb or FM02comb, to evaluate the difference in effect size between males and females for each SNP using a t-test.

Note, to limit results to only X chromosome variants the input dataframes should only include X-chromosome variants.

Table 10: Description of the input arguments of SexDiff() function.

| **Arguments** | **Description** |
|---|---|
| Mfile | R dataframe of summary statistics of GWAS or XWAS of male samples with six mandatory columns, SNP(Variant), CHR(Chromosome number), BP(Base pair position), A1(Minor allele), BETA_M(Effect size) and SE_M(Standard error). This can be generated by running FMcombx01 or FMcombx02 model with GXwas function. |
| Ffile | R dataframe of summary statistics of GWAS or XWAS of female samples with six mandatory columns, SNP, CHR, BP, A1(Minor allele), BETA_M and SE_M. This can be generated by running FMcombx01 or FMcombx02 model with GXwas function. |

Usage of SexDiff():
SexDiff(Mfile, Ffile)

Output of SexDiff ():

R dataframe with seven columns, SNP(Variant),CHR(Chromosome number) ,BP(Base pair position),A1(Minor allele), tstat(t-statistics for effect-size test), P(p-value) and adjP (Bonferroni corrected p-value).

#### SexDiffZscore: Z-score-based sex difference test.

This function tests for sex differences in any kind of metric,(e.g., SNP heritability, and GWAS SNP effects [β values]) using a Z-score and its associated p-value. When the ratio of the effect estimate to its standard error (STAT/SE) is normally distributed and the SNPs are independent, the test is well calibrated. If the statistics are positively correlated, this test is conservative. A user might choose to define SNPs with sex-differential effects as those variants at the extreme ends of the distribution with an absolute value of the Z-score greater than 3(|Z-score| > 3), which is roughly equivalent to $p < 10^{-3}$, and represents 0.3% of tested SNPs. Note, to limit results to only X chromosome variants, the input dataframes should only include X-chromosome variants.

Table 11: Description of the input arguments of SexDiffZscore() function.

| Arguments | Description |
|---|---|
| inputdata | A dataframe with five columns, such as, 'ID' (i.e., SNP ID or the phenotype of interest, etc.), 'Fstat' (i.e., the measured statistics in females), 'Fse' (i.e., the standard error of the measured statistics in females), 'Mstat' (i.e., the measured statistics in males), 'Mse' (i.e., the standard error of the measured statistics in males). |

Usage of SexDiffZscore():
SexDiffZscore(inputdata)

Output of SexDiffZscore():

R dataframe with columns, 'Trait','Stat','SE', 'P0'(p-value for deviation from zero test) and 'P1'(p-value for deviation from 1 test)

#### DiffZeroOne: Assessing the Z-score for deviation from one and zero.

This function tests the null hypothesis that measured statistics (example: genetic correlation, rg for a trait) < 1 using a 1-tailed test, compared to a normal distribution (z = (1 − measure statistics)/Standard error). For multiple tests (e.g., multiple traits), users are encouraged to apply a Bonferroni multiple-testing correction.

Table 12: Description of the input arguments of DiffZeroOne() function.

| Arguments | Description |
|---|---|
| inputdata | A dataframe object, containing three columns, namely, 'Trait' (phenotype of interest), 'Stat'(measured statistics) and 'SE'(standard error of the measured statistics). |
| diffzero | Boolean value, 'TRUE' or 'FALSE', specifying to perform diviation from 0 test. |
| diffone | Boolean value, 'TRUE' or 'FALSE', specifying to perform diviation from 1 test. |

Usage of DiffZeroOne():
DiffZeroOne(inputdata, diffzero = TRUE, diffone = TRUE)

Output of DiffZeroOne():

R dataframe with columns, 'Trait', 'Stat', 'SE', 'P0'(p-value for deviation from zero test) and 'P1'(p-value for deviation from 1 test)

**TestXGene: Performing gene-based association test using GWAS/XWAS summary statistics.**

This function performs gene-based association tests using GWAS/XWAS summary statistics and SNP-SNP correlation matrices. For SNP-SNP correlation matrices, users have the flexibility to use either original genotype data of their study cohort or 1000 Genomes Phase 3 reference genotype data. Users also have options to define customized SNP sets. This function computes gene-wise SNP-SNP correlation matrices and can perform nine different gene-based tests, such as, “BT"(burden test), "SKAT"(sequence kernel association test), "SKATO"(combination of BT and SKAT), "sumchi"(sum of χ2-statistics), "ACAT"(aggregated Cauchy association test for combining P values), "PCA"(principal component approach), "FLM"( functional multiple linear regression model), "simpleM" (Bonferroni correction test), "minp" (minimum Pvalue) leveraging PLINK1.9[3]and sumFREGAT[10,11] tools. This function can be applied for chrX or genome-wide

Table 13: Description of the input arguments of TestXGene() function.

| Arguments | Description |
|---|---|
| DataDir | A character string for the file path of the all the input files. |
| ResultDir | A character string for the file path where all output files will be stored. The default is tempdir(). |
| finput | Character string, specifying the prefix of the input PLINK binary files for the genotype data. This file is used to compute the correlation between SNPs. This file needs to be in DataDir. If the original study genotype data is unavailable, then users can use the 1000 Genomes Project samples. Users should use the population that most closely represents the genetic ancestry of the original study sample. For ACAT model, this parameter is not mandatory and could be set NULL. |
| sumstat | A dataframe object containing GWAS summary statistics. When the base-genotype data is used to compute genetic correlations, the mandatory columns are Column 1: "CHROM" (i.e., chromosome number), Column 2: "POS" (i.e., base-pair position), Column 3: "ID" (i.e. SNP IDs), Column 4: “P” (i.e., p-values), Column 5: “BETA” (i.e., effect-size), Column 6: "A1" (i.e., effect allele for which the effect size is estimated), Column 7: “A2” (i.e., alternative allele) and Column 8: "EAF" (i.e., the effect allele frequency) are mandatory when base-genotype data is used to compute genetic correlations. Otherwise, if the users are using reference data, then columns 5 to 8 are optional. Also, in that case, columns, such as "REF" (i.e., reference allele), and "ALT" (i.e., alternative allele) could be present to compare alleles with those in the reference file and exclude genetic variants if alleles do not match. There could be an additional column, “ANNO” with functional annotations (like "intron_variant", "synonymous", "missense" etc.) |
| gene_file | Character string, specifying the prefix of the name of a .txt file listing genes in refFlat format. This file needs to be in DataDir. The X-linked gene files, "Xlinkedgenes_hg19.txt" and "Xlinkedgenes_hg38.txt" and autosomal gene files, “Autosomes_hg19.txt” and “Autosomes_hg38.txt” can be specified. The default is "Xlinkedgenes_hg19.txt". The genome build should match the analysis. |
| gene_range | Integer value, specifying the upstream and downstream range (in kilobases) of a gene for SNPs to be considered. The default is 500000. |
| kernel_p_method | Character string, specifying the method for computing the p-value in kernel-based tests, such as SKAT, SKATO and sumchi. Available methods are "kuonen", "davies" and "hybrid"[10,11]. The default is "kuonen". |
| score_file | Character string, specifying the prefix of a file which will be used to produce score files with Z scores from p-values and beta input from GWAS summary statistics. |

| | |
|---|---|
| ref_data | Character string, specifying the path to a reference dataframe with additional data needed to recode user data according to correlation matrices that will be used. It contains "ID" column with names of SNPs, "REF" and "ALT" columns with alleles that were coded as 0 and 1, respectively. Effect sizes from data will be inverted for variants with effect alleles different from "ALT" alleles in reference data. If provided, the "REF" and "ALT" columns in the input data will be used to identify and resolve variants whose alleles differ from those in the reference dataset. This dataframe can also be a source of SNP base-pair position and allele frequencies if they are not present in the data file. "AF" column in the reference file represents the allele frequency of "ALT" allele. The default is "ref1KG.MAC5.EUR_AF.RData" containing allele frequencies of European samples from the 1000 Genomes dataset. |
| max_gene | Positive integer value, specifying the number of genes for which the gene-based test will be performed. The default is NULL to consider all genes. |
| sample_size | Positive integer value, specifying the sample size of the GWAS. Only needed for FLM and PCA models. |
| genebasedTest | Character string, specifying the name of the gene-based test. Nine different tests can be specified, "SKAT", "SKATO", "sumchi", "ACAT", "BT", "PCA", "FLM", "simpleM", "minp". The default is "SKAT". |
| gene_approximation | Boolean value, 'TRUE' or 'FALSE', specifying whether approximation for large genes (>= 500 SNPs) should be used. Applicable for SKAT, SKATO, sumchi, PCA, FLM. The default is TRUE for these methods). |
| beta_par | Boolean value, 'TRUE' or 'FALSE', specifying whether approximation for large genes (>= 500 SNPs) should be used. Applicable for SKAT, SKATO, sumchi, PCA, FLM (default = TRUE for these methods). |
| weights_function | A function of MAF to assign weights for each genetic variant. By default is NULL. In this case the weights will be calculated using the beta distribution. |
| geno_variance_weights | Character string, indicating whether scores should be weighted by the variance of genotypes: "none" (i.e., no weights applied, resulting in a sum chi-square test);<br><br>"se.beta" (i.e., scores weighted by variance of genotypes estimated from p-values and effect sizes); "af" (i.e., scores weighted by variance of genotypes calculated as AF * (1 AF), where AF is allele frequency. |
| acc_devies | Positive numeric value, specifying the accuracy parameter for "davies" method. The default is 1e-8. |
| lim_devies | Positive numeric value, specifying the limit parameter for "davies" method. The default is 1e+6. |
| rho | Logical value, 'TRUE' or 'FALSE' or can be a vector of grid values from 0 to 1. If TRUE, the optimal test (SKAT-O) is performed[11]. The default grid is c(0, 0.1^2, 0.2^2, 0.3^2,<br><br>0.4^2, 0.5^2, 0.5, 1). |
| skato_p_threshold | Positive numeric value, specifying the largest p-value that will be considered as important when performing computational optimization in SKAT-O. All p-values larger than skato_p_threshold will be processed via burden test. The default is 0.8 |
| anno_type | A character (or character vector) indicating annotation types to be used. The default is "" (i.e, nothing). |
| mac_threshold | Integer value, specifying the threshold of MACs (Minor allele content) calculated from MAFs. In ACAT, scores with MAC <= 10 will be combined using Burden test. |

| | |
|---|---|
| reference_matrix_used | Boolean value, 'TRUE' or 'FALSE' logical indicating whether the correlation matrices were generated using the reference matrix. The default is FALSE. If TRUE, regularization algorithms will be applied to ensure the invertibility and numerical stability of the matrices. |
| regularize_fun | Character string, specifying the one of two regularization algorithms if ‘reference_matrix’ is TRUE: 'LH' (default) or 'derivLH'. |
| pca_var_fraction | Positive numeric value, specifying the minimal proportion of genetic variance within the region that should be explained by principal components used in PCA method. This is also valid in 'simpleM'. The default is 0.85. |
| flm_basis_function | Character string, specifying the name of a basis function type for beta-smooth in FLM method. Can be set to "bspline" (B-spline basis) or "fourier" (Fourier basis, default). |
| flm_num_basis | Positive integer value, specifying the number of basis functions to be used for betasmooth in FLM method. The default is 25. |
| flm_poly_order | Positive integer value, specifying the polynomial order to be used in "bspline" for FLM model. The default = 4, which corresponds to the cubic B-splines. This has no effect if only Fourier bases are used |
| flip_genotypes | Logical value, 'TRUE' or 'FALSE', indicating whether the genotypes of some genetic variants should be flipped (re-labelled). The default is FALSE. |
| omit_linear_variant | Logical value, 'TRUE' or 'FALSE', indicating whether to omit linearly dependent genetic variants. It was done in the FLM test[10]. The default is FALSE. |

Usage of TestXGene():

TestXGene(DataDir, ResultDir = tempdir(), finput, sumstat, gene_file, gene_range = 5e+05, score_file, ref_data = NULL, max_gene = NULL, sample_size = NULL, genebasedTest = c("SKAT", "SKATO", "sumchi", "ACAT", "BT", "PCA", "FLM", "simpleM", "minp"), gene_approximation = TRUE, beta_par, weights_function, geno_variance_weights, kernel_p_method = "kuonen", acc_devies = 1e-08, lim_devies = 1e+06, rho = TRUE, skato_p_threshold = 0.8, anno_type = "", mac_threshold, reference_matrix_used, regularize_fun, pca_var_fraction = 0.85, flm_basis_function = "fourier", flm_num_basis = 25, flm_poly_order = 4, flip_genotypes = FALSE, omit_linear_variant = FALSE)

Output of TestXGene():

A data frame with columns "gene", "chrom", "start", "end", "markers" (i.e., numbers of SNPs), "filtered.markers" (i.e. filtered SNPs) and "pvalue" (i.e., p-value). Additionally, for “BT”, there will be “beta” (i.e., gene-level estimates of betas) and “beta.se” (i.e., standard errors of betas). For “FLM”, there will be the “model” column with the names of the functional models used for each region. Names shortly describe the functional basis and the number of basis functions used, e.g., "F25" means 25 Fourier basis functions, "B15" means 15 B-spline basis functions. For “PCA”, there will be the “ncomponents” (the number of the principal components used for each region) and “explained.variance.fraction”(i.e., the proportion of genetic variance they capture) columns.

#### MetaGWAS: Combining summary-level results from two or more GWA studies into a single estimate.

This function combines K sets of GWAS summary statistics deriving from a single (or similar) phenotype(s). This function uses PLINK's inverse variance-based analysis[3] to run a number of models, including: a) Fixed-effect model and b) Random-effect model, assuming there may be variation between the underlying effects, i.e., effect size beta. 'This function also calculates weighted Z-score-based p-values after METAL[12]. For more information about the algorithms,  refer to REF.

Table 14: Description of the input arguments of MetaGWAS() function.

| Arguments | Description |
|---|---|
| DataDir | A character string for the file path of the input files needed for 'SummData' and 'SNPfile' arguments. |
| SummData | Vector value containing the name/names of the .Rda file/files containing GWAS summary statistics, with 'SNP' (i.e., SNP idenitifier), 'BETA' (i.e., effect-size or logarithm of odds ratio), 'SE' (i.e., standard error of BETA), 'P' (i.e., p-values), 'NMISS' (i.e., effective sample size), 'L95' (i.e., lower limit of 95% confidence interval) and 'U95' (i.e., upper limit of 95% confidence interval) as mandatory column headers. These files need to be in DataDir. If the numbers of cases and controls are unequal, effective sample size should be $\frac{4}{\text{No. of cases+no.of controls}}$. A smaller "effective" sample size may be used for samples that include related individuals, however simulations indicate that small changes in the effective sample size have relatively little effect on the final p-value (3). Columns, such as, 'CHR'(Chromosome code), 'BP'(Basepair position), 'A1' (First allele code), 'A2' (Second allele code) columns are optional. If these are present, setting 'useSNPposition' to FALSE, causes 'CHR', 'BP' and 'A1' to be ignored, and setting 'UseA1' to be 'FALSE' causes A1 to be ignored. If both these arguments are 'TRUE', this function handles A1/A2 allele flips properly. Otherwise, A1 mismatches are thrown out. Values of CHR/BP are allowed to vary. |
| ResultDir | A character string for the file path where all output files will be stored. The default is tempdir(). |
| SNPfile | Character string specifying the name of the plain-text file with a column of SNP names. These could be LD clumped SNPs or any other list of chosen SNPs for meta-analysis. This file needs to be in DataDir. |
| useSNPposition | Boolean value, 'TRUE' or 'FALSE' for using 'CHR', 'BP', and 'A1' or not. The default is FALSE. Note: if this is 'FALSE', Manhattan and QQ plot will not be generated. |
| UseA1 | Boolean value, 'TRUE' or 'FALSE' for 'A1' to be used or not. The default is FALSE. |
| GCse | Boolean value, 'TRUE' or 'FALSE' for applying study specific genomic control to adjust each study for potential population structure. The default is TRUE. If users want to apply genomic control separately for directly genotyped and imputed SNPs prior using the function, set this parameter as FALSE. |
| plotname | Character string, specifying the plot name of the file containing forest plots for the SNPs. The default is "Meta_Analysis.plot". |
| pval_filter | Character value as "R","F" or "W", specifying whether p-value threshold should be chosen based on "Random", "Fixed" or "Weighted" effect model for the SNPs to be included in the forest plots. |
| top_snp_pval | Numeric value, specifying the threshold to be used to filter the SNPs for the forest plots. The default is 1e-08. |
| max_top_snps | Integer value, specifying the maximum number of top SNPs (SNPs with the lowest p-values) to be plotted in the forest plot file. The default is 6. |
| chosen_snps_file | Character string specifying the name of the plain-text file with a column of SNP names for the forest plots. The default is NULL. |
| byCHR | Boolean value, 'TRUE' or 'FALSE', specifying whether the meta-analysis will be performed separately for each chromosome or not. The default is FALSE. |

| | |
|---|---|
| pval_threshold_manplot | Numeric value, specifying the p-value threshold for plotting Manhattan plots. |

Usage of MetaGWAS():

MetaGWAS(DataDir, SummData = c(""), ResultDir = tempdir(), SNPfile = NULL, useSNPposition = TRUE, UseA1 = FALSE,GCse = TRUE, plotname = "Meta_Analysis.plot", pval_filter = "R", top_snp_pval = 1e-08, max_top_snps = 6, chosen_snps_file = NULL, byCHR = FALSE, pval_threshold_manplot = 1e-05)

Output of MetaGWAS():

A list object containing five dataframes. The first three dataframes, such as *Mfixed*, *Mrandom* and *Mweighted* contain results for fixed effect, random effect and weighted models. Each of these dataframes can have a maximum of 12 columns, such as 'CHR'(Chromosome code),'BP'(Basepair position),'SNP'(SNP identifier), 'A1'(First allele code), 'A2'(Second allele code), Q(p-value for Cochrane's Q statistic), I(I^2 heterogeneity index (0-100)), 'P(P-value from meta-analysis), ES(Effect-size estimate from meta-analysis), SE(Standard Error from meta-analysis), 'CI_L(Lower limit of confidence interval) and CI_U(Upper limit of confidence interval). The fourth dataframe contains the same columns "CHR", "BP", "SNP", "A1", "A2", "Q", "I", with column 'N'(Number of valid studies for this SNP), 'P'(Fixed-effects meta-analysis p-value), and other columns as 'Fx'(Study x (0-based input file indices) effect estimate, Examples: F0, F1 etc.). The fifth dataframe, ProblemSNP has three columns: 'File'(file name of input data), 'SNP'(Problematic SNPs that are thrown) and 'Problem'(Problem code). Problem codes are, 'BAD_CHR'(Invalid chromosome code), 'BAD_BP'(Invalid base-position), 'BAD_ES'(Invalid effect-size), 'BAD_SE' (Invalid standard error), 'MISSING_A1'(Missing allele 1 label), 'MISSING_A2'(Missing allele 2 label), 'ALLELE_MISMATCH'(Mismatching allele codes across files). A .pdf file comprising the forest plots of the SNPs is produced in the ResultDir with Plotname as prefix. If useSNPposition is set TRUE, a .jpeg file with Manhattan Plot and Q-Q plot will be in the ResultDir with Plotname as prefix.

#### ComputePRS: Computing polygenic risk score (PRS).

This function calculates the polygenic risk score, which is the total of allele counts (genotypes) weighted by estimated effect sizes from genome-wide association studies. It uses C+T (clumping + thresholding) filtering techniques. Users can perform clumping per individual chromosome and genome-wide. Also, the function offers the option to include genetic principal components and other covariates. Using this function, users can experiment with various clumping and thresholding arrangements to test a range of various parameter values.

Table 15: Description of the input arguments of ComputePRS() function.

| **Arguments** | **Description** |
|---|---|
| DataDir | A character string for the file path of the all the input files. |
| ResultDir | A character string for the file path where all output files will be stored. The default is tempdir(). |
| finput | Character string, specifying the prefix of the input PLINK binary files for the genotype data i.e., the target data for the clumping procedure. This file needs to be in DataDir. If the target dataset is small (e.g. N < 500), then users can use the 1000 Genomes Project samples. Use the population that most closely reflects represents the genetic ancestry of the study population. |
| summarystat | A dataframe object containing GWAS summary statistics. The mandatory column headers in this dataframe are 'CHR'(Chromosome code), 'BP'(basepair position), 'A1' (effect allele), ‘SNP’ (i.e., SNP identifier), ‘BETA’ or "OR"(i.e., effect-size or logarithm of odds ratio), and ‘P’ (i.e., pvalues). Note: The first three columns needed to be "SNP", "A1" and "BETA" or "OR". |
| phenofile | A character string, specifying the name of the mandatory phenotype file. This is a plain text file with no header; columns contain: family ID, individual ID, and phenotype. For a binary trait, the phenotypic value |

| | |
|---|---|
| | should be coded as 0 or 1, then it will be recognized as a case-control study (0 for controls and 1 for cases). Missing values should be represented by "-9" or "NA". The phenotype column should be labeled as "Pheno1". This file needs to be in the DataDir. |
| covarfile | A character string, specifying the name of the covariate file which is a plain text file with no header; columns contain: family ID, individual ID, and the covariates. The default is NULL. This file needs to be in DataDir. |
| effectsize | Boolean value, 'BETA' or 'OR', specifying the type of the GWAS effectsize. The default is 'BETA'. |
| ldclump | Boolean value, 'TRUE' or 'FALSE', specifying whether to perform clumping or not. |
| LDreference | A character string, specifying the prefix of the PLINK files of the population reference panel of the same ancestry, and ideally the same one that was used for imputing the study data. These files should be in DataDir. |
| clump_p1 | Numeric value, specifying the significance threshold for index SNPs if 'ldclump' was set to be TRUE. The default is 0.0001. |
| clump_p2 | Numeric value, specifying the secondary significance threshold for clumped SNPs if 'ldclump' was set to be TRUE. The default is 0.01 |
| clump_r2 | Numeric value, specifying the linkage disequilibrium (LD) threshold for clumping if 'ldclump' was set to be TRUE. The default is 0.50. |
| clump_kb | Integer value, specifying the physical distance threshold in basepairs for clumping if 'ldclump' was set to be TRUE. The default is 250. |
| byCHR | Boolean value, 'TRUE' or 'FALSE', specifying to perform per chromosome clumping if 'ldclump' was set to be TRUE. The default is TRUE. |
| pthreshold | Numeric vector, containing several p-value thresholds to maximize predictive ability of the derived polygenic scores. |
| highLD_regions | Character string, specifying the .txt file name containing coordinates of genomic regions with high LD. The default is NULL. |
| ld_prunning | Boolean value, 'TRUE' or 'FALSE' for LD-based filtering for computing genetic PCs as covariates. |
| window_size | Integer value, specifying a window size in variant count or kilobases for LD-based filtering in computing genetic PCs. The default is 50. |
| step_size | Integer value, specifying a variant count to shift the window at the end of each step for LD filtering in computing genetic PCs. The default is 5. |
| r2_threshold | Numeric value between 0 to 1 specifying thepairwise r^2 threshold for LD-based filtering in computing genetic PCs. The default is 0.02. |
| nPC | Positive integer value, specifying the number of genetic PCs to be included as predictor in the PRS model fit. The default is 6. |
| pheno_type | Boolean value, 'binary' or 'quantitative', specifying the type of the trait. The default is 'binary'. |

Usage of ComputePRS():

ComputePRS(DataDir, ResultDir = tempdir(), finput, summarystat, phenofile, covarfile = NULL, effectsize = c("BETA","OR"), ldclump = FALSE, LDreference, clump_p1, clump_p2, clump_r2, clump_kb, byCHR = TRUE, pthreshold = c(0.001,0.05,0.1,0.2,0.3,0.4,0.5), highLD_regions, ld_prunning = FALSE, window_size = 50, step_size = 5,r2_threshold = 0.02, nPC = 6,pheno_type = "binary")

Output of ComputePRS():

A list object containing a dataframe and a numeric value. The dataframe, PRS, contains four mandatory columns: IID (i.e., Individual ID), FID (i.e., Family ID), Pheno1 (i.e., the trait for PRS) and Score (i.e., the best PRS). Additional columns containing covariates are optional. The numeric value BestP indicates the p-value threshold that yielded the best-fitting PRS model.

The function also produces several plots, including: p-value thresholds vs PRS model fit and PRS distribution for male and females. For case-control data, it shows the PRS distribution among cases and controls and ROC curves as well.

#### GeneticCorrBT: Computing genetic correlation between two traits.

This function computes genetic correlation, a quantitative metric that describes the extent to which two heritable traits share genetic variance. This function can perform a bivariate Genomic-Relatedness-based Restricted Maximum Likelihood (GREML) analysis to determine the genetic correlation between two quantitative traits, two binary traits, and between a quantitative trait and a binary trait, following Yang et. al.[13] and Lee et. al[14]. The function can compute the genetic correlation genome-wide or per chromosome.

Table 15: Description of the input arguments of GeneticCorrBT() function.

| Arguments | Description |
|---|---|
| DataDir | A character string for the file path of the all the input files. |
| ResultDir | A character string for the file path where all output files will be stored. The default is tempdir(). |
| finput | Character string, specifying the prefix of the input PLINK binary files for the genotype data i.e., the data for which clumping will be performed. This file needs to be in DataDir. If the target dataset is small (e.g. N < 500) then users can use the 1000 Genomes Project samples. Use the population that most closely reflects represents the genetic ancestry of the study population. |
| byCHR | Boolean value, 'TRUE' or 'FALSE', specifying whether the analysis will be performed per chromosome or not. The default is FALSE. |
| REMLalgo | Integer value of 0, 1 or 2, specifying the algorithm to run REML iterations, 0 for average information (AI), 1 for Fisher-scoring and 2 for EM. The default option is 0, i.e. AI-REML[15]. |
| nitr | Integer value, specifying the number of iterations for performing the REML. The default is 100. |
| phenofile | A dataframe for Bivar RELM has four columns: family ID, individual ID and two trait columns. For a binary trait, the phenotypic value should be coded as 0 or 1, then it will be recognized as a case-control study (0 for controls and 1 for cases). Missing values should be represented by "-9" or "NA". |
| cat_covarfile | A character string, specifying the name of the categorical covariate file which is a plain text file with no header; columns are: family ID, individual ID, and categorical covariates. The default is NULL. This file needs to be in DataDir. |
| quant_covarfile | A character string, specifying the name of the quantitative covariate file which is a plain text file with no header; columns are: family ID, individual ID, and continuous covariates. The default is NULL. This file needs to be in DataDir. |
| computeGRM | Boolean value, 'TRUE' or 'FALSE', specifying whether to compute GRM matrices or not. The default is TRUE. |
| grmfile_name | A string of characters specifying the prefix of autosomal .grm.bin file. Users need to provide separate GRM files for autosomes and the X chromosome in ResultDir. The X chromosome GRM file should have "x" added in the autosomal prefix as file name. |

| | |
|---|---|
| | For example, if the autosomal file is "ABC.grm.bin", then the X chromosomal file should be "xABC.grm.bim". If a user is providing per chromosome GRMs, then the prefix should add "ChrNumber_" at the starting of the prefix, i.e., "Chr1_ABC.grm.bin". The default is NULL. |
| partGRM | Boolean value, 'TRUE' or 'FALSE', specifying whether the GRM will be partitioned into n parts (by row) in the GREML model. The default is FALSE. |
| autosome | Boolean value, 'TRUE' or 'FALSE', specifying whether estimate of heritability will be done for autosomes or not. The default is 'TRUE'. |
| Xsome | Boolean value, 'TRUE' or 'FALSE', specifying whether to estimate heritability for the X chromosome or not. The default is 'TRUE'. |
| nGRM | Integer value, specifying the number of the partition of the GRM in GREML model. The default is 3. |
| cripticut | Numeric value, specifying the threshold to create a new GRM of "unrelated" individuals in GREML model. The default is arbitrary chosen as 0.025. |
| minMAF | Positive numeric value (< maxMAF), specifying the minimum threshold for the MAF filter of the SNPs in the Bivariate GREML model. |
| maxMAF | Positive numeric value (minMAF,1), specifying the maximum threshold for the MAF filter of the SNPs in the Bivariate GREML model. |
| excludeResidual | Boolean value, 'TRUE' or 'FALSE', specifying whether to drop the residual covariance from the model. Recommended to set this TRUE if the traits were measured on different individuals. The default is TRUE. |
| ncores | Integer value, specifying the number of cores to be used. |

Usage of GeneticCorrBT():
GeneticCorrBT(DataDir, ResultDir, finput, byCHR = FALSE, REMLalgo = c(0,1,2), nitr = 100, phenofile, cat_covarfile = NULL, quant_covarfile = NULL, computeGRM = TRUE, grmfile_name = NULL, partGRM = FALSE, autosome = TRUE, Xsome = TRUE, nGRM = 3, cripticut = 0.025, minMAF = NULL, maxMAF = NULL, excludeResidual = FALSE, ncores = 2)

Output of GeneticCorrBT():

A dataframe containing a minimum of three columns: "Source" (i.e., source of heritability), "Variance" (i.e., estimated heritability), and "SE" (i.e., standard error of the estimated heritability). Source column has rows: V(G)_tr1 (genetic variance for trait 1), V(G)_tr2 (genetic variance for trait 2), C(G)_tr12 (genetic covariance between traits 1 and 2), V(e)_tr1 (residual variance for trait 1), V(e)_tr2 (residual variance for trait 2), C(e)_tr12 (residual covariance between traits 1 and 2), Vp_tr1 (proportion of variance explained by all SNPs for trait 1), Vp_tr2 (proportion of variance explained by all SNPs for trait 2), V(G)/Vp_tr1 (phenotypic variance for trait 1), V(G)/Vp_tr2 (phenotypic variance for trait 2), rG (genetic correlation) and n (sample size). In case of per chromosome analysis, there is a 'chromosome' column specifying the chromosome.

#### EstimateHerit: Computing SNP heritability i.e., the proportion of phenotypic variance explained by SNPs.

This function performs two types of heritability estimation, (i)GREML: Genomic relatedness matrix (GRM) restricted maximum likelihood-based method following GCTA[15] and (ii)LDSC: LD score regression-based method following[16]. Prior to using this function, users are recommended to apply QCsnp and QCsample to ensure data quality control.

Table 17: Description of the input arguments of EstimateHerit() function.

| Arguments | Description |
|---|---|
| DataDir | A character string for the file path of the all the input files. |
| ResultDir | A character string for the file path where all output files will be stored. The default is tempdir(). |
| finput | Character string, specifying the prefix of the input PLINK binary files for the genotype data. This file needs to be in DataDir. For the LDSC model, if the original genotype data is not available, Hapmap 3 or 1000 Genomes data can be used. If users specify NULL, then users must provide 'precomputedLD' argument. See below. |
| precomputedLD | A dataframe object as LD matrix with columns: "CHR", "SNP", "BP", "ld_size", "MAF", "ld_score". The default is NULL. |
| chi2_thr1 | Numeric value for threshold on chi2 in step 1 of LDSC regression. Default is 30. |
| chi2_thr2 | Numeric value for threshold on chi2 in step 2. Default is Inf (none). |
| intercept | Numeric value to constrain the intercept to some value (e.g. 1) in LDSC regression. Default is NULL. |
| summarystat | A dataframe object with GWAS summary statistics. The mandatory column headers are: 'chr'(Chromosome ID), 'pos'(basepair position), 'a1' (First allele code), 'rsid' (i.e., SNP identifier), 'beta' (i.e., effect-size or logarithm of odds ratio), 'beta_se' (i.e., standard error of beta), 'P' (i.e., p-values) and 'n_eff' (i.e., effective sample size). For a case-control study, the effective sample size should be 4 / (1/<# of cases> + 1/<# of controls>). The default is NULL. |
| ncores | Integer value, specifying the number of cores to be used for running LDSC model. The default is 2. |
| model | Character string, specifying the heritability estimation model. There are two options: "GREML" or "LDSC". The default is "GREML". Note: argument For LDSC, DataDir and finput can be NULL. |
| byCHR | Boolean value, 'TRUE' or 'FALSE', specifying whether the analysis will be performed per chromosome or not. The default is FALSE. |
| r2_LD | Numeric value, specifying the LD threshold for clumping in the LDSC model. The default is 0. |
| LDSC_blocks | Integer value, specifying the block size for performing jackknife variance estimator in the LDSC model following LDpred model[17]. The default is 200. |
| REMLalgo | Integer value of 0, 1 or 2, specifying the algorithm to run REML iterations, 0 for average information (AI), 1 for Fisher-scoring, and 2 for EM. The default option is 0, i.e. AI-REML[15]. |
| nitr | Integer value, specifying the number of iterations for performing the REML. The default is 100. |
| cat_covarfile | A character string, specifying the name of the categorical covariate file, which is a plain text file with no header; columns are: family ID, individual ID, and categorical covariates. The default is NULL. This file needs to be in DataDir. |
| quant_covarfile | A character string, specifying the name of the quantitative covariate file, which is a plain text file with no header; columns are: family ID, individual ID, and continuous covariates. The default is NULL. This file needs to be in DataDir. |

| | |
|---|---|
| prevalance | Numeric value, specifying the disease prevalence. The default is 0.01. |
| computeGRM | Boolean value, 'TRUE' or 'FALSE', specifying whether to compute GRM matrices or not. The default is TRUE. |
| grmfile_name | A string of characters specifying the prefix of autosomal .grm.bin file.<br><br>Users need to provide separate GRM files for autosomes and the X chromosome in ResultDir. The X chromosome GRM file should have "x" added in the autosomal prefix as file name. For example, if the autosomal file is named "ABC.grm.bin", then the X chromosome file should be "xABC.grm.bim". If users are providing per chromosome GRMs, then the prefix should add "ChrNumber_" at the starting of the prefix, like, "Chr1_ABC.grm.bin". The default is NULL. |
| partGRM | Boolean value, 'TRUE' or 'FALSE', specifying whether the GRM will be partitioned into n parts (by row) in GREML model. The default is FALSE. |
| autosome | Boolean value, 'TRUE' or 'FALSE', specifying whether to estimate heritability for autosomes or not. The default is 'TRUE'. |
| Xsome | Boolean value, 'TRUE' or 'FALSE', specifying whether to estimate heritability for the X chromosome or not. The default is 'TRUE'. |
| nGRM | Integer value, specifying the number of the partition of the GRM in GREML model. The default is 3. |
| cripticut | Numeric value, specifying the threshold to create a new GRM of "unrelated" individuals in GREML model. The default is chosen as 0.025[15]. |
| minMAF | Positive numeric value (< maxMAF), specifying the minimum threshold for the MAF filter of the SNPs in the GREML model. |
| maxMAF | Positive numeric value (minMAF,1), specifying the maximum threshold for the MAF filter of the SNPs in the GREML model. |
| hg | Boolean value, specifying the genome build, "hg19" or "hg38" to use chromosome length from the UCSC genome browser, and obtaining genes and proteins according to this build. The default is "hg19". |
| PlotIndepSNP | Boolean value, 'TRUE' or 'FALSE', specifying whether to use independent SNPs i.e., per chromosome LD-pruned SNPs in the plots or not. The default is TRUE. |
| IndepSNP_window_size | Integer value, specifying a window size in variant count or kilobase for LD-based filtering. The default is 50. |
| IndepSNP_step_size | Integer value, specifying a variant count to shift the window at the end of each step for LD filtering of pruned SNPs in the plots. The default is 5. |
| IndepSNP_r2_threshold | Numeric value between 0 to 1 of pairwise r^2 threshold for LD-based filtering of pruned SNPs in the plots. The default is 0.02. |
| highLD_regions | Character string, specifying the .txt file name containing coordinates of genomic regions with known high LD. This file needs to be in DataDir. |
| plotjpeg | Boolean value, 'TRUE' or 'FALSE', specifying whether to save the plots in a jpeg file in ResultDir. The default is TRUE. |
| plotname | String of character value specifying the name of the jpeg file with the plots. The default is "Heritability_Plots". |

Usage of EstimateHerit():
EstimateHerit(DataDir = NULL, ResultDir = tempdir(), finput = NULL, precomputedLD = NULL, indepSNPs = NULL,summarystat= NULL, ncores = 2, model = c("LDSC","GREML"), computeGRM = TRUE , grmfile_name = NULL,byCHR = FALSE, r2_LD = 0, LDSC_blocks = 20, intercept = NULL, chi2_thr1 = 30, chi2_thr2 = Inf, REMLalgo = c(0,1,2), nitr = 100, cat_covarfile = NULL, quant_covarfile = NULL,prevalence = NULL, partGRM = FALSE, autosome = TRUE, Xsome = TRUE, nGRM = 3, cripticut = 0.025, minMAF = NULL, maxMAF = NULL, hg = c("hg19","hg38"), PlotIndepSNP = TRUE, IndepSNP_window_size = 50, IndepSNP_step_size = 5, IndepSNP_r2_threshold = 0.02, highLD_regions = NULL, plotjpeg = TRUE, plotname = "Heritability_Plots")

Output of EstimateHeri():

A dataframe containing a maximum of eight columns for GREML (three columns if running genome-wide), and ten columns for the LDSC model if byCHR is TRUE. The columns, such as, "chromosome"(i.e., chromosome code),"snp_proportion" (i.e., per chromosome SNP propotion)", "no.of.genes" (i.e., number of genes per chromosome), "no.of.proteins" (i.e., number of proteins per chromosome),"size_mb" (i.e., chromosome length), "Source" (i.e., source of heritability), "Variance" (i.e., estimated heritability), and "SE" (i.e., standard error of the estimated heritability) are common for both GREML and LDSC model. The column, "Intercept" (i.e., LDSC regression intercept) and "Int_SE" (i.e., standard error of the intercept) will be two additional columns when running LDSC models. Source column will have rows, such as V(1) (i.e., genetic variance type), V(e) (i.e., residual variance), V(p) (i.e., phenotypic variance), V(1)/Vp (i.e., ratio of genetic variance to phenotypic variance), and V(1)/Vp_L (i.e., ratio of genetic variance to phenotypic variance in liability scale for binary phenotypes). If byCHR is FALSE, then the first five columns will not be reported in the dataframe.

#### SumstatGenCorr: Genetic Correlation Calculation from GWAS Summary Statistics.

This function calculates the genetic correlation between two sets of GWAS summary statistics using a pre-specified reference linkage disequilibrium (LD) matrix from the UK Biobank.

Table 18: Description of the input arguments of SumstatGenCorr() function.

| Arguments | Description |
|---|---|
| ResultDir | Directory where results should be saved. |
| referenceLD | Reference LD matrix identifier. These are the LD matrices and their eigen-decomposition from 335,265 genomic British UK Biobank individuals. Two sets of reference panels are provided:<br>1) 307,519 QC-ed UK Biobank Axiom Array SNPs. The size is ~7.5 GB after unzipping.<br>2) 1,029,876 QC-ed UK Biobank imputed SNPs. The size is ~31 GB after unzipping. Although it takes more time, the imputed panel provides more accurate estimates of genetic correlations.<br>Therefore, if the GWAS includes most of the HapMap3 SNPs, then the imputed reference panel should be used. |
| sumstat1 | Data frame for the first set of summary statistics.<br>The input data frame should include following columns: SNP, SNP ID; A1, effect allele; A2, reference allele; N, sample size; Z, z-score; If Z is not given, alternatively, the user may provide: b, estimate of marginal effect in GWAS; se, standard error of the estimates of marginal effects in GWAS. |
| sumstat2 | Data frame for the second set of summary statistics with the same columns as sumstat1. |

| | |
|---|---|
| Nref | Sample size of the reference sample where LD is computed. If the default UK Biobank reference sample is used, Nref = 335,265. |
| N0 | Number of individuals included in both cohorts. The estimated genetic correlation is usually robust against misspecified N0.<br>If not given, the default value is set to the minimum sample size across all SNPs in cohort 1 and cohort 2. |
| eigen.cut | Specifies which eigenvalues and eigenvectors in each LD score matrix should be used for HDL.<br>Users can specify a numeric value between 0 and 1 for eigen.cut. For example, eigen.cut = 0.99 means using the leading eigenvalues explaining 99% of the variance and their correspondent eigenvectors. If the default 'automatic' is used, the eigen.cut that provides the most stable heritability estimates will be used. |
| lim | Tolerance limitation, default lim = exp(-18). |
| parallel | Boolean value, TRUE or FALSE for whether to perform parallel computation. The default is FALSE. |
| numCores | The number of cores to be used. The default is 2. |

Usage of SumstatGenCorr():
SumstatGenCorr(ResultDir = tempdir(), referenceLD, sumstat1, sumstat2, Nref = 335265, N0 = min(sumstat1$N), eigen.cut = "automatic", lim = exp(-18), parallel = FALSE, numCores = 2)

Output of SumstatGenCorr():

A list is returned with: *rg*: the estimated genetic correlation; *rg.se*: the standard error of the estimated genetic correlation; *P*: p-value based on the Wald test; *estimates.df*: a detailed matrix that includes the estimates and standard errors of heritabilities, genetic covariance, and genetic correlation; *eigen.use*: the eigen.cut used in computation.

##### SexRegress: Performing linear regression analysis with quantitative response variable.

This function can be used to check the association between two variables e.g., PRS with sex.

Table 19: Description of the input arguments of SexRegress() function.

| **Arguments** | **Description** |
|---|---|
| response_index | Integer value, specifying the column number of the response variable. |
| fdata | R dataframe object. The column with header "response" should contain the response variable. All other columns are the regressors. |
| regressor_index | Integer value, specifying the column number of the main regressor variable. |

Usage of SexRegress():
SexRegress(fdata, regressor_index, response_index)

Output of SexRegress():

Numeric value containing the regression estimate ("Estimate"), standard error ("Std. Error"), statistics ("t value") and p-value ("Pr(>|t|)")

#### FilterPlinkSample: Making PLINK files with desired samples.

This function prepares PLINK binary files containing user-specified samples

Table 20: Description of the input arguments of FilterPlinkSample() function.

| Arguments | Description |
|---|---|
| DataDir | Character string for the file path of all input files. |
| ResultDir | Ccharacter string for the file path where the output PLINK files will be stored. |
| finput | Character string, specifying the prefix of the input PLINK binary files. |
| foutput | Character string, specifying the prefix of the output PLINK binary files. |
| filter_sample | Character string, specifying the sample type to be retained. The options are: "cases","controls","males" and "females". The default is "cases". |
| keep_remove_sample_file | Character string, specifying the prefix of a space/tab-delimited text file with no header. For the samples that users want to keep or remove, the family IDs should be in the first column and within-family IDs in the second column. This file needs to be in the DataDir. The default is NULL. |
| keep | Boolean value, "TRUE" or "FALSE" for specifying samples to keep or remove. The default is "TRUE". |

Usage of FilterPlinkSample():
FilterPlinkSample(DataDir, ResultDir, finput, foutput = NULL, filter_sample = "cases", keep_remove_sample_file = NULL, keep = TRUE)

Output of FilterPlinkSample():

The output plink files with samples meeting the specification will be saved in ResultDir.

#### ComputeGeneticPC: Computing principal components from genetic relationship matrix.

This function performs principal components analysis (PCA) based on the variance-standardized relationship matrix[3].

Table 21: Description of the input arguments of ComputeGeneticPC() function.

| Arguments | Description |
|---|---|
| finput | Character string, specifying the prefix of the input PLINK binary files. This file needs to be in DataDir. |
| countPC | Integer value, specifying the number of principal components. The default is 10. |
| plotPC | Boolean value, 'TRUE' or 'FALSE', specifying whether to plot the first two PCs. |
| highLD_regions | An R dataframe containing coordinates of genomic regions with high LD for use in finding LD-pruned SNPs in the plots. The default is NULL. |
| ld_prunning | Numeric value between 0 to 1 of pairwise r^2 threshold for LD-based filtering of pruned SNPs in the plots. The default is 0.02. |
| window_size | Integer value, specifying a window size in variant count or kilobases for LD-based filtering. The default is 50. |
| step_size | Integer value, specifying a variant count to shift the window at the end of each step for LD filtering of pruned SNPs in the plots. The default is 5. |

| | |
|---|---|
| r2_threshold | Numeric value between 0 to 1 of pairwise r^2 threshold for LD-based filtering of pruned SNPs in the plots. The default is 0.02. |

Usage of ComputeGeneticPC():
ComputeGeneticPC(DataDir, ResultDir = tempdir(), finput, countPC = 10, plotPC = TRUE, highLD_regions = NULL, ld_prunning = TRUE, window_size = 50, step_size = 5, r2_threshold = 0.02)

Output of ComputeGeneticPC():

A dataframe containing genetic principal components. The first two columns are IID (i.e., Individual Id) and FID (i.e., Family ID). The other columns contain the PCs.

#### ClumpLD: Clumping SNPs using linkage disequilibrium between SNPs.

This function, which is based on empirical estimations of linkage disequilibrium between SNPs, groups the SNP-based results across one or more datasets or analysis. This approach can be used in two basic scenarios: (i) To summarize the top independent associations from a genome-wide association study. (ii) To provide a simple approach to facilitate merging datasets from multiple studies, when those studies may have used different SNP sets for genotyping.

The clumping process begins with the index SNPs that are significant at threshold p1 and have not yet been clumped. It then creates clumps of all additional SNPs that are within a specified kb of the index SNP, and that are in LD with the index SNP (based on a user-specified r-squared threshold). As this method is greedy[3], each SNP will, at most, only appear in one clump. The P value and ALLELES would always, at random, be chosen from the first input file if the same SNP appeared in several input files in SNPdata argument. Instead of the best p-value, the function refers to the SNP that has the strongest LD to the index as the best proxy. Based on the genotype data, the SNP with the highest LD will be the same for all input files.

Table 22: Description of the input arguments of ClumpLD() function.

| **Arguments** | **Description** |
|---|---|
| DataDir | A character string for the file path of the input PLINK binary files. |
| finput | Character string, specifying the prefix of the input PLINK binary files which will be used to calculate linkage disequilibrium between the SNPs. This genotype data may or may not be the same dataset that was used to generate the summary statistics. This file needs to be in DataDir. |
| SNPdata | a list of R dataframes containing a single or multiple summary statistics with SNP and P (i.e., pvalues) in mandatory column headers. Other columns could be present. |
| ResultDir | A character string for the file path where all output files will be stored. The default is tempdir(). |
| clump_p1 | Numeric value, specifying the significance threshold for index SNPs. The default is 0.0001. |
| clump_p2 | Numeric value, specifying the secondary significance threshold for clumped SNPs. The default is 0.01 |
| clump_r2 | Numeric value, specifying the LD threshold for clumping. The default is 0.50. |
| clump_kb | Integer value, specifying the physical distance threshold in base-pair for clumping. The default is 250. |
| byCHR | Boolean value, 'TRUE' or 'FALSE', specifying whether to perform the clumping chromosome-wise. |

Usage of ClumpLD():
ClumpLD(DataDir, finput, SNPdata, ResultDir = tempdir(), clump_p1, clump_p2, clump_r2, clump_kb, byCHR = TRUE, clump_best = TRUE, clump_index_first = TRUE)

Output of ClumpLD():

A list with specifying dataframes. These include *BestClump*: a dataframe with eight columns showing the single best proxy SNP for each index SNP with columns "INDEX"(Index SNP identifier), "PSNP"(Best proxy SNP), "RSQ LD"(r-squared) between index and proxy, "KB"(Physical distance between index and proxy), P(p-value for proxy SNP), "ALLELES"(The associated haplotypes for the index and proxy SNP), and "F"(Which file used for clumping from which this result came from). *AllClump*: a dataframe with eight columns providing a detailed summary of each clump identified by PLINK. It includes "INDEX_SNP" (the identifier for the index SNP that represents the clump), "SNP" (the SNP being reported, which for the index SNP is the same as INDEX_SNP), "DISTANCE" (the physical distance in base pairs between the index SNP and the reported SNP, with 0.0 indicating the index itself), "RSQ" (the r-squared value specifying LD between the index SNP and the SNP in the clump), "ALLELES" (the allele information, which in some cases may appear misaligned if the data isn't formatted as expected), "F" (a statistic or indicator related to the association test, which may be NA when not applicable), "P" (the p-value for the association test of the SNP), and "CHR" (the chromosome on which the SNP is located).

#### GetMFPlink: Getting male and female PLINK binary files.

This function prepares separate male and female PLINK binary files from combined PLINK files.

Table 23: Description of the input arguments of GetMFPlink() function.

| Arguments | Description |
|---|---|
| DataDir | Character string for the file path of the input PLINK binary files. |
| ResultDir | A character string for the file path where all output files will be stored. The default is tempdir(). |
| finput | Character string, specifying the prefix of the input PLINK binary files. |
| foutput | Character string, specifying the prefix of the output PLINK binary files. |
| sex | Boolean value, 'males' or 'females', specifying output plink binary files with male or female samples. |
| xplink | Boolean value, 'TRUE' or 'FALSE', specifying output plink binary files with only X chromosome or not. Default is FALSE. |
| autoplink | Boolean value, 'TRUE' or 'FALSE', specifying output plink binary files with only autosomes or not. Default is FALSE. |

Usage of GetMFPlink():
GetMFPlink(DataDir, ResultDir = tempdir(), finput, foutput, sex, xplink = FALSE, autoplink = FALSE)

Output of GetMFPlink():

Male- and female-specific PLINK files will be stored in the ResultDir.

#### plinkVCF: Converting VCF files to PLINK binary files and vice-versa.

This function performs the conversion between VCF files to PLINK binary formats.

For VCF to PLINK files conversion, if a uses does not specify a FAM file when converting from VCF to PLINK format, then PLINK will create a 'dummy' FAM file with the same name as the dataset, but missing phenotypes and missing sex.

Table 24: Description of the input arguments of plinkVCF() function.

| Arguments | Description |
|---|---|
| DataDir | A character string for the file path of the input PLINK binary files. |
| ResultDir | A character string for the file path where all output files will be stored. The default is tempdir(). |
| finput | Character string, specifying the prefix of the input PLINK binary files or vcf files. This file needs to be in DataDir. |
| foutput | Character string, specifying the prefix of the output PLINK binary files if a filtering option for the SNPs is specified. The default is "FALSE". |
| VtoP | Boolean value, TRUE or FALSE, specifying the conversion of VCF files to PLINK binary files or not. The default is TRUE. |
| PtoV | Boolean value, TRUE or FALSE, specifying the conversion of PLINK binary files to VCF files or not. The default is TRUE. |
| Famfile | Character string, specifying the name of the original .fam file if VtoP was set to be TRUE. This file needs to be in DataDir. The default is NULL |
| PVbyCHR | Boolean value, TRUE or FALSE specifying to do the PLINK to vcf conversion chromosome-wise or not. The default is TRUE. |

Usage of plinkVCF():
plinkVCF(DataDir, ResultDir = tempdir(), finput, foutput, VtoP = FALSE, PtoV = TRUE, Famfile = NULL, PVbyCHR = TRUE)

Output of plinkVCF():
The output files will be saved in ResultDir.

#### MergeRegion: Merging two sets of PLINK binary files.

This function combines two genotype datasets based on either common SNPs (intersection) or all the SNPs between them (union).

Table 25: Description of the input arguments of MergeRegion() function.

| Arguments | Description |
|---|---|
| DataDir | A character string for the file path of the input PLINK binary files. |
| ResultDir | A character string for the file path where all output files will be stored. The default is tempdir(). |
| finput1 | Character string, specifying the prefix of the first input PLINK binary files. |
| finput2 | Character string, specifying the prefix of the first input PLINK binary files. |
| foutput | Character string, specifying the prefix of the output PLINK binary files if filtering option for the SNPs is chosen. The default is "FALSE". |
| use_common_snps | Boolean value, TRUE or FALSE, specifying to use common SNPs for merging or to use all the SNPs. |

Usage of MergeRegion():

MergeRegion(DataDir, ResultDir, finput1, finput2, foutput, use_common_snps = TRUE)

Output of MergeRegion():
The output files will be saved in ResultDir.

#### FilterAllele: Filtering out the multi-allelic variants.

This function filters out multi-allelic SNPs from the input dataset.

Table 26: Description of the input arguments of FilterAllele() function.

| Arguments | Description |
|---|---|
| DataDir | A character string for the file path of the input PLINK binary files. |
| ResultDir | A character string for the file path where all output files will be stored. The default is tempdir(). |
| finput | Character string, specifying the prefix of the input PLINK binary files or vcf files. This file needs to be in DataDir. |
| foutput | Character string, specifying the prefix of the output PLINK binary files. If multi-allelic variants are present, this file will be produced after filtering out these variants |

Usage of FilterAllele():
FilterAllele(DataDir, ResultDir, finput, foutput)
Output of plinkVCF():
The output files will be saved in ResultDir.

#### PlinkSummary: Summary of PLINK format genotype dataset.

This function produces a summary of PLINK format genotype dataset

Table 27: Description of the input arguments of PlinkSummary() function.

| Arguments | Description |
|---|---|
| DataDir | A character string for the file path of the input PLINK binary files. |
| ResultDir | A character string for the file path where all output files will be stored. The default is tempdir(). |
| finput | Character string, specifying the prefix of the input plink binary files or vcf files. This file needs to be in DataDir. |

Usage of PlinkSummary():
PlinkSummary(DataDir, ResultDir, finput)

Output of PlinkSummary():
The output files will be saved in ResultDir.

#### FilterSNP: Filter out SNPs.

This function filters out user-specified SNPs.

Table 28: Description of the input arguments of MergeRegion() function.

| Arguments | Description |
|---|---|

| | |
|---|---|
| DataDir | A character string for the file path of the input PLINK binary files. |
| ResultDir | A character string for the file path where all output files will be stored. The default is tempdir(). |
| finput | Character string, specifying the prefix of the finput PLINK binary files. |
| foutput | Character string, specifying the prefix of the output PLINK binary files if filtering option for the SNPs is chosen. The default is "FALSE". |
| SNPvec | R dataframe with SNP names to be excluded. |
| extract | Boolean value, TRUE or FALSE, specifying whether to extract the SNPs or discard the SNPs. The default is FALSE. |

Usage of MergeRegion():
MergeRegion(DataDir, ResultDir, finput1, finput2, foutput, use_common_snps = TRUE)

Output of MergeRegion():

The output files will be saved in ResultDir.

##### DummyCovar: Function to recode a categorical variable to a set of binary dummy variables.

When working with categorical variables in genetic analysis, a common approach is to convert these into dummy variables[3]. This function creates K-1 new dummy variables for a variable with K categories. One level is automatically excluded from the dummy variables, which serves as the reference category for subsequent analyses. This setup implicitly sets the excluded category as the baseline against which other categories are compared.

Table 29: Description of the input arguments of DummyCovar() function.

| **Arguments** | **Description** |
|---|---|
| DataDir | A character string for the file path of the input PLINK binary files. |
| bfile | Character string, specifying the prefix of the input PLINK binary files for which covariate file will be generated. |
| incovar | Character string, specifying the prefix of the input covariate file. The first two columns are: FID (i.e., Family ID), IID (i.e., Sample ID), the the rest of the columns specifying covariates. |
| outcovar | Character string, specifying the prefix of the Output covariate file |

Usage of DummyCovar():
MergeRegion(DataDir, bfile, incovar, outcovar)

Output of DummyCovar():

R dataframe object with covariates

##### GXWASmiami: Miami plots for GWAS and XWAS.

This function generates Miami plots for GWAS and XWAS.

Table 30: Description of the input arguments of GXWASmiami() function.

| **Arguments** | **Description** |
|---|---|
| DataDir | A character string for the file path of the input PLINK binary files. |

| | |
|---|---|
| FemaleWAS | R dataframe of summary statistics from GWAS or XWAS of female samples with four columns, SNP(Variant), CHR(Chromosome number), POS(basepair position) and pvalue(p-value of the test). This can be generated by running FM01comb or FM02comb model with GXWAS function. |
| MaleWAS | R dataframe of summary statistics of GWAS or XWAS of male samples with four columns, SNP(Variant),CHR(Chromosome number), POS(basepair position) and pvalue(p-value of the test). This can be generated by running the FM01comb or FM02comb model with the GXWAS function. |
| snp_pval | Numeric value corresponding to p-value threshold for annotation. SNPs with p≤p-value threshold will be annotated on the plot. The default is 1e-08. |
| Xchr | Boolean value, TRUE or FALSE, specifying whether to generate Miami plot for stratified XWAS or not. The default is TRUE. |

Usage of GXWASmiami():
GXWASmiami(ResultDir = tempdir(), FemaleWAS, MaleWAS, snp_pval = 1e-08, Xchr = FALSE)

Output of GXWASmiami():

Miami plots.

#### Download_reference: Download Hapmap phase 3 and 1000 Genome phase 3 Data.

Downloads reference data sets from specified URLs based on the reference dataset name and working directory. Currently supports 'HapMapIII_NCBI36' and 'ThousandGenome'.

Table 31: Description of the input arguments of Download_reference() function.

| **Arguments** | **Description** |
|---|---|
| refdata | A character string specifying the reference dataset to download. Should be one of 'HapMapIII_NCBI36' or 'ThousandGenome'. |
| wdir | A character string specifying the working directory where the reference data will be downloaded and extracted. |

Usage of Download_reference():
Download_reference(refdata, wdir = tempdir())

Output of Download_reference():

Invisible. The function prints a message upon successful download and extraction of the reference data.

#### executePlinkMAF: Execute PLINK to Calculate Minor Allele Frequencies (MAF).

This function executes PLINK to calculate minor allele frequencies (MAF) for a given dataset. It sets up the required PLINK environment, runs the PLINK command, and returns the MAF results as a DataFrame. Intermediate files generated by PLINK are deleted after execution.

Table 32: Description of the input arguments of executePlinkMAF() function.

| **Arguments** | **Description** |
|---|---|
| DataDir | A character string for the file path of the input PLINK binary files. |
| ResultDir | A character string for the file path where all output files will be stored. The default is tempdir(). |
| finput | Character string, specifying the prefix of the input PLINK binary files with both male and female samples. This file needs to be in DataDir. |

Usage of executePlinkMAF():
executePlinkMAF(DataDir, ResultDir = tempdir(), finput)

Output of executePlinkMAF():
R dataframe containing the minor allele frequencies (MAFs) for each SNP.

##### LDPrune: Performs LD pruning of genotype data.

This function performs linkage disequilibrium (LD) pruning of genetic data. It identifies and removes SNPs that are in high LD with each other within specified windows.

Table 33: Description of the input arguments of LDPrune() function.

| **Arguments** | **Description** |
|---|---|
| DataDir | A character string for the file path of the input PLINK binary files. |
| ResultDir | A character string for the file path where all output files will be stored. The default is tempdir(). |
| finput | Character string, specifying the prefix of the input PLINK binary files with both male and female samples. This file needs to be in DataDir. |
| window_size | Integer, specifying the number of SNPs to include in the sliding window. |
| step_size | Integer, specifying the number of SNPs the window moves in each step. |
| r2_threshold | Numeric, specifying the $R^2$ threshold for LD pruning. |

Usage of LDPrune():

LDPrune(DataDir, finput, ResultDir = tempdir(), window_size = 50, step_size = 5, r2_threshold = 0.2)

Output of LDPrune():
A character vector of SNP identifiers that remain after LD pruning or NULL if an error occurs.

#### Description of statistical models used in the functions of GXwasR

##### Function GXwas():

The models *FMcomb* and *GWAcxci* are used for running GWAS including XWAS

*FMcomb:* Combining the p-values of the *FMstratified* tests to obtain a final sex-combined GWAS p-values using Fisher method[5], Fisher method with permutation[6] and Stouffer method[7] to allow the SNP tested to have different, even an opposite, effect risk in males and females. This functionality is automated into the *FMstratified* model of *GXwas()* function and recommended to be useful for ChrX variants.

For the fisher.method, the function for combining p-values uses a statistic, $S = -2 \sum_{i=1}^{k} \log(p_i)$ (where k is the number of summary statistics to be combined), which follows a $\chi^2$ distribution with 2k degrees of freedom[5]. For the fisher.method.perm, using p-values from sex-stratified tests, the summary statistic for combining p-values is $S = -2 \Sigma \log(p)$. A p-value for this statistic can be derived by randomly generating summary statistics[6]. Therefore, a p-value is randomly sampled from each contributing study, and a random statistic is calculated. The fraction of random statistics greater or equal to S corresponds to the final p-value.

For the stouffer.method, the function applies Stouffer's method[7] to the p-values assuming that the p-values to be combined are independent. Letting $p_1, p_2, ..., p_k$ denote the individual (one- or two-sided) p-values of the k summary statistics to be combined, the test statistic is then computed with $Z = (1/\sqrt{k}) \Sigma Z_i$ where $Z_i = \Phi^{-1}(1 - p_i)$ and $\Phi^{-1}(\cdot)$ denotes the inverse of the cumulative distribution function of a standard normal distribution. Under the joint null hypothesis, the test statistic follows a standard normal distribution which is used to compute the combined p-value. This functionality is taken from the R package *poolr*[8].

*GWAcxci:* This model represents a female-male combined analysis for autosomes, akin to *FMcombx01* or *FMcombx02*. However, for ChrX, it incorporates a continuous XCI pattern[9], particularly suitable for binary traits. It encompasses coding for complex XCI patterns like XCI-SN (Skewed inactivation towards the Normal allele) and XCI-SR (Skewed inactivation towards the Risk allele). Unlike sex-stratified tests, which cannot incorporate the XCI patterns and may thus lose power if the variant is in a region that undergoes XCI[9], GWAcxci integrates these patterns.

In the context of ChrX, the *GWAcxci* model provides genotypic scores proposed by Su et. al[9] for four different genotype codings, including XCI-Random, XCI-Escape, XCI-SN, and XCI-SR, (Figure 3). The model adjusts the coding for female genotypes to 0, γ, and 2, where γ (ranging between 0 and 2) quantifies the degree of XCI skewness. For example, $\gamma = 0$ implies complete inactivation of risk alleles in heterozygous females, aligning with the XCI-SN pattern, whereas $\gamma = 2$ indicates inactivation of all non-risk alleles, aligning with XCI-SR[9]. Within the GXwasR framework, this model enables autosomal association analysis to include sex as a covariate. However, for ChrX variant analysis, sex is not an external covariate but is intrinsically considered.

All GWAS models can incorporate multiple covariates and their interactions. To facilitate this, GXwasR provides the function "*DummyCovar()*" to generate a new covariate file with categorical variables recoded as binary dummy variables for the covariate file with categorical variables. For example, if a variable has K categories, K-1 new dummy variables are constructed, and the original covariate is now estimated with a coefficient for each category.

These models assume additive allelic effects, and each additional minor allele's influence is represented by the direction of the regression coefficient (i.e., a positive regression coefficient means that the minor allele increases risk i.e., mean of the phenotype). Importantly, the parameter estimates will become unstable in the presence of multi-collinearity, or when the predictor variables are too tightly correlated to one another. The *GXwas()* function handles this situation by displaying NA for the test statistics and a p-value for all terms in the model. Including more terms in the modelincreases the likelihood that these issues will arise.

#### Function SexDiff():

The *SexDiff()* function is applied to summary statistics derived from sex-stratified GWAS and XWAS tests, and uses the following t-statistic to test for a difference in effect size:

$$t - stat = \frac{\log(OR_{male}) - \log(OR_{female})}{\sqrt{SE_{male}^2 - SE_{female}^2 - 2rSE_{male}SE_{female}}}$$

where $OR$ stands for the odds ratio estimated in either the male-only or female-only analysis, $SE$ is the standard error in either test, and $r$ is the Spearman rank correlation coefficient between $\log(OR_{male})$ and $\log(OR_{female})$ across all SNPs. To test for sex differences in effect size for ChrX SNPs, the input dataframes should only include ChrX variants.

#### Function SexDiffZscore():

The *SexDiffZscore()* can be used totest for a difference in multiple different statistical genetic metrics, including SNP-association effect size, SNP heritability etc., using a Z-score and associated p-value. For testing for sex differences in effect size, the Z-score is defined by the following equation:

$$Z_{score} = \frac{\log(OR_{male}) - \log(OR_{female})}{\sqrt{SE_{male}^2 - SE_{female}^2}}$$

A user might choose to define SNPs with sex-differential effect (SDEs) as those variants at the extreme ends of the distribution with an absolute value of the Z-score greater than 3 (|Z-score| > 3), which is roughly equivalent to $p < 10^{-3}$, and represents 0.3% of all tested SNPs (as in [18]).

##### Function DiffZeroOne():

The function, *DiffZeroOne(),* evaluates a Z-score to assess deviations from one and zero, testing the null hypothesis that a statistic (e.g., genetic correlation between males and females) is less than one using a one-tailed test against a normal distribution. The Z-score in this case is calculated as:

$$Z_{score} = \frac{1 - Measured\ Statistic}{Standard\ Error}$$

##### Function EstimateHerit():

GXwasR performs two types of heritability estimation, (i) GREML: Genetic relatedness matrix (GRM) restricted maximum likelihood utilizing GCTA[15] and (ii) LDSC: LD score regression[16,17] using the *EstimateHerit()* function.

The GREML model uses genotype data to estimate the proportion of phenotypic variance explained for a given trait by all of the provided SNPs (genome-wide), or separately for each chromosome. The GREML equation is:

$y = X\beta + g + \varepsilon$, where the variance of y is: $Var(y) = A\sigma_{g^2} + I\sigma_{\varepsilon^2}$

where,
$y = n \times 1$ vector of phenotypes, n = sample size, β = a vector of fixed effects, which may include factors such as sex, age, and/or one or more eigenvectors from principal component analysis,
$g = a\ n \times 1$ vector representing the total genetic effects for the whole genome or for a single chromosome, ε = a vector of residual effects,
$\sigma_{\varepsilon^2}$ = the residual variance, A = the GRM between all individuals, I = the $n \times n$ identity matrix,
$\sigma_{g^2}$ = the variance explained by all the SNPs, i.e., SNP heritability.

Convergence is a known issue in running REML analysis when variance components become very small or negative, such that the V matrix becomes non-positive definite. In this case, GXwasR provides options for various SNP partitioning methods, such as MAF-based partitioning, per chromosome partitioning, etc.

The LDSC model uses GWAS summary statistics and LD scores computed from the genotype data from which the summary statistics are generated. If the corresponding genotype data is not available, HapMap 3 or 1000 Genomes SNPs can be used[17], however, for the most accurate LD scores, the original GWAS genotype data should be used.

Briefly, LDSC is based on the principle that in a single-SNP analysis, the $\chi^2$ test statistic for $SNP_j$ has an expected value represented by:

$$E(\chi_j^2) = 1 + nh_j^2 + n\sum_{k \neq j} r_{j,k}^2 h_k^2 + na_j$$

where:
$r_{j,k}^2$ denotes the squared correlation between $SNP_j$ and $SNP_k$.
$h_j^2$ is the heritability contribution from $SNP_j$.
$a_j$ represents the bias due to confounding factors such as population structure and familial relatedness.

Under a polygenic model, where every SNP contributes equally, and with the assumption that the bias is constant across SNPs (i.e., $a_j = a$), the expected value of $E(\chi_j^2)$ can be expressed as:

$$E(\chi_j^2) = 1 + \frac{n l_j h_{SNP}^2}{m} + na$$

where:

m is the number of SNPs.

$l_j = \sum_{k \neq 1} r_{j,k}^2$ is referred to as the LD score of $SNP_j$.

$\frac{h_{SNP}^2}{m}$ is the average heritability explained per SNP.

Therefore, estimates of $h_{SNP}^2$ and a are computed by regressing test statistics on LD scores. This relationship holds for meta-analyses, and also for studies with binary phenotypes, in which case $h_{SNP}^2$ is on the observed scale[16,17].

#### Function GeneticCorrBT():

The GCTA-based[15] function, *GeneticCorrBT()*,computes genetic correlation, a metric that quantifies the shared genetic architecture underlying two traits or studies and has been predicted to indicate pleiotropic gene activity or correlation between causal loci in two traits, or studies. For example, it performs a bivariate GREML analysis to determine the genetic association between two quantitative traits, two binary disease traits from case-control studies, and between a quantitative trait and a binary disease trait. The function models the residual covariance between the two traits in this analysis. However, the residual covariance will be automatically removed from the model if the traits were measured on separate individuals (e.g., two traits). This function provides can compute the genetic correlation genome-wide or separately for each chromosome. It can also compute genetic correlation using a variety of different SNP partitioning methods in case of a convergence issue in running the GREML model.

#### Function MetaGwas():

The *MetaGwas()* function combines summary statistics from GWASes of the same or similar phenotype(s). This function uses PLINK's inverse variance-based approach to enable fixed-effect and random-effect models, and quantifies effect size heterogeneity using Cochran's Q[19] and the $I^2$ statistics[20]. This function also calculates weighted Z-score-based p-values using METAL[12]. The function can also apply correction for genomic inflation factor[21].

In the random-effect model, the effect size of the reference allele at the j-th SNP in the i-th study is denoted as $\beta_{ij}$. The combined allelic effect across all studies at the j-th SNP is calculated as:

$$\beta_{ij} = \frac{\sum_{i=1}^{N} \beta_{ij} w_{ij}}{\sum_{i=1}^{N} w_{ij}},$$

where $w_{ij} = \frac{1}{Var(\beta_{ij})}$ represents the inverse of the variance of the estimated allelic effect in the i-th study, derived from the standard error. In cases where the j-th SNP has not been directly genotyped or imputed in the i-th study, $w_{ij}$ is set to zero.

The corresponding statistic, $X_j^2 = \frac{\beta_j^2}{V_j}$, follows an approximate chi-square distribution with one degree of freedom, providing the basis for testing the association of the trait with the j-th SNP across all studies.

To address the deflation in the variance of the fixed effects estimate when heterogeneity in allelic effects across studies is present, random-effects meta-analysis is often employed[22]. In this model, it is assumed that each study inflates the variance of the predicted allelic effect by incorporating the random-effects variance component at the j-th SNP, denoted as $\tau_j^2$. The cumulative allelic effect across all studies at the SNP is then recalculated as:

$$\beta^*_{ij} = \frac{\sum_{i=1}^{N} \beta_{ij} w^*_{ij}}{\sum_{i=1}^{N} w_{ij}},$$

where $w_{ij} = \frac{1}{\tau_j^2 + \mathrm{Var}(\beta^*_{ij})}$.

The variance of this combined allelic effect across the studies, represented as $V^*_{ij}$, is calculated as:

$$V^*_{ij} = \left(\sum_{i=1}^{N} w^*_{ij}\right)^{-1}.$$

Furthermore, the statistic $X_j^2$, which is defined as $X_j^2 = \frac{\beta_j^2}{V_j}$, follows an approximate chi-square distribution with one degree of freedom. This statistic is used to account for study heterogeneity in the association of the trait with the j-th SNP.

METAL's Z-score: A reference allele is selected for each SNP, and a z-statistic is computed to quantify the strength and direction of the effect relative to that allele. The overall z-statistic and p-value are obtained using a weighted total of individual statistics, with weights proportional to the square root of the number of individuals in each sample.

When population structure is present in a GWAS, it can lead to over-dispersion of the test statistics[23]. To address this, the genomic control inflation factor, denoted as $\lambda_i$, is used. This factor is calculated by dividing the median of the test statistics by 0.456, the expected median under the null hypothesis[24]. The genomic control inflation factor adjusts the test statistics of each study to control for potential population structure. This is achieved by increasing the variance associated with each SNP in the analysis, resulting in a new weight calculation:

$$w_{ij} = \frac{1}{\lambda_i \times \mathrm{Var}(\beta_{ij})}.$$

Furthermore, to correct for between-study variation in the meta-analysis, the test statistic $X_j^2$ is adjusted as follows:

$$X_j^2 = \frac{\beta_j^2}{\lambda_i V_j}.$$

To quantify the consistency of allelic effects across studies at the same SNP, the *MetaGwas()* function computes two metrics of heterogeneity: the Cochran's Q[19] and the $I^2$ statistics[20]. Cochran's Q tests the heterogeneity of allelic effects at the j-th SNP and follows an approximate $\chi^2$ distribution with $N_{j-1}$ degrees of freedom under the null hypothesis of consistency, where $N_j$ is the number of studies for which an allelic effect has been recorded. The $I^2$ statistic, more resistant to variation in the number of studies than Q, quantifies the degree of heterogeneity in allelic effects across studies.

#### Function TestXGene():

The *TestXGene()* function enables gene-based association testing, utilizing GWAS/XWAS summary statistics and SNP-SNP correlation matrices using either user-provided genotype data or 1000 Genomes Phase 3 reference data. The function is able to perform a variety of gene-based tests, including the burden test (BT[25]), sequence kernel association test (SKAT[26]), SKAT-O[27] (a combination of BT and SKAT), sum of $\chi^2$-statistics (sumchi), aggregated Cauchy association test (ACAT[28]), principal component approach (PCA[29]), functional multiple linear regression model (FLM[30]), Bonferroni correction test (simpleM[10]), and minimum P-value (minp[10]). These tests are executed by leveraging the functionality of PLINK1.9 and sumFREGAT[10] tools.

For the BT, SKAT, and SKATO methods, *TestXGene()* conducts gene-level association tests using random-effects models. These models require z-scores, the SNP-SNP correlation matrix, and two diagonal matrices, V and W:

*Matrix* W*:* Contains user-defined weights for genetic variants, which can be determined using the beta distribution density function:

$$W_i = \frac{MAF_i^{a-1} \times (1 - MAF_i^{b-1})}{B(a,b)}.$$

Here, $MAF_i$ represents the minor allelic frequency for the i-th genetic variant. The parameters a and b are the shape parameters of the beta distribution, and B(a, b) is the beta function, which is a normalization constant ensuring that the total probability integrates to 1.

*Matrix* V*:* Holds the genotypic variances, calculated using the standard errors of genetic effects.

In contrast, the PCA and FLM approaches employ regression models with fixed effects, establishing an orthogonal basis set to reduce the number of predictors in the regression model. Users could use 1000 Genome-based correlation matrices in the absence of study genotype data. If so, this function provides the option to adjust the correlation matrices to prevent the false-positive outcomes of the fixed-effects model-based approaches using Tikhonov regularization[31], which was implemented in sumFREGAT.

Each gene-based test within *TestXGene()* is uniquely characterized:

*SKAT and SKATO:* These methods employ a linear weighted kernel function but differ slightly in their approach to setting the inter-individual similarity matrix[10].

*ACAT:* This test computes gene-based statistics from P values (Z scores) alone, without needing SNP-SNP correlation information[10].

*PCA and simpleM:* Based on the spectral decomposition of the SNP-SNP correlation matrix, it selects the top principal components that explain a specified fraction of the region variance[10].

A similar principle is used in 'simpleM' to calculate the effective number of independent tests[10].

*FLM:* Assumes the effects of multiple genetic variants can be described as a continuous function. When the number of basis functions (set by k) is less than the number of variants (m) within the region, the FLM test has the advantage of using fewer degrees of freedom. For genes with fewer variants than the number of basis functions (m <= k), it equates to a standard multiple linear regression. For this test, the "model" column will be returned in the output as an R dataframe object[10].

#### Tutorial for running the QC pipeline on pre-imputed genotype data using GXwasR:

The PLINK bed, bim, and fam files are the three mandatory files representing the test genotype dataset to run this pipeline. These file extensions are described at https://www.cog-genomics.org/plink/1.9/formats. GXwasR_example.bed; GXwasR_example.bim and GXwasR_example.fam PLINK files contain genotypes for 276 individuals (males and females) simulated from 1000Genomes data of individuals of European ancestry with 26,515 variants across twelve chromosomes (1-10,23,24). This dataset contains 125 males, 151 females, 108 cases and 168 controls.

##### Pre-imputation QC steps

**Filtering Multi-Allelic Variants:** This step removes variants that have more than two alleles.

**Removal of Ambiguous SNPs and Indels:** We remove SNPs with ambiguous mappings (like AT and GC pairings) and indels (insertions and deletions.

**QC of SNPs with Relaxed Genotype Call Rate:** We remove SNPs based on a less stringent genotype call rate to balance between retaining informative SNPs and excluding those with too much missing data.

**Filtering Samples with Relaxed Missing Rate Threshold:** Similar to SNPs, we apply a relaxed threshold for sample missingness to make sure that we don't prematurely exclude samples that might be informative despite having a slightly higher rate of missing data.

**Application of Sex Check:** We verify the consistency between reported and genetic sex. Inconsistencies can indicate sample mix-ups or issues with data integrity.

**Filter for Needed Chromosomes:** This step streamlines the dataset to the chromosomes of interest.

**Second Round of SNP Filtering:** This includes: filtering for minor allele frequency (MAF) < 0.01, call rate < 0.02, case-control differential missingness, Hardy-Weinberg equilibrium (cases < e-10; conrtols < e-06), and the removal of monomorphic SNPs.

**Second Round of Sample Filtering:** This includes: stringent thresholds for call rate (< 0.02), heterozygosity, and relatedness (IBD F-stat < 0.2).

**Applying Ancestry Check:** We assess the ancestry of the samples to identify and exclude outliers from different ancestries.

**Third Round of SNP Filtering:** This final filtering step ensures that all previously applied SNP thresholds are maintained even after the rigorous sample filtering. It's a crucial step to guarantee the overall quality and consistency of the SNP data set.

Loading the GXwasR library

```
## Call some libraries

Library(GXwasR)

## Load the example .Rda files

data("GXwasRData")
```

#### Example Dataset Summary

##### Dataset: GXwasR_example

```
DataDir <- system.file("extdata", package = "GXwasR")
ResultDir <- tempdir()
finput <- "GXwasR_example"
x <- PlinkSummary(DataDir, ResultDir, finput)
```

```
## [1] "Program is set up."
## [1] "Dataset:GXwasR_example"
## [1] "This is a case-control data."
## [1] "Number of males:125"
## [1] "Number of females:151"
## [1] "Number of missing phenotypes:0"
## [1] "Number of chromosomes:12"
##  [1] "Chr:1"  "Chr:2"  "Chr:3"  "Chr:4"  "Chr:5"  "Chr:6"  "Chr:7"  "Chr:8"
##  [9] "Chr:9"  "Chr:10" "Chr:23" "Chr:24"

## [1] "Total number of SNPs:26527"
## [1] "Total number of samples:276"
```

##### Filtering multi-allelic variants

First, users need to ensure that their input PLINK files contain only bi-allelic variants. To do so, run the FilterAllele() function with the input dataset.

```
foutput <- "PreimputeEX_QC1"
x <- FilterAllele(DataDir, ResultDir, finput, foutput)
```

```
## [1] "Program is set up."
## [1] "There is no multi-allelic SNP present in the input dataset."
```

Since there there are multi-allelic SNPs in the dataset, users can continue to the next step using the same input data.

##### QC of SNPs for genotype call rate

This step removes variants that are missing genotype data for more than 20% of the individuals in the dataset. This function also implicitly removes ambiguous SNPs (i.e., AT<>GC), and indels.

```
# Running
foutput <- "PreimputeEX_QC1"
geno <- 0.2
maf <- NULL
casecontrol <- FALSE
caldiffmiss <- FALSE
diffmissFilter <- FALSE
dmissX <- FALSE
dmissAutoY <- FALSE
monomorphicSNPs <- TRUE
ld_prunning <- FALSE
casecontrol <- FALSE
hweCase <- NULL
hweControl <- NULL
monomorphicSNPs <- FALSE ld_prunning <-
FALSE

x <- QCsnp(DataDir = DataDir, ResultDir = ResultDir, finput = finput,foutput = foutput, geno = geno, maf = maf,hweCase = hweCase,
           hweControl = hweControl, ld_prunning = ld_prunning, casecontrol = casecontrol, monomorphicSNPs = monomorphicSNPs,
           caldiffmiss = caldiffmiss, dmissX = dmissX, dmissAutoY = dmissAutoY, diffmissFilter = diffmissFilter)
```

```
## [1] "Program is set up."
## [1] "4214 Ambiguous SNPs (A-T/G-C), indels etc. were removed."
## [1] "Thresholds for maf, geno and hwe worked."
## [1] "11 variants removed due to missing genotype data (--geno)."
## [1] "No filter based on differential missingness will be applied."
## [1] "Output plink files prefixed as ,PreimputeEX_QC1, with passed SNPs are saved in ResultDir."
## [1] "Input file has 26527 SNPs."
## [1] "Output file has 22302 SNPs after filtering."
```

Copying the plink files from ResultDir to DataDir.

```r
ftemp <- list.files(paste0(ResultDir,"/"),pattern = "PreimputeEX_QC1")
file.copy(paste0(ResultDir,"/",ftemp),DataDir)
```

```
## [1] FALSE FALSE FALSE FALSE
```

```r
PlinkSummary(DataDir, ResultDir, finput = "PreimputeEX_QC1")
```

```
## [1] "Program is set up."
## [1] "Dataset:PreimputeEX_QC1"
## [1] "This is a case-control data."
## [1] "Number of males:125"
## [1] "Number of females:151"
## [1] "Number of missing phenotypes:0"
## [1] "Number of chromosomes:11"
##  [1] "Chr:1"  "Chr:2"  "Chr:3"  "Chr:4"  "Chr:5"  "Chr:6"  "Chr:7"  "Chr:8"
##  [9] "Chr:9"  "Chr:10" "Chr:23"

## [1] "Total number of SNPs:22302"
## [1] "Total number of samples:276"
```

##### Filtering samples with a high missing rate threshold.

This step excludes samples that are missing genotype data for more than 20% of the genetic variants in the dataset.

```r
# Running
finput <- "PreimputeEX_QC1"
foutput <- "PreimputeEX_QC2"
imiss = 0.2
het = NULL
IBD = NULL
x = QCsample(DataDir = DataDir,ResultDir = ResultDir, finput = finput,foutput = foutput, imiss =
         imiss,het = het, IBD = NULL)
```

```
## [1] "Program is set up."
## [1] "No. of ambiguous samples filtered out: 0"
## [1] "Plots are initiated."
```

```
## [1] "No samples filtered for missingness."
## [1] "No samples filtered for heterozygosity."
## [1] "No samples filtered for missingness and heterozygosity."
```

```
## An error occurred: object 'failed_ibd' not found
```

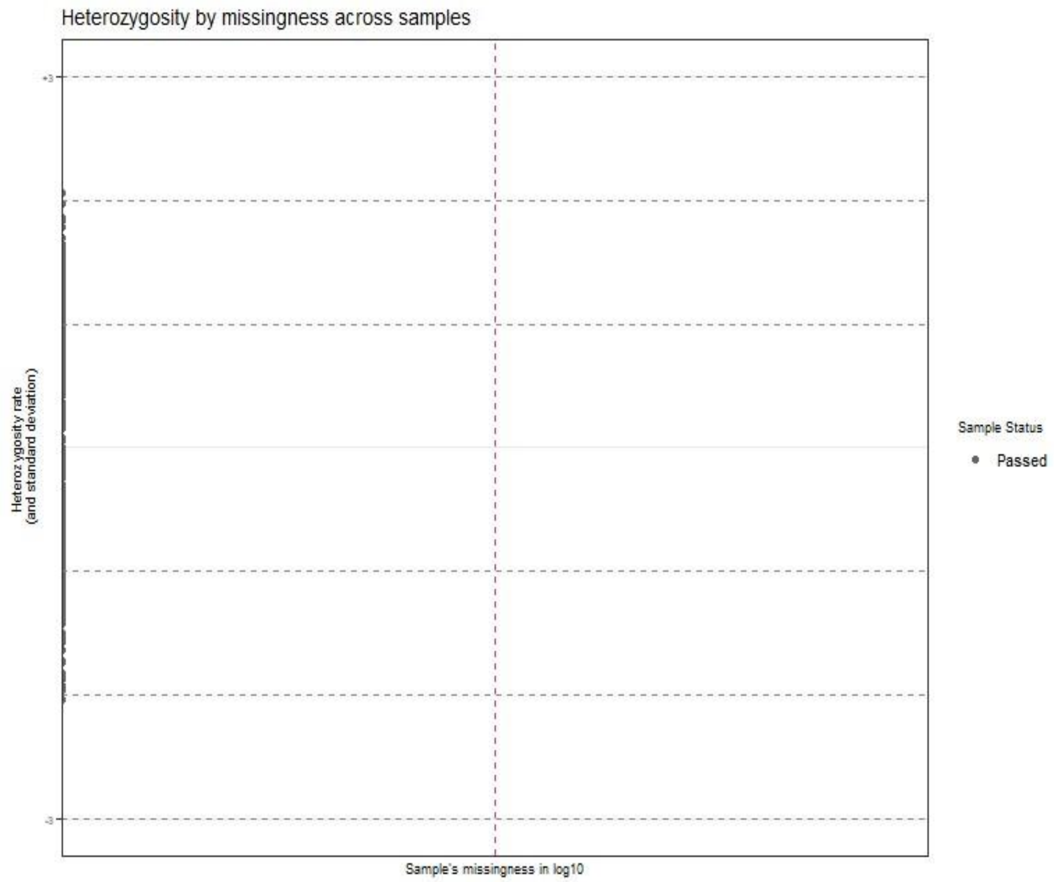

Figure1 : An example of heterozygosity by missingness across samples. Here all samples are passed the test.

#### Apply sex check

```r
library(GXwasR)
finput <- "PreimputeEX_QC1" # Using the same input file
LD = TRUE
```

```r
LD_window_size <- 50
LD_step_size <- 5
LD_r2_threshold <- 0.02
fmax_F <- 0.3
mmin_F <- 0.7
# We will not impute sex
impute_sex <- FALSE
compute_freq <- FALSE
SexCheckResult <-SexCheck(DataDir=DataDir,ResultDir = ResultDir,
         finput=finput,impute_sex=impute_sex,compute_freq
         =compute_freq,LD_window_size=LD_window_size,LD_step_size=LD_step_size,LD_r2_threshold=0.02,fmax_F
         = fmax_F,mmin_F=mmin_F)
```

```
## [1] "Program is set up."
```

```
## Warning: There are no Y chromosomes in the input PLINK files. Estimates will be based solely on the
X chromosome.
```

```r
problematic_sex <- SexCheckResult[SexCheckResult$STATUS != "OK",]
problematic_sex <- problematic_sex[,1:2]
print(paste0("Number of samples with problematic sex assignment: ",nrow(problematic_sex)))
```

```
## [1] "Number of samples with problematic sex assignment: "
```

```r
write.table(problematic_sex,file = paste0(DataDir,"/problematic_sex_samples"),quote = F, row.names =
         F,col.names = F)
SexCheckResult[1:5,]
```

```
## NULL
```

There is no filtering of the samples based on a sex mismatch.

#### Filter for needed chromosomes.

```
test1 <- read.table(paste0(DataDir,"/",finput,".bim"))
unique(test1$V1)
```

```
##  [1]  1  2  3  4  5  6  7  8  9 10 23
```

```
foutput <- "PreimputeEX_QC2"
x <- FilterRegion(DataDir = DataDir, ResultDir = ResultDir, finput = finput, foutput = foutput, CHRX =
         FALSE, CHRY = FALSE, filterPAR = FALSE, filterXTR = FALSE, filterAmpliconic = FALSE,
         regionfile = FALSE, filterCHR = c(24, 25, 26),Hg = "38", exclude = TRUE)
```

```
## [1] "Program is set up."
## [1] "0 SNPs are discarded."
## [1] "Plink files with passed SNPs are in C:\\Users\\Sarah\\AppData\\Local\\Temp\\Rtmpe0NA0W prefixed
as PreimputeEX_QC2"
## [1] "Plink files with discarded SNPs are in C:\\Users\\Sarah\\AppData\\Local\\Temp\\Rtmpe0NA0W
prefixed as PreimputeEX_QC2_snps_extracted"
```

Copying the plink files from ResultDir to DataDir.

```
ftemp <- list.files(paste0(ResultDir,"/"),pattern = "PreimputeEX_QC2")
file.copy(paste0(ResultDir,"/",ftemp),DataDir)
```

```
## [1] FALSE FALSE FALSE FALSE FALSE
```

```
PlinkSummary(DataDir, ResultDir, finput = "PreimputeEX_QC2")
```

```
## [1] "Program is set up."
## [1] "Dataset:PreimputeEX_QC2"
## [1] "This is a case-control data."
## [1] "Number of males:125"
## [1] "Number of females:151"
## [1] "Number of missing phenotypes:0"
## [1] "Number of chromosomes:11"
##  [1] "Chr:1"  "Chr:2"  "Chr:3"  "Chr:4"  "Chr:5"  "Chr:6"  "Chr:7"  "Chr:8"
##  [9] "Chr:9"  "Chr:10" "Chr:23"
## [1] "Total number of SNPs:22302"
## [1] "Total number of samples:276"
```

```
## NULL
```

#### Second round of SNP filtering.

```
library(GXwasR)
finput <- "PreimputeEX_QC2"
foutput <- "PreimputeEX_QC3"
geno <- 0.02
maf <- 0.01
casecontrol <- TRUE
caldiffmiss <- TRUE
diffmissFilter <- TRUE
dmissX <- TRUE
dmissAutoY <- TRUE
monomorphicSNPs <- TRUE
ld_prunning <- FALSE
hwe = NULL
hweCase <- 1e-10
hweControl <- 1e-06
x <- QCsnp(DataDir = DataDir, ResultDir = ResultDir, finput = finput,foutput = foutput, geno = geno,
         maf = maf,hweCase = hweCase, hweControl = hweControl, ld_prunning = ld_prunning, casecontrol =
         casecontrol, monomorphicSNPs = monomorphicSNPs, caldiffmiss = caldiffmiss, dmissX = dmissX,
         dmissAutoY = dmissAutoY, diffmissFilter = diffmissFilter)
```

```
## [1] "Program is set up."
## [1] "0 Ambiguous SNPs (A-T/G-C), indels etc. were removed."
## [1] "Thresholds for maf, geno and hwe worked."
## [1] "0 variants removed due to missing genotype data (--geno)."
## [2] "1930 variants removed due to minor allele threshold(s)"
## [1] "In cases,  1 variant removed due to Hardy-Weinberg exact test."
## [1] "In controls,  19 variants removed due to Hardy-Weinberg exact test."
## [1] "Program is set up."
```

#### [1] "Merging is done using the common SNPs between the input genotype files." ## [1] "Plink files with merged regions are in C:\\Users\\Sarah\\AppData\\Local\\Temp\\Rtmpe0NA0W prefixed as filtered_temp2" ## [1] "There are no monomorphic SNPs." ## [1] "No SNP with differential missingness between cases and controls."

#### [1] "Output plink files prefixed as ,PreimputeEX_QC3, with passed SNPs are saved in ResultDir."

#### [1] "Input file has 22302 SNPs."

#### [1] "Output file has 20353 SNPs after filtering."

```
ftemp <- list.files(paste0(ResultDir,"/"),pattern = "PreimputeEX_QC3")
file.copy(paste0(ResultDir,"/",ftemp),DataDir)
```

```
## [1] FALSE FALSE FALSE FALSE
```

```
PlinkSummary(DataDir, ResultDir, finput = "PreimputeEX_QC3")
```

```
## [1] "Program is set up."
## [1] "Dataset:PreimputeEX_QC3"
## [1] "This is a case-control data."
## [1] "Number of males:125"
## [1] "Number of females:151"
## [1] "Number of missing phenotypes:0"
## [1] "Number of chromosomes:11"
##  [1] "Chr:1"  "Chr:2"  "Chr:3"  "Chr:4"  "Chr:5"  "Chr:6"  "Chr:7"  "Chr:8"
##  [9] "Chr:9"  "Chr:10" "Chr:23"
## [1] "Total number of SNPs:20353"
## [1] "Total number of samples:276"
```

```
## NULL
```

#### Second round of sample filtering

Here we apply more stringent thresholds for the sample filter.

```
finput <- "PreimputeEX_QC3"
foutput <- "PreimputeEX_QC4"
imiss = 0.02
het = 3
IBD = 0.2
IBDmatrix = FALSE
small_sample_mod = TRUE
filterSample = TRUE
x = QCsample(DataDir = DataDir,ResultDir = ResultDir, finput = finput,foutput = foutput, imiss =
         imiss,het = het, IBD = IBD, IBDmatrix = IBDmatrix)
```

```
## [1] "Program is set up."
## [1] "No. of ambiguous samples filtered out: 0"
## [1] "Plots are initiated."
```

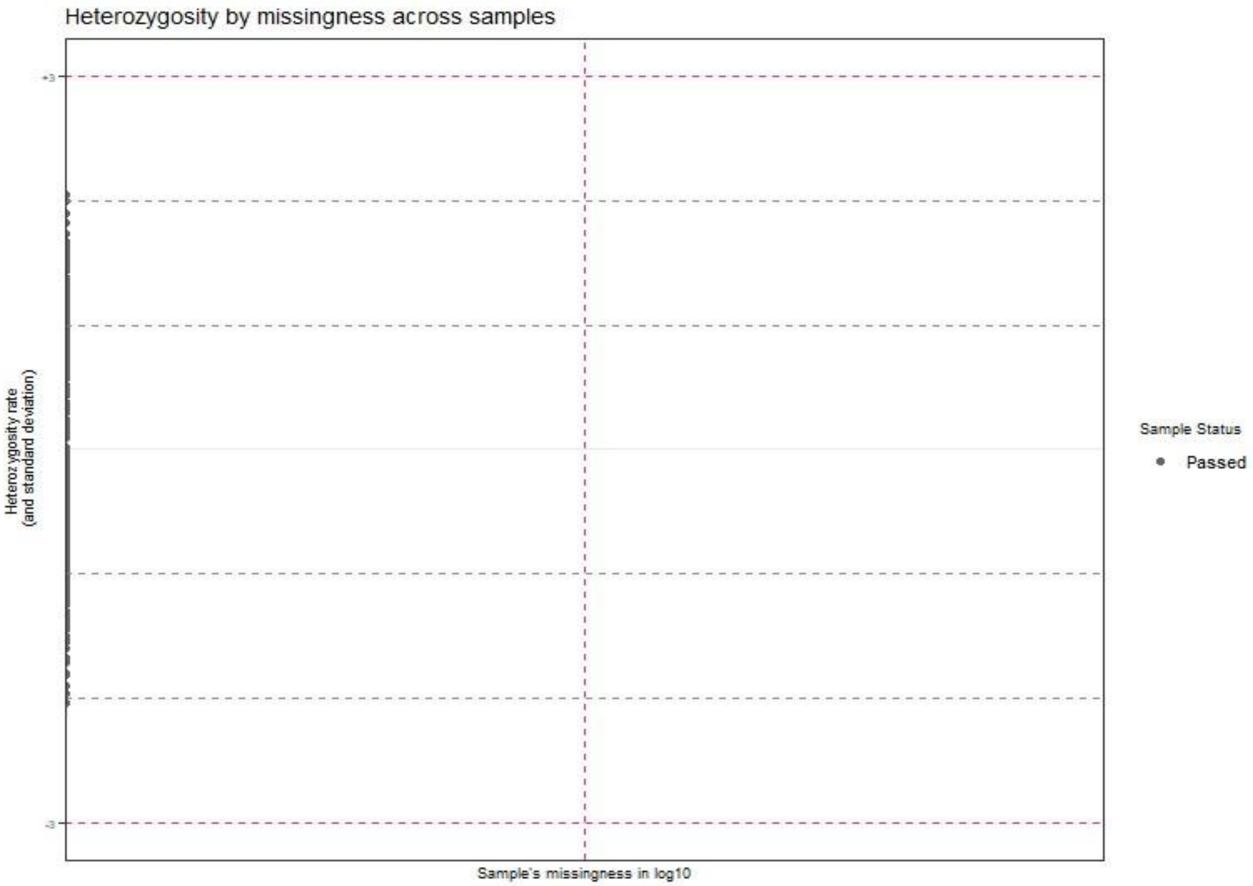

Figure 2 : An example of heterozygosity by missingness across samples after second round of filtering.

```
## [1] "No samples filtered for missingness."
## [1] "No samples filtered for heterozygosity."
## [1] "No samples filtered for missingness and heterozygosity."
## [1] "1 samples are filtered out for IDB after missingness and heterozygosity filter."
## [1] "No. of samples in input plink files: 276"
## [1] "No. of samples in output plink files: 274"
```

```
## [1] "Output plink files, PreimputeEX_QC4 with final samples are in
C:\\Users\\AppData\\Local\\Temp\\Rtmpe0NA0W."
```

Copying the plink files from ResultDir to DataDir.

```
ftemp <- list.files(paste0(ResultDir,"/"),pattern = "PreimputeEX_QC4")
file.copy(paste0(ResultDir,"/",ftemp),DataDir)
```

```
## [1] FALSE FALSE FALSE FALSE
```

```
PlinkSummary(DataDir, ResultDir, finput = "PreimputeEX_QC4")
```

```
## [1] "Program is set up."
## [1] "Dataset:PreimputeEX_QC4"
## [1] "This is a case-control data."
## [1] "Number of males:124"
## [1] "Number of females:150"
## [1] "Number of missing phenotypes:0"
## [1] "Number of chromosomes:11"
##  [1] "Chr:1"  "Chr:2"  "Chr:3"  "Chr:4"  "Chr:5"  "Chr:6"  "Chr:7"  "Chr:8"
##  [9] "Chr:9"  "Chr:10" "Chr:23"
## [1] "Total number of SNPs:20353"
## [1] "Total number of samples:274"
```

```
## NULL
```

#### Ancestry check

```
finput <- "PreimputeEX_QC4"
reference <- "HapMapIII_NCBI36"
highLD_regions <- highLD_hg19
study_pop <- example_data_study_sample_ancestry
studyLD_window_size = 50
```

```
studyLD_step_size = 5
studyLD_r2_threshold = 0.02
filterSNP = TRUE
studyLD = TRUE
referLD = TRUE
referLD_window_size = 50
referLD_step_size = 5
referLD_r2_threshold = 0.02
outlier = TRUE
outlier_threshold = 3
outlierOf = "CEU"
x <- AncestryCheck(DataDir = DataDir, ResultDir = ResultDir, finput = finput, reference =
         reference,highLD_regions = highLD_regions,study_pop = study_pop, studyLD = studyLD, referLD =
         referLD, outlier = outlier, outlier_threshold = outlier_threshold )
```

```
## [1] "Program is set up."
## [1] "Reference data 'HapMapIII_NCBI36' downloaded and extracted in
C:\\Users\\Sarah\\AppData\\Local\\Temp\\Rtmpe0NA0W."
## [1] "No SNP had 'A-T' and 'G-C' in study data."
## [1] "111854 SNPs were 'A-T' and 'G-C' in reference data. These SNPs were removed."
## [1] "LD pruning was done for study dataset."
## [1] "LD pruning was done for reference dataset."
## [1] "PCA done."
```

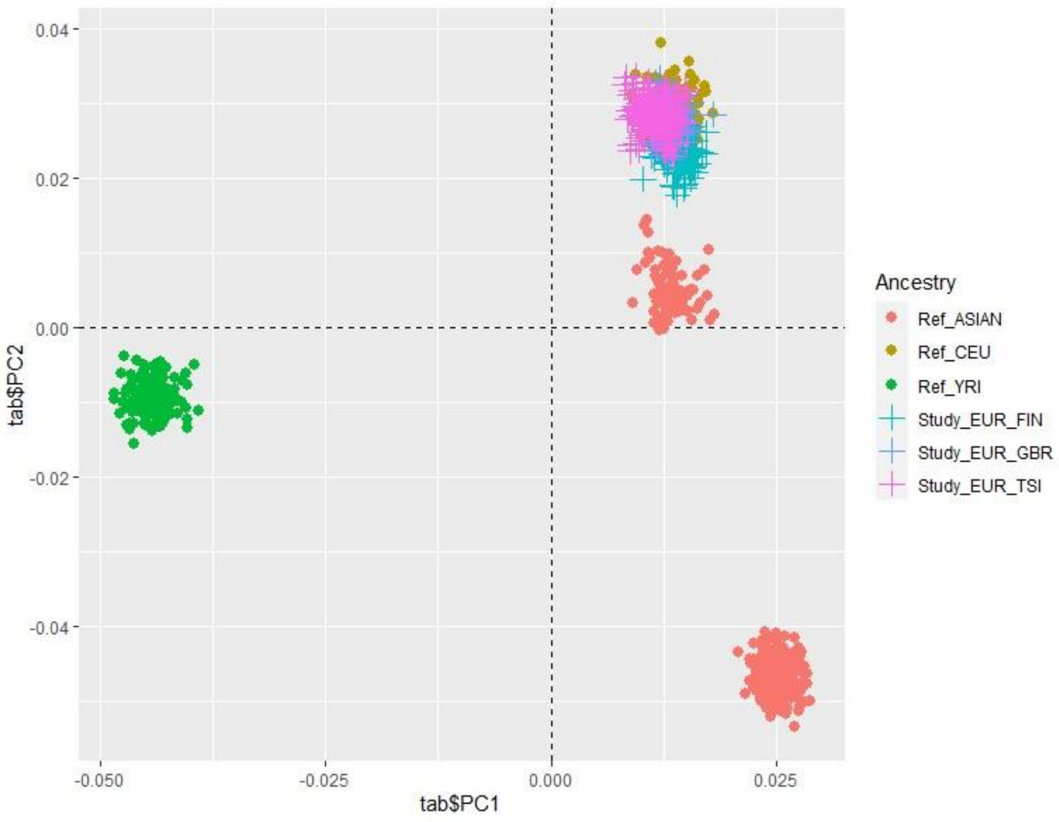

Figure 3: PCA plot showing the distribution of individual genetic samples in the quality-controlled (QC-ed) genotype dataset along the first two principal components (PC1 and PC2), based on HapMapIII reference populations. Each point represents a genetic sample, color-coded by ancestry group. Reference populations

include: Ref_ASIAN = East Asian ancestry (e.g., Han Chinese, Japanese); Ref_CEU = Utah residents with Northern and Western European ancestry; Ref_YRI = Yoruba in Ibadan, Nigeria. Study samples are labeled by sub-population: Study_EUR_FIN = Finnish; Study_EUR_GBR = British; Study_EUR_TSI = Toscani in Italy.
.

```
## [1] "406 allele flips identified between study and reference
## [1] "3448 SNPs were finally retained in study and reference data after correcting for
mismatch and allele
## [1] "There is no non-European sample as outlier sample.
```

There was no outlier sample.

##### Third round of SNP filtering

To make sure all the filtering thresholds for SNPs are maintained after sample filtering.

```
library(GXwasR)
finput <- "PreimputeEX_QC4"
foutput <- "Preimpute_Final"
geno <- 0.02
maf <- 0.01
casecontrol <- TRUE
caldiffmiss <- TRUE
diffmissFilter <- TRUE
dmissX <- TRUE
dmissAutoY <- TRUE
monomorphicSNPs <- TRUE
ld_prunning <- FALSE
 hwe = NULL
hweCase <- 1e-10
hweControl <- 1e-06
x <- QCsnp(DataDir = ResultDir, ResultDir = ResultDir, finput = finput,foutput = foutput, geno = geno,
         maf = maf,hweCase = hweCase, hweControl = hweControl, ld_prunning = ld_prunning, casecontrol =
         casecontrol, monomorphicSNPs = monomorphicSNPs, caldiffmiss = caldiffmiss, dmissX = dmissX,
         dmissAutoY = dmissAutoY, diffmissFilter = diffmissFilter)
```

```
## [1] "Program is set up."
## [1] "0 Ambiguous SNPs (A-T/G-C), indels etc. were removed."
## [1] "Thresholds for maf, geno and hwe worked."
## [1] "0 variants removed due to missing genotype data (--geno)."
## [2] "3 variants removed due to minor allele threshold(s)"
## [1] "In cases,  0 variants removed due to Hardy-Weinberg exact test."
## [1] "In controls,  0 variants removed due to Hardy-Weinberg exact test."
## [1] "Program is set up."
## [1] "Merging is done using the common SNPs between the input genotype files."
## [1] "Plink files with merged regions are in C:\\Users\\Sarah\\AppData\\Local\\Temp\\Rtmpe0NA0W
prefixed as filtered_temp2"
## [1] "There are no monomorphic SNPs."
## [1] "No SNP with differential missingness between cases and controls."
## [1] "Output plink files prefixed as ,Preimpute_Final, with passed SNPs are saved in ResultDir."
## [1] "Input file has 20353 SNPs."
## [1] "Output file has 20350 SNPs after filtering."
```

##### Summary of final QC-ed dataset.

```
PlinkSummary(ResultDir,ResultDir,foutput)
```

```
## [1] "Program is set up."
## [1] "Dataset:Preimpute_Final"
## [1] "This is a case-control data."
## [1] "Number of males:124"
## [1] "Number of females:150"
## [1] "Number of missing phenotypes:0"
## [1] "Number of chromosomes:11"
##  [1] "Chr:1"  "Chr:2"  "Chr:3"  "Chr:4"  "Chr:5"  "Chr:6"  "Chr:7"  "Chr:8"
##  [9] "Chr:9"  "Chr:10" "Chr:23"
## [1] "Total number of SNPs:20350"
## [1] "Total number of samples:274"
```

```
## NULL
```

#### Decoding Ancestry: A guide to using GXwasR for genetic ancestry estimation

GXwasR estimates sample ancestry relevant to reference populations through the use of the AncestryCheck function.
The core of ancestry estimation lies in combining the genotypes of a study population with those from a reference dataset, such as the HapMap or 1000 Genomes studies. This integration allows for the detection of population structure down to the level of the reference dataset, revealing continental and subcontinental genetic structure.

##### Accessing GXwasR and Simulated Genotype Datasets

GXwasR is available on the Comprehensive R Archive Network (CRAN) and can be installed using install.packages("GXwasR"). The package includes simulated genotype dataset prefixed as **GXwasR_example** as plink bed files (i.e., .bed, .fam and .bim)[3], for users to experiment with. This simulated dataset consists of genotypes from 276 individuals across 26,527 genetic markers, providing a simplified dataset for testing functionality.

To see details of the sample genotype dataset 'Study data':

```
## Call GXwasR
library(GXwasR)
DataDir <- system.file("extdata", package = "GXwasR")
ResultDir <- tempdir()
```

```r
finput <- "GXwasR_example"
x <- PlinkSummary(DataDir,ResultDir, finput)
```

```
## [1] "Program is set up."
## [1] "Dataset:GXwasR_example"
## [1] "This is a case-control data."
## [1] "Number of males:125"
## [1] "Number of females:151"
## [1] "Number of missing phenotypes:0"
## [1] "Number of chromosomes:12"
##  [1] "Chr:1"  "Chr:2"  "Chr:3"  "Chr:4"  "Chr:5"  "Chr:6"  "Chr:7"  "Chr:8"
##  [9] "Chr:9"  "Chr:10" "Chr:23" "Chr:24"
## [1] "Total number of SNPs:26527"
## [1] "Total number of samples:276"
```

##### AncestryCheck Function Overview

AncestryCheck is a function to assess genetic ancestry. By leveraging PLINK via R, it integrates several steps: SNP filtering for data integrity, linkage disequilibrium (LD) pruning to reduce data complexity, correction of chromosome number and SNP position mismatches, and checking for allele flips. Additionally, it merges study and reference datasets, performs Principal Component Analysis (PCA) to detect population structure, and incorporates outlier detection and visualization. This functioncan compare study samples' ancestry labels against those of reference populations, enabling analysis of population structure.

##### Workflow for ancestry estimation in AncestryCheck

##### Data Preparation:

**Directories and initial quality check:** The AncestryCheck function requires specific directories for inputting PLINK binary files and for saving output files. At this stage, the input genotype dataset, i.e., the study dataset, should have already undergone quality control (see Section Description of GXwasR functions).

**Genomic Build and X-Chromosome Compatibility:** To use AncestryCheck , the study and reference datasets must use the same genomic build, with consistent chromosome coding across datasets. This alignment is crucial, particularly since sex chromosomes (X and Y) often have inconsistent naming conventions across datasets. However, users are advised to use tools like Liftover[32] or CrossMap[33] for verifying and aligning genomic builds and chromosome naming between their study and reference datasets
**Reference Dataset Selection:** Users can select from two established reference populations, including **HapMap phase III**[1] or the **1000 Genomes phase 3**[2] Projects. It is specifically designed to work seamlessly with the preprocessed genotypes of these reference panels.

##### SNP Filtering:

The function incorporates SNP filtering for both study and reference datasets to exclude SNPs that do not conform to A-T or G-C base pairings.

##### LD Pruning:

The function includes LD-based filtering for both the study and reference datasets to remove variants in high LD. This ensures accurate PCA of the dataset SNPs. The default setting targets variants in LD with an $r2$ value greater than 0.2 within a 50kb window. However, AncestryCheck offers users the flexibility to adjust these parameters or exclude LD-pruning altogether. This adaptability ensures compatability with a variety of study designs and research objectives

The AncestryCheck function then uses the list of LD-pruned variants to filter the reference dataset. This step is crucial for accurate comparative analysis between reference and study datasets, and ensures consistency in the ancestral estimation process.

##### Filter out regions of known high-LD structure:

AncestryCheck includes an option to exclude SNPs located in regions of high LD, which is particularly useful for analyses using genomic builds hg19 and hg38. The GXwasR package provides built-in files with the genomic coordinates of these high-LD regions, which are automatically used by AncestryCheck when this option is enabled. Alternatively, users can supply their own high-LD region coordinates as an R data frame object..

##### Genotype Data Processing for Merge:

The function prepares genotype data for merging study and reference datasets:

##### Correcting Mismatches:

The function corrects mismatches in chromosome IDs, positions, and allele annotations between study and reference datasets. This step is essential to ensure that the study and reference data align accurately. By rectifying any discrepancies, AncestryCheck safeguards against potential errors that could arise from inconsistencies between the datasets.

##### Handling Allele Flips:

Allele flips are a frequent complication in genotype data, where alleles may be reported in reverse order across datasets. The function can identify and rectify these flips.

##### Handling Sex Chromosomes:

The Encoding of sex chromosomes often differs from that of autosomal chromosomes, which can introduce additional complexity when matching study and reference datasets. For example, the X chromosome may be labeled as ChrX, X, Chr23, or 23 depending on the source. AncestryCheck is designed to recognize and harmonize these variations to ensure accurate alignment across datasets.

##### Removing Mismatches Post-Flipping:

After the allele flipping process, the function performs another check for any remaining mismatches. Any alleles that are still not aligned correctly after the flipping process are identified and removed from the reference dataset. This final step ensures that only matched SNPs are retained for the merged dataset.

##### Principal Component Analysis (PCA):

**Merging Datasets:** After preprocessing, the study and reference datasets are merged.

**Performing PCA:** PCA is then performed on the combined genotype dataset to detect and explore dimensions of genetic variation in study and reference datasets.

##### Outlier Detection and Analysis:

**Flexibility in Reference Population Selection:** The function is designed to accommodate multiple reference populations, allowing users to select a specific group against which the study samples are compared. For example, setting the parameter *outlierOF* to CEU (Ref_CEU: Utah residents with Northern and Western European ancestry) will identify samples that are genetically distant from the CEU reference group. Other reference populations, such as YRI (Ref_YRI: Yoruba in Ibadan, Nigeria) and ASIAN (Ref_ASIAN: individuals of East Asian ancestry, such as Han Chinese and Japanese).

##### Setting Outlier Thresholds:
The function allows users to specify thresholds for outlier detection. For each sample, it calculates the Euclidean distance from the center of the chosen reference population's principal components (PC). Samples exceeding the user-defined threshold are flagged as outliers. For example, in the case of CEU reference samples, the function computes the median of the first two principal components (median(PC1 of Ref_CEU), median(PC2 of Ref_CEU)) to determine the center. It then calculates the maximum Euclidean distance (maxDist) from this center. Any study sample whose Euclidean distance from the center equals or exceeds the radius r= outlier threshold * maxDist is considered an outlier. This approach can identify samples that significantly deviate from the selected reference population, providing quantifiable method to identify outliers.

##### Visualization and Reporting:
**PCA Plot Visualization:** AncestryCheck provides a visual representation of the summarized PCA results in the form of a color-coded scatter plot displaying sample and reference individuals in principal component space (i.e., the first two genotype principal components).

**Resultant Dataframe:** Following PCA and outlier analysis, AncestryCheck generates a dataframe containing identifiers for the outlier samples.

**Reports:** In addition to the visual and tabular outputs, the function also produces reports on allele flips and a thorough breakdown of the outlier analysis.

#### AncestryCheck Usage
The function can be used in the following manner with our study data and reference data as HapMapIII:

```
# Define parameters
DataDir <- system.file("extdata", package = "GXwasR")
ResultDir <- tempdir()
finput <- "GXwasR_example"
reference <- "HapMapIII_NCBI36"
data("GXwasRData")
highLD_regions <- highLD_hg19
study_pop <- example_data_study_sample_ancestry
studyLD_window_size = 50
studyLD_step_size = 5
studyLD_r2_threshold = 0.02
filterSNP = TRUE
studyLD = TRUE
referLD = TRUE
referLD_window_size = 50
referLD_step_size = 5
referLD_r2_threshold = 0.02
outlier = TRUE
outlier_threshold = 3

# Call the AncestryCheck function

ancestry_results <- AncestryCheck(
    DataDir = DataDir,
    ResultDir = ResultDir,
    finput = finput,
    reference = reference,
```

```
    filterSNP = TRUE,
    studyLD = TRUE,
    studyLD_window_size = 50,
    studyLD_step_size = 5,
    studyLD_r2_threshold = 0.02,
    referLD = FALSE,
    referLD_window_size = 50,
    referLD_step_size = 5,
    referLD_r2_threshold = 0.02,
    highLD_regions = highLD_regions,
    study_pop = study_pop,
    outlier = TRUE,
    outlierOf = "CEU",
    outlier_threshold = 3
)
```

```
## [1] "Program is set up."
## [1] "Reference data 'HapMapIII_NCBI36' downloaded and extracted in
C:\\Users\\Sarah\\AppData\\Local\\Temp\\RtmpozCJp7."
## [1] "4214 SNPs had 'A-T' and 'G-C' in study data. These SNPs were removed."
## [1] "111854 SNPs were 'A-T' and 'G-C' in reference data. These SNPs were removed."
## [1] "LD pruning was done for study dataset."
## [1] "LD pruning for reference dataset is recommended. Set referLD == TRUE."
## [1] "PCA done."
```

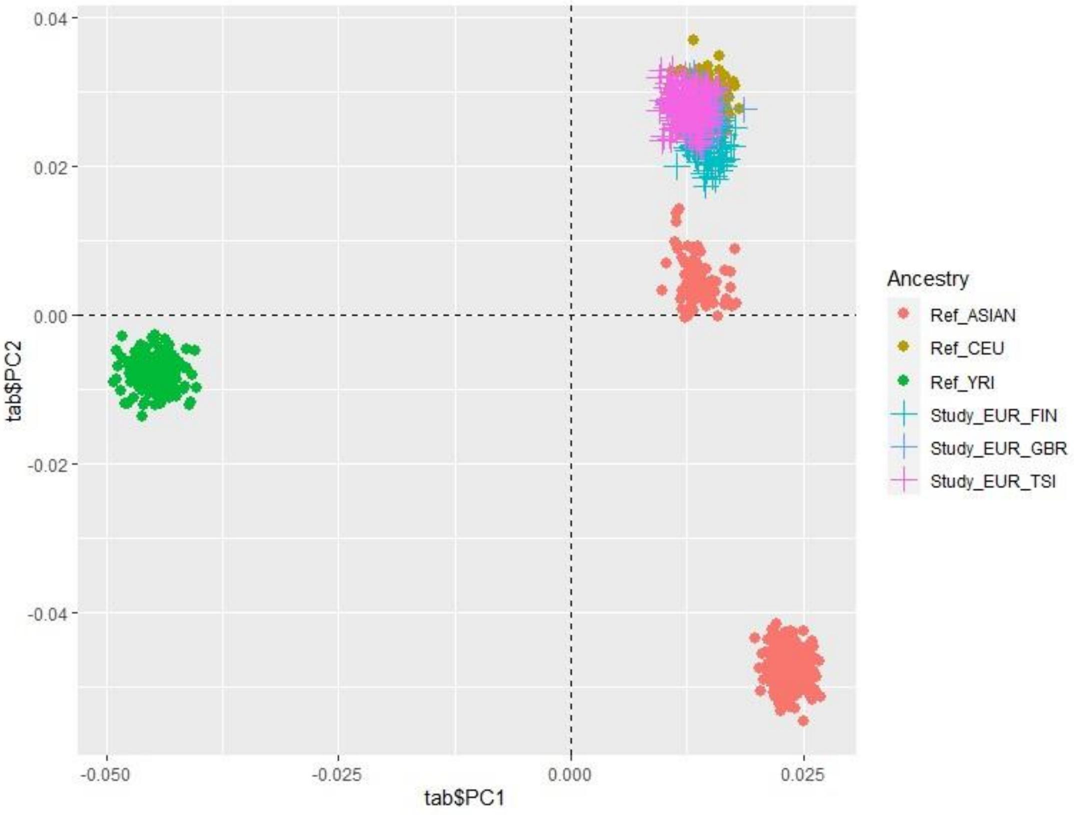

Figure 4: PCA plot showing the distribution of individual  samples in the study dataset (list the pops here) and reference datasets, along the first two principal components (PC1 and PC2). Each point represents a an individual, color-coded based on study and reference populations.

```
## [1] "401 allele flips identified between study and reference data."
## [1] "3619 SNPs were finally retained in study and reference data after correcting for position mismatch and allele flips."
## [1] "There is no non-European sample as outlier."
```

The plot shows the distribution of individual samples along the first two principal components (PC1 and PC2). Each point represents a sample, and they are color-coded based on reference populations (Ref_ASIAN, Ref_CEU, and Ref_YRI) and study populations (Study_EUR_FIN, Study_EUR_GBR, and Study_EUR_TSI).

The points are spread across the PCA plot, with each cluster representing genetic similarity. The reference populations, Ref_ASIAN, Ref_CEU, and Ref_YRI cluster separately, revealing genetic differentiation between these populations. The study populations overlap with the Ref_CEU population, indicating European ancestry, with each study subgroup (FIN, GBR, TSI) forming subclusters, suggesting finer-scale differences in ancestry within Europe.

The lack of outlier samples, those significantly distanced from the reference group, suggests relative genetic homogeneity between the study data and the reference data. Given that our study data were simulated based on CEU 1000 Genomes project data, the absence of outliers is expected.

### Tutorial: Running post-imputation QC using GXwasR

#### Post-imputation QC steps

**Sex-combined QC:** Remove SNPs with IMPUTE2 info score < 0.6 and certainty < 0.8. This step should be done on the imputation server with the VCF files.

**Sex-specific autosomal QC at the variant level:** 1) Filter PAR, XTR, and Ampliconic regions from the X chromosome, and combine these with autosomes for autosomal QC; 2) Any SNPs in the controls with a sex difference in MAF should be carefully examined and flagged for further examination, since the differences may be due to technical confounding or sampling biases for the study cohorts; 3) Prepare separate male and female subsets of the genotypes; 4) Remove SNPs with MAF < 0.05 from each sex.

**Sex-specific X-chromosome SNP QC:** 1) Separate PAR, XTR, Ampliconic regions from the X-chromosome; 2) Prepare separate male and female subsets of the genotypes; 3) Remove SNPs with a missingness difference in cases vs controls, separately in each sex; 4) In each sex, filter out SNPs based on 1% MAF, 10% missingness per individual, and 5% missingness per genotype. We recommend setting the threshold filters to be the same in males and females for analyses of common variants: 5) Identify the set of SNPs found in both of the post-QC male and female datasets and create a combined dataset with only these SNPs; 6) Test for HWE across the X chromosome in females (cases and controls combined), and later remove these SNPs from analysis in all samples. If assuming equal allele frequency between males and females, HWE for chrX can be considered in females only as described Khramtsova et. al.[4]; 7) Test for significantly different MAF ($p < 0.05$/No. of SNPs) between sexes in control samples only for binary traits. ChrX variants with MAF sex differences in controls might still be biologically meaningful, so users might consider flagging these variants rather than removing them[4].

**Sex-specific sample QC:** 1) Remove any individual with missing genotype rate > 0.2) Remove any individual with the absolute value of the heterozygosity F statistic > 0.20; 3) Remove any pair of individuals with IBD statistic pihat > 0.2.

##### Post-imputation QC pipeline

We will use PLINK binary files from DataDir, prefixed as “GXwasR_example” for the input of the post-imputation QC functions. These variants have all previously passed IMPUTE2 info score < 0.6 and certainty < 0.8 filters.

###### Filtering Ambiguous SNPs and indels

Use QCsnp() to filter ambiguous SNPs and indels.

```
finput <- "GXwasR_example"
foutput <- "PostimputeEX_QC1"
geno <- NULL
maf <- NULL
casecontrol <- FALSE
hweCase <- NULL
hweControl <- NULL
hweCase <- NULL
monomorphicSNPs <- FALSE
caldiffmiss <- FALSE
ld_prunning <- FALSE
x <- QCsnp(DataDir = DataDir, ResultDir = ResultDir, finput = finput,foutput = foutput, geno = geno,
         maf = maf,hweCase = hweCase, hweControl = hweControl, ld_prunning = ld_prunning, casecontrol =
         casecontrol, monomorphicSNPs = monomorphicSNPs, caldiffmiss = caldiffmiss)
```

```
## [1] "Program is set up."
## [1] "4214 Ambiguous SNPs (A-T/G-C), indels etc. were removed."
## [1] "Thresholds for maf, geno and hwe worked."
## character(0)
## [1] "There are no monomorphic SNPs."
## [1] "No filter based on differential missingness will be applied."
## [1] "Output plink files prefixed as ,PostimputeEX_QC1, with passed SNPs are saved in ResultDir."

## [1] "Input file has 26527 SNPs."
## [1] "Output file has 22313 SNPs after filtering."
```

Copying the plink files from ResultDir to DataDir.

```
ftemp <- list.files(paste0(ResultDir,"/"),pattern = "PostimputeEX_QC1")
file.copy(paste0(ResultDir,"/",ftemp),DataDir)
```

```
## [1] TRUE TRUE TRUE TRUE
```

###### Filter PAR, XTR and Ampliconic Regions

Now, we will filter the Pseudo-Autosomal Region (PAR), X-transposed region (XTR), and Ampliconic regions from the X-chromosome and will combine these regions with autosomes for autosomal QC.

```r
# Running
finput <- "PostimputeEX_QC1"
x <- PlinkSummary(DataDir, ResultDir, finput)
```

```
## [1] "Program is set up."
## [1] "Dataset:PostimputeEX_QC1"
## [1] "This is a case-control data."
## [1] "Number of males:125"
## [1] "Number of females:151"
## [1] "Number of missing phenotypes:0"
## [1] "Number of chromosomes:12"
##  [1] "Chr:1"  "Chr:2"  "Chr:3"  "Chr:4"  "Chr:5"  "Chr:6"  "Chr:7"  "Chr:8"
##  [9] "Chr:9"  "Chr:10" "Chr:23" "Chr:24"
## [1] "Total number of SNPs:22313"
## [1] "Total number of samples:276"
```

```r
foutput <- "PostimputeEX_QC2"
```

```
x <- FilterRegion(DataDir = DataDir,ResultDir = ResultDir,finput = finput,foutput = foutput,CHRX =
         TRUE,CHRY = FALSE, filterPAR = TRUE,filterXTR = TRUE,filterAmpliconic = TRUE,regionfile =
         FALSE,filterCHR = NULL, Hg = "38",exclude = TRUE)
```

```
## [1] "Program is set up."
## [1] "line 3491"
## [1] "chrX"
## [1] "There is no PAR region in the input data. Argument filterPAR cannot set to be TRUE."
## [1] "Changing it as filterPAR = FALSE"
## [1] "XTR SNPs:37"
## [1] "Ampliconic SNPs:8"
## [1] "45 SNPs are discarded."
## [1] "Plink files with passed SNPs are in C:\\Users\\Sarah\\AppData\\Local\\Temp\\RtmpEtlLko prefixed
as PostimputeEX_QC2"
## [1] "Plink files with discarded SNPs are in C:\\Users\\Sarah\\AppData\\Local\\Temp\\RtmpEtlLko
prefixed as PostimputeEX_QC2_snps_extracted"
```

```
XTR_SNPs <- x$XTR
Ampli_SNPs <- x$Ampliconic
PAR_SNPs <- x$PAR
save(XTR_SNPs, file = paste0(ResultDir,"/Postimpute_XTR_SNPs.Rda"))
save(Ampli_SNPs, file = paste0(ResultDir,"/Postimpute_Ampli_SNPs.Rda"))
save(PAR_SNPs, file = paste0(ResultDir,"/Postimpute_PAR_SNPs.Rda"))
```

```
# Output
# SNPs in PAR
PAR_SNPs <- x$PAR ## No PAR SNP
# SNPs in XTR
XTR_SNPs <- x$XTR ## 42 SNPs
knitr::kable(XTR_SNPs, caption = 'SNPs in XTR.')
```

```
# Get SNPs in Ampliconic region
Ampliconic_SNPs <- x$Ampliconic # 7 SNPs
knitr::kable(Ampliconic_SNPs, caption = 'SNPs in Ampliconic region.')
```

Now, users need to move the QC-ed files (i.e., “PostimputeEX_QC2” and “PostimputeEX_QC2_snps_extracted”) to DataDir to be used as input datasets for the next step of the pipeline.

Copying the plink files from ResultDir to DataDir.

```
ftemp <- list.files(paste0(ResultDir,"/"),pattern = "PostimputeEX_QC2")
file.copy(paste0(ResultDir,"/",ftemp),DataDir)
```

```
## [1] TRUE TRUE TRUE TRUE TRUE TRUE TRUE TRUE
```

##### Preparing autosomal and X-chromosomal plink binary files

Now, we will prepare two sets of PLINK binary files from "PostimputeEX_QC2", containing autosomal SNPs and chromosome X SNPs, respectively. We use FilterRegion().

```
finput <- "PostimputeEX_QC2"
foutput <- "PostimputeEX_QC3"

y <- FilterRegion(DataDir = DataDir,ResultDir = ResultDir,finput = finput,foutput = foutput,CHRX =
        FALSE,CHRY = FALSE, filterPAR = FALSE,filterXTR = FALSE,filterAmpliconic = FALSE,regionfile =
        FALSE,filterCHR = 23, Hg = "19",exclude = TRUE)
```

```
## [1] "Program is set up."
## [1] "938 SNPs are discarded."
## [1] "Plink files with passed SNPs are in C:\\Users\\Sarah\\AppData\\Local\\Temp\\RtmpEtlLko prefixed
as PostimputeEX_QC3"
## [1] "Plink files with discarded SNPs are in C:\\Users\\Sarah\\AppData\\Local\\Temp\\RtmpEtlLko
prefixed as PostimputeEX_QC3_snps_extracted"
```

"PostimputeEX_QC3_snps_extracted" contains SNPs from the X chromosome, while "PostimputeEX_QC3" contains autosomal SNPs.

```
#Now, copying these files to DataDir.
ftemp <- list.files(paste0(ResultDir,"/"),pattern = "PostimputeEX_QC3")
invisible(file.copy(paste0(ResultDir,"/",ftemp),DataDir))
```

##### Merging the autosomal regions with PAR, XTR and Ampliconic regions

We now merge the autosomal files with PAR, XTR, and Ampliconic regions using MergeRegion().

```
finput1 <- "PostimputeEX_QC3"
finput2 <- "PostimputeEX_QC2_snps_extracted"
foutput <- "PostimputeEX_QC3_autopxa"
y <- MergeRegion(DataDir, ResultDir, finput1, finput2, foutput, use_common_snps = FALSE)
```

```
## [1] "Program is set up."
## [1] "Merging is done with all the SNPs i.e., union of the SNPs."
## [1] "Plink files with merged regions are in C:\\Users\\Sarah\\AppData\\Local\\Temp\\RtmpEtlLko
prefixed as PostimputeEX_QC3_autopxa"
```

```
## Copying the files to DataDir
ftemp <- list.files(paste0(ResultDir,"/"),pattern = "PostimputeEX_QC3_autopxa")
invisible(file.copy(paste0(ResultDir,"/",ftemp),DataDir))
```

With "PostimputeEX_QC3_autopxa", we will flag the SNPs in the controls with a sex difference in MAF, since the differences may be due to technical confounding or sampling biases for the study cohorts.

##### Flag the SNPs in the controls with a sex difference in MAF

```
finput <- "PostimputeEX_QC3_autopxa"
foutput <- "Test_output"
x <- MAFdiffSexControl(DataDir, ResultDir, finput,foutput, filterSNP = FALSE)
```

```
## [1] "Program is set up."
## [1] "No SNP to be flagged or excluded."
```

Since no SNP in the test dataset has a MAF difference in sexes in controls, we proceed to the next steps.

##### Prepare separate male and female subsets of the genotypes

```
finput <- "PostimputeEX_QC3_autopxa"
foutput <- "PostimputeEX_QC4_Female"
sex <- "females"
x <- GetMFPlink(DataDir = DataDir, ResultDir = ResultDir, finput = finput, foutput = foutput,sex = sex,
         xplink = FALSE, autoplink = FALSE)
```

```
## [1] "Program is set up."
## [1] "Output plink files, prefixed as PostimputeEX_QC4_Female, are in
C:\\Users\\Sarah\\AppData\\Local\\Temp\\RtmpEtlLko"
```

```
# Making male plink files
foutput <- "PostimputeEX_QC4_Male"
sex <- "males"
x <- GetMFPlink(DataDir = DataDir, ResultDir = ResultDir, finput = finput, foutput = foutput,sex = sex,
         xplink = FALSE, autoplink = FALSE)
```

```
## [1] "Program is set up."
## [1] "Output plink files, prefixed as PostimputeEX_QC4_Male, are in
C:\\Users\\Sarah\\AppData\\Local\\Temp\\RtmpEtlLko"
```

Remove the previous QC-ed file from DataDir and copy the new QC-ed file to DataDir.

```
## Removing the previous QC-ed file from DataDir and copying the new QC-ed file to DataDir.
ftemp <- list.files(paste0(DataDir,"/"),pattern = "PostimputeEX_QC1")
invisible(file.remove(paste0(DataDir,"/",ftemp)))

## Removing the previous QC-ed file from DataDir and copying the new QC-ed file to DataDir.
ftemp <- list.files(paste0(DataDir,"/"),pattern = "PostimputeEX_QC2")
invisible(file.remove(paste0(DataDir,"/",ftemp)))

## Copying new plink files to DataDir

ftemp <- list.files(paste0(ResultDir,"/"),pattern = "PostimputeEX_QC4")
invisible(file.copy(paste0(ResultDir,"/",ftemp),DataDir))
```

**Remove SNPs with MAF < 0.05 from each sex from the sex-specific autosomal PLINK files**

Here, we perform QC of SNPs using MAF, call rate, monomorphic status, and differential missingness in cases vs controls from each sex from the sex-specific autosomal datasets.

We recommend setting the threshold filters to be the same in males and females for analyses of common variants.

For this, we will use QCsnp() function.

```
## Applying filter to female-specific plink files
finput <- "PostimputeEX_QC4_Female"
foutput <- "PostimputeEX_QC5_Female"
geno <- 0.05
maf <- 0.05
casecontrol <- TRUE
hweCase <- NULL
hweControl <- NULL
hweCase <- NULL
monomorphicSNPs <- TRUE
caldiffmiss <- TRUE
ld_prunning <- FALSE
x <- QCsnp(DataDir = DataDir, ResultDir = ResultDir, finput = finput,foutput = foutput, geno = geno,
         maf = maf,hweCase = hweCase, hweControl = hweControl, ld_prunning = ld_prunning, casecontrol =
         casecontrol, monomorphicSNPs = monomorphicSNPs, caldiffmiss = caldiffmiss)
```

```
## [1] "Program is set up."
## [1] "0 Ambiguous SNPs (A-T/G-C), indels etc. were removed."
## [1] "Thresholds for maf, geno and hwe worked."
## [1] "11 variants removed due to missing genotype data (--geno)."
## [2] "5378 variants removed due to minor allele threshold(s)"
## [1] "In cases, "
## [1] "In controls, "
```

```
## [1] "Program is set up."
## [1] "Merging is done using the common SNPs between the input genotype files."
## [1] "Plink files with merged regions are in C:\\Users\\Sarah\\AppData\\Local\\Temp\\RtmpEtlLko
prefixed as filtered_temp2"
## [1] "There are no monomorphic SNPs."
## [1] "Filtering for differential missingness between cases and controls is turned off."
## [1] "No SNP with differential missingness between cases and controls."
## [1] "Output plink files prefixed as ,PostimputeEX_QC5_Female, with passed SNPs are saved in
ResultDir."
## [1] "Input file has 21375 SNPs."
## [1] "Output file has 15986 SNPs after filtering."
```

```
# Applying filter to male-specific plink files
finput <- "PostimputeEX_QC4_Male"
foutput <- "PostimputeEX_QC5_Male"
x <- QCsnp(DataDir = DataDir, ResultDir = ResultDir, finput = finput,foutput = foutput, geno = geno,
         maf = maf,hweCase = hweCase, hweControl = hweControl, ld_prunning = ld_prunning, casecontrol =
         casecontrol, monomorphicSNPs = monomorphicSNPs, caldiffmiss = caldiffmiss)
```

```
## [1] "Program is set up."
## [1] "0 Ambiguous SNPs (A-T/G-C), indels etc. were removed."
## [1] "Thresholds for maf, geno and hwe worked."
## [1] "11 variants removed due to missing genotype data (--geno)."
## [2] "5323 variants removed due to minor allele threshold(s)"
## [1] "In cases, "
## [1] "In controls, "
## [1] "Program is set up."
## [1] "Merging is done using the common SNPs between the input genotype files."
## [1] "Plink files with merged regions are in C:\\Users\\Sarah\\AppData\\Local\\Temp\\RtmpEtlLko
prefixed as filtered_temp2"
## [1] "There are no monomorphic SNPs."
## [1] "Filtering for differential missingness between cases and controls is turned off."
## [1] "No SNP with differential missingness between cases and controls."
## [1] "Output plink files prefixed as ,PostimputeEX_QC5_Male, with passed SNPs are saved in
ResultDir."
## [1] "Input file has 21375 SNPs."
## [1] "Output file has 16041 SNPs after filtering."
```

Remove the previous QC-ed file from DataDir and copy the new QC-ed file to DataDir.

```
ftemp <- list.files(paste0(DataDir,"/"),pattern = "PostimputeEX_QC4")
invisible(file.remove(paste0(DataDir,"/",ftemp)))
ftemp <- list.files(paste0(ResultDir,"/"),pattern = "PostimputeEX_QC4")
invisible(file.remove(paste0(ResultDir,"/",ftemp)))

## Copying the new QC-ed file to DataDir.
ftemp <- list.files(paste0(ResultDir,"/"),pattern = "PostimputeEX_QC5")
invisible(file.copy(paste0(ResultDir,"/",ftemp),DataDir))
```

##### Combine sex-specific autosomal QC-ed files using common SNPs:

```
finput1 <- "PostimputeEX_QC5_Female"
finput2 <- "PostimputeEX_QC5_Male"
foutput <- "PostimputeEX_Auto"
y <- MergeRegion(DataDir, ResultDir, finput1, finput2, foutput, use_common_snps = TRUE)
```

```
## [1] "Program is set up."
## [1] "Merging is done using the common SNPs between the input genotype files."
## [1] "Plink files with merged regions are in C:\\Users\\Sarah\\AppData\\Local\\Temp\\RtmpEtlLko
prefixed as PostimputeEX_Auto"
```

Remove the previous QC-ed file from DataDir and copythe new QC-ed file to DataDir.

```
ftemp <- list.files(paste0(DataDir,"/"),pattern = "PostimputeEX_QC5")
invisible(file.remove(paste0(DataDir,"/",ftemp)))

ftemp <- list.files(paste0(ResultDir,"/"),pattern = "PostimputeEX_Auto")
invisible(file.copy(paste0(ResultDir,"/",ftemp),DataDir))

ftemp <- list.files(paste0(ResultDir,"/"),pattern = "PostimputeEX_QC5")
invisible(file.remove(paste0(ResultDir,"/",ftemp)))

ftemp <- list.files(paste0(ResultDir,"/"),pattern = "PostimputeEX_Auto")
invisible(file.remove(paste0(ResultDir,"/",ftemp)))
```

##### Obtain sex-specific X chromosomal PLINK files

We start with PLINK files prefixed as "PostimputeEX_QC3_snps_extracted". Note: these PLINK files contain X chromosome variants without PAR, XTR, and Ampliconic sites.

We use the GetMFPlink() function to make male and female PLINK binary files for X chromosome variants. We will move these files from ResultDir to DataDir, and then run the sex-specific QC steps.

```r
finput <- "PostimputeEX_QC3_snps_extracted"
foutput <- "PostimputeEX_QC3_XFemale"
# Making female-specific plink files X chromosome
sex <- "females"
x <- GetMFPlink(DataDir = DataDir, ResultDir = ResultDir, finput = finput, foutput = foutput,sex = sex,
         xplink = FALSE, autoplink = FALSE)
```

```
## [1] "Program is set up."
## [1] "Output plink files, prefixed as PostimputeEX_QC3_XFemale, are in
C:\\Users\\Sarah\\AppData\\Local\\Temp\\RtmpEtlLko"
```

```r
# Making male-specific plink files X chromosome
foutput <- "PostimputeEX_QC3_XMale"
sex <- "males"
x <- GetMFPlink(DataDir = DataDir, ResultDir = ResultDir, finput = finput, foutput = foutput,sex = sex,
         xplink = FALSE, autoplink = FALSE)
```

```
## [1] "Program is set up."
## [1] "Output plink files, prefixed as PostimputeEX_QC3_XMale, are in
C:\\Users\\Sarah\\AppData\\Local\\Temp\\RtmpEtlLko"
```

```r
## Copying sex-specific plink files
ftemp <- list.files(paste0(ResultDir,"/"),pattern = "PostimputeEX_QC3_X")

invisible(file.copy(paste0(ResultDir,"/",ftemp),DataDir))
```

##### QC of sex-specific X-chromosomal SNPs

Here, we remove SNPs with a missingness difference in cases vs controls, separately in each sex, and will filter out SNPs based on ≤ 1% MAF, ≥ 5% missingness per genotype, and monomorphic SNPs. We recommend setting the threshold filters to be the same in males and females for analyses of common variants.

For this, we use the QCsnp() function.

```r
## Female-specific QC
finput <- "PostimputeEX_QC3_XFemale"
foutput <- "PostimputeEX_QC4_XFemale"
geno <- 0.05
maf <- 0.05
casecontrol <- TRUE
hweControl <- NULL
hweCase <- NULL
monomorphicSNPs <- FALSE
caldiffmiss <- TRUE
diffmissFilter <- TRUE
dmissX <- TRUE # Since our input has X chromosome, setting this as TRUE wouldn’t change the result.
dmissAutoY <- FALSE # Since our input has X chromosome, setting this as TRUE wouldn’t change the
         result.
x <- QCsnp(DataDir = DataDir, ResultDir = ResultDir, finput = finput,foutput = foutput, geno = geno,
         maf = maf,hweCase = hweCase, hweControl = hweControl, ld_prunning = ld_prunning, casecontrol =
         casecontrol, monomorphicSNPs = monomorphicSNPs, caldiffmiss = caldiffmiss, diffmissFilter =
         diffmissFilter, dmissX = dmissX, dmissAutoY = dmissAutoY)
```

```
## [1] "Program is set up."
## [1] "0 Ambiguous SNPs (A-T/G-C), indels etc. were removed."
## [1] "Thresholds for maf, geno and hwe worked."
## [1] "0 variants removed due to missing genotype data (--geno)."
## [2] "237 variants removed due to minor allele threshold(s)"
## [1] "In cases, "
## [1] "In controls, "
## [1] "Program is set up."
## [1] "Merging is done using the common SNPs between the input genotype files."
```

```
## [1] "Plink files with merged regions are in C:\\Users\\Sarah\\AppData\\Local\\Temp\\RtmpEtlLko
prefixed as filtered_temp2"
## [1] "No SNP with differential missingness between cases and controls."
## [1] "Output plink files prefixed as ,PostimputeEX_QC4_XFemale, with passed SNPs are saved in
ResultDir."
## [1] "Input file has 938 SNPs."
## [1] "Output file has 701 SNPs after filtering."
```

```
## For male-specific QC:
finput <- "PostimputeEX_QC3_XMale"
foutput <- "PostimputeEX_QC4_XMale"
x <- QCsnp(DataDir = DataDir, ResultDir = ResultDir, finput = finput,foutput = foutput, geno = geno,
         maf = maf,hweCase = hweCase, hweControl = hweControl, ld_prunning = ld_prunning, casecontrol =
         casecontrol, monomorphicSNPs = monomorphicSNPs, caldiffmiss = caldiffmiss, diffmissFilter =
         diffmissFilter, dmissX = dmissX, dmissAutoY = dmissAutoY)
```

```
## [1] "Program is set up."
## [1] "0 Ambiguous SNPs (A-T/G-C), indels etc. were removed."
## [1] "Thresholds for maf, geno and hwe worked."
## [1] "0 variants removed due to missing genotype data (--geno)."
## [2] "244 variants removed due to minor allele threshold(s)"
## [1] "In cases, "
## [1] "In controls, "
## [1] "Program is set up."
## [1] "Merging is done using the common SNPs between the input genotype files."
## [1] "Plink files with merged regions are in C:\\Users\\Sarah\\AppData\\Local\\Temp\\RtmpEtlLko
prefixed as filtered_temp2"
## [1] "No SNP with differential missingness between cases and controls."
## [1] "Output plink files prefixed as ,PostimputeEX_QC4_XMale, with passed SNPs are saved in
ResultDir."
## [1] "Input file has 938 SNPs."
## [1] "Output file has 694 SNPs after filtering."
```

```
## Removing the previous QC-ed file from DataDir and copying the new QC-ed file to DataDir.
ftemp <- list.files(paste0(DataDir,"/"),pattern = "PostimputeEX_QC3")
invisible(file.remove(paste0(DataDir,"/",ftemp)))
## Removing the previous QC-ed file from DataDir and copying the new QC-ed file to DataDir.

ftemp <- list.files(paste0(ResultDir,"/"),pattern = "PostimputeEX_QC3")
invisible(file.remove(paste0(ResultDir,"/",ftemp)))

## Copying sex-specific plink files
ftemp <- list.files(paste0(ResultDir,"/"),pattern = "PostimputeEX_QC4_X")
invisible(file.copy(paste0(ResultDir,"/",ftemp),DataDir))
ftemp <- list.files(paste0(ResultDir,"/"),pattern = "PostimputeEX_QC4")
invisible(file.remove(paste0(ResultDir,"/",ftemp)))
```

##### Combine sex-specific QC-ed X chromosomal dataset using common SNPs

```
finput1 <- "PostimputeEX_QC4_XFemale"
finput2 <-
foutput <- "PostimputeEX_QC5_Xchr"
y <- MergeRegion(DataDir, ResultDir, finput1, finput2, foutput, use_common_snps = TRUE)
```

```
## [1] "Program is set up."
## [1] "Merging is done using the common SNPs between the input genotype files."
## [1] "Plink files with merged regions are in C:\\Users\\Sarah\\AppData\\Local\\Temp\\RtmpEtlLko
prefixed as PostimputeEX_QC5_Xchr"
```

```
## Copying merged plink files to DataDir
ftemp <- list.files(paste0(ResultDir,"/"),pattern = "PostimputeEX_QC5_Xchr")
invisible(file.copy(paste0(ResultDir,"/",ftemp),DataDir))
invisible(file.remove(paste0(ResultDir,"/",ftemp)))
## Removing other QC-ed files from DataDir
ftemp <- list.files(paste0(DataDir,"/"),pattern = "PostimputeEX_QC4")
invisible(file.remove(paste0(DataDir,"/",ftemp)))
```

##### Test for HWE across the X chromosome in females

Test for HWE across the X chromosome in females (cases and controls combined), and remove any SNPs violating HWE from analysis for all samples.

```r
finput <- "PostimputeEX_QC5_Xchr"
foutput <- "PostimputeEX_Xchr"
x <- Xhwe(DataDir = DataDir, ResultDir = ResultDir, finput = finput, foutput = foutput, filterSNP =
         TRUE)
```

```
## [1] "Program is set up."
## [1] "Program is set up."
## [1] "Output plink files, prefixed as female, are in
C:\\Users\\Sarah\\AppData\\Local\\Temp\\RtmpEtlLko"
## This test is running on a case-control dataset with female samples.
## [1] "Failed SNPs are excluded from the output plink files prefixed as PostimputeEX_Xchr is in
C:\\Users\\Sarah\\AppData\\Local\\Temp\\RtmpEtlLko"
```

```r
## No. of the SNPs failed the test.
length(x)# 2 SNPs in X chr in females failed HWE test.
```

```
## [1] 2
```

```r
## The failed SNPs
x
```

```
## [1] "rs56053951" "rs12353847"
```

```r
## Removing the previous QC-ed file from DataDir and copying the new QC-ed file to DataDir.
ftemp <- list.files(paste0(DataDir,"/"),pattern = "PostimputeEX_QC5_Xchr")
invisible(file.remove(paste0(DataDir,"/",ftemp)))

## Copying new QC-ed plink files to DataDir
ftemp <- list.files(paste0(ResultDir,"/"),pattern = "PostimputeEX_Xchr")
invisible(file.copy(paste0(ResultDir,"/",ftemp),DataDir))
```

**Check for Xchromosome SNPs with sex difference in MAF in controls**

This test should be performed only for binary traits. We will flag the failed SNPs, if any.

```
finput <- "PostimputeEX_Xchr"
foutput <- "Test_output"
x <- MAFdiffSexControl(DataDir, ResultDir, finput,filterSNP = FALSE,foutput = foutput)
```

```
## [1] "Program is set up."
## [1] "No SNP to be flagged or excluded."
```

```
x
```

```
## NULL
```

#### Combine autosomal and X chromosomal QC-ed datasets

```
finput1 <- "PostimputeEX_Auto"
finput2 <- "PostimputeEX_Xchr"
foutput <- "PostimputeQC"
use_common_snps = FALSE
y <- MergeRegion(DataDir, ResultDir, finput1, finput2, foutput, use_common_snps = FALSE)
```

```
## [1] "Program is set up."
## [1] "Merging is done with all the SNPs i.e., union of the SNPs."
## [1] "Plink files with merged regions are in C:\\Users\\Sarah\\AppData\\Local\\Temp\\RtmpEtlLko
prefixed as PostimputeQC"
```

```
## Removing the previous QC-ed file from DataDir and copying the new QC-ed file to DataDir.
ftemp <- list.files(paste0(DataDir,"/"),pattern = "PostimputeEX")
invisible(file.remove(paste0(DataDir,"/",ftemp)))

ftemp <- list.files(paste0(ResultDir,"/"),pattern = "PostimputeQC")
invisible(file.copy(paste0(ResultDir,"/",ftemp),DataDir))
invisible(file.remove(paste0(ResultDir,"/",ftemp)))
```

##### Prepare separate male and female genotype datasets for sample level QC

```
# Making female-specific plink files
finput <- "PostimputeQC"
foutput <- "Postimpute_Female"
sex <- "females"
x <- GetMFPlink(DataDir = DataDir, ResultDir = ResultDir, finput = finput, foutput = foutput,sex = sex,
         xplink = FALSE, autoplink = FALSE)
```

```
## [1] "Program is set up."
## [1] "Output plink files, prefixed as Postimpute_Female, are in
C:\\Users\\Sarah\\AppData\\Local\\Temp\\RtmpEtlLko"
```

```
# Making male-specific plink files
finput <- "PostimputeQC"
foutput <- "Postimpute_Male"
sex <- "males"
x <- GetMFPlink(DataDir = DataDir, ResultDir = ResultDir, finput = finput, foutput = foutput,sex = sex,
         xplink = FALSE, autoplink = FALSE)
```

```
## [1] "Program is set up."
## [1] "Output plink files, prefixed as Postimpute_Male, are in
C:\\Users\\Sarah\\AppData\\Local\\Temp\\RtmpEtlLko"
```

```
## Copying sex-specific plink files to DataDir
ftemp <- list.files(paste0(ResultDir,"/"),pattern = "Postimpute_Female")
invisible(file.copy(paste0(ResultDir,"/",ftemp),DataDir))
invisible(file.remove(paste0(ResultDir,"/",ftemp)))

ftemp <- list.files(paste0(ResultDir,"/"),pattern = "Postimpute_Male")
invisible(file.copy(paste0(ResultDir,"/",ftemp),DataDir))
invisible(file.remove(paste0(ResultDir,"/",ftemp)))
```

#### Sex-specific sample level QC

For each sex-specific PLINK file, remove individuals with missing genotype rate > 0.1, with absolute value of the heterozygosity F statistic > 0.20, and IBD statistics pihat > 0.2.

For this, we use QCsample().

```
## Running the function female-specific QC
finput <- "Postimpute_Female"
foutput <- "Postimpute_Female1"
imiss = 0.02
#Since het = 3 was removing a lot of samples (185), I decided to increase the threshold after looking
         at the plot.
het = 3
small_sample_mod = TRUE
IBD = 0.2
x = QCsample(DataDir = DataDir,ResultDir = ResultDir, finput = finput,foutput = foutput, imiss =
         imiss,het = het, small_sample_mod = small_sample_mod, IBD = IBD)
```

```
## [1] "Program is set up."
## [1] "No. of ambiguous samples filtered out: 0"
## [1] "Plots are initiated."
```

```
## [1] "Output plink files, Postimpute_Female1 with final samples are in
C:\\Users\\Sarah\\AppData\\Local\\Temp\\RtmpEtlLko."
```

```
## Running the function male-specific QC

finput <- "Postimpute_Male"
foutput <- "Postimpute_Male1"
imiss = 0.02
#Since het = 3 was removing a lot of samples (185), I decided to increase the threshold after looking
         at the plot.
het = 5
small_sample_mod = TRUE
IBD = 0.2
x = QCsample(DataDir = DataDir,ResultDir = ResultDir, small_sample_mod = small_sample_mod, finput =
         finput,foutput = foutput, imiss = imiss,het = het, IBD = IBD)
```

```
## [1] "Program is set up."
## [1] "No. of ambiguous samples filtered out: 0"
## [1] "Plots are initiated."
```

```
## [1] "No samples filtered for missingness."
## [1] "2 samples filtered for heterozygosity."
## [1] "2 samples filtered for missingness and heterozygosity."
## [1] "No sample is filtered out for IDB after missingness and heterozygosity filter."
## [1] "No. of samples in input plink files: 151"
## [1] "No. of samples in output plink files: 149"
```

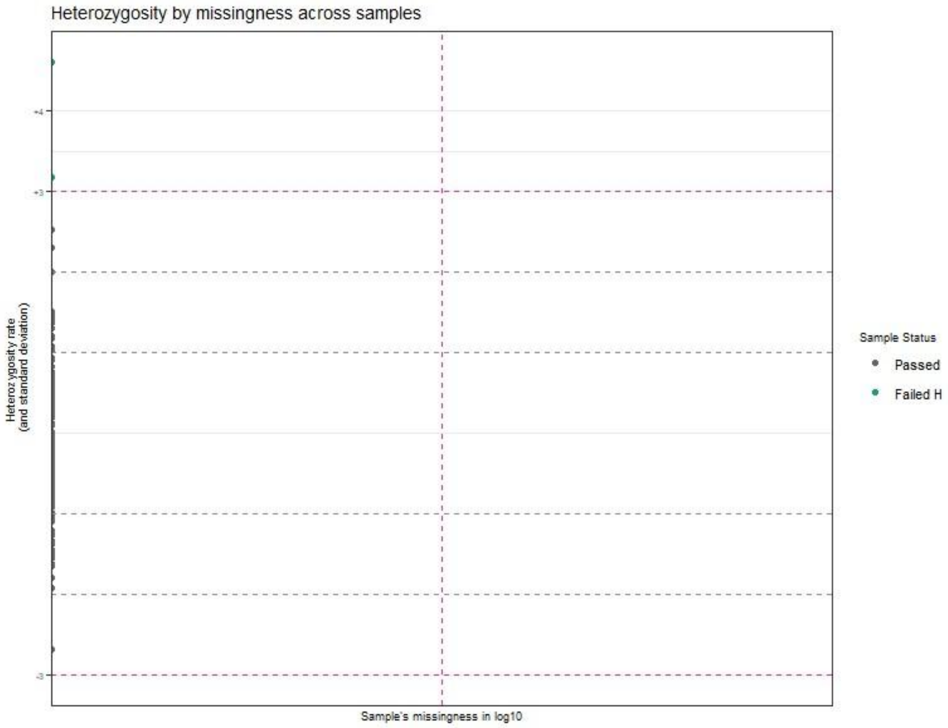

Figure 5: Plot of heteozygosity by missingness across female samples.

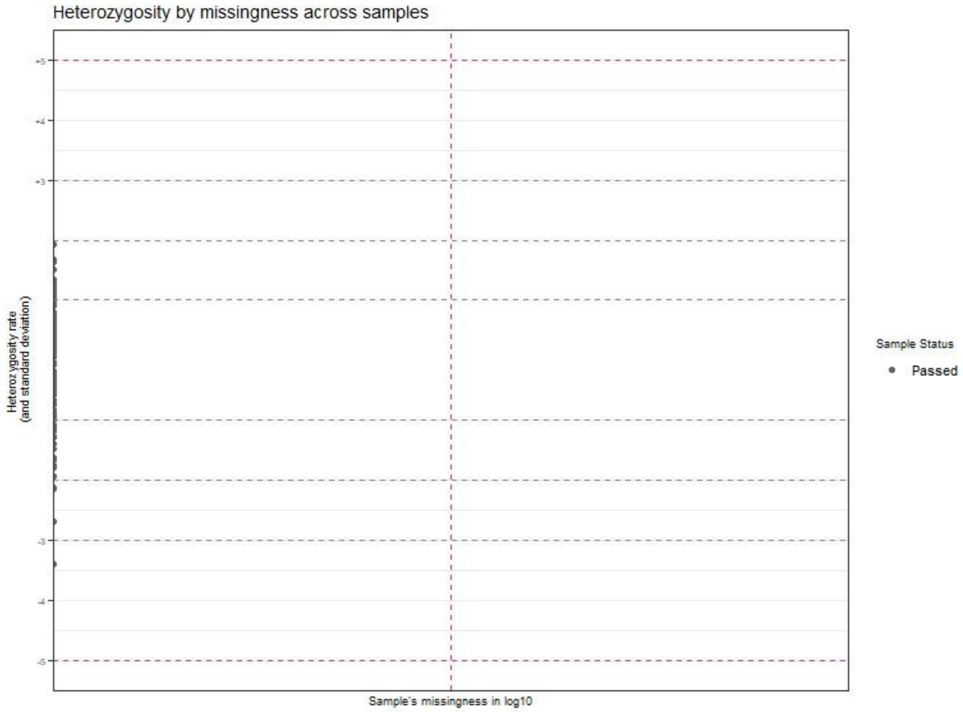

Figure 6 : Plot of heteozygosity by missingness across male samples.

```
## [1] "No samples filtered for missingness."
## [1] "No samples filtered for heterozygosity."
## [1] "No samples filtered for missingness and heterozygosity."
## [1] "No sample is filtered out for IDB after missingness and heterozygosity filter."
## [1] "No. of samples in input plink files: 125"
## [1] "No. of samples in output plink files: 125"

## [1] "Output plink files, Postimpute_Male1 with final samples are in
C:\\Users\\Sarah\\AppData\\Local\\Temp\\RtmpEtlLko."
```

```
## Copying filtered plink files to DataDir
ftemp <- list.files(paste0(ResultDir,"/"),pattern = "Postimpute_")
invisible(file.copy(paste0(ResultDir,"/",ftemp),DataDir))
invisible(file.remove(paste0(ResultDir,"/",ftemp)))
```

### Combine male and female-specific QC-ed genotype datasets

```
## Combine male and female-specific files
## Running the function
finput1 <- "Postimpute_Female1"
finput2 <- "Postimpute_Male1"
foutput <- "PostimputeFinal"
y <- MergeRegion(DataDir, ResultDir, finput1, finput2, foutput, use_common_snps = TRUE)
```

```
## [1] "Program is set up."
## [1] "Merging is done using the common SNPs between the input genotype files."
## [1] "Plink files with merged regions are in C:\\Users\\Sarah\\AppData\\Local\\Temp\\RtmpEtlLko
prefixed as PostimputeFinal"
```

```
## Removing the not needed files from DataDir.
ftemp <- list.files(paste0(DataDir,"/"),pattern = "Postimpute_")
invisible(file.remove(paste0(DataDir,"/",ftemp)))
```

### Summary of the final QC-ed genotype dataset

```
finput <- "PostimputeFinal"
x <- PlinkSummary(ResultDir, ResultDir, finput)
```

```
## [1] "Program is set up."
## [1] "Dataset:PostimputeFinal"
## [1] "This is a case-control data."
## [1] "Number of males:125"
## [1] "Number of females:149"
## [1] "Number of missing phenotypes:0"
## [1] "Number of chromosomes:11"
##  [1] "Chr:1"  "Chr:2"  "Chr:3"  "Chr:4"  "Chr:5"  "Chr:6"  "Chr:7"  "Chr:8"
##  [9] "Chr:9"  "Chr:10" "Chr:23"
## [1] "Total number of SNPs:16063"
## [1] "Total number of samples:274"
```

#### Tutorial for running GWAS, XWAS, and sex-differential effect-size tests using GXwasR

Here we demonstrate the pipeline to run GWAS with XWAS models, and tests for sex-differentiated SNP-trait effects using post-imputation QC'd data.

##### Running GXwas

```
## Running
DataDir <- system.file("extdata", package = "GXwasR")
ResultDir = tempdir()
```

```r
finput <- "GXwasR_example"
standard_beta = TRUE
xsex = FALSE
sex = TRUE
Inphenocov = NULL
covartest = NULL
interaction = FALSE
MF.na.rm = FALSE
B = 10000
MF.zero.sub = 0.00001
trait = "binary"
xmodel = "FMstatrified"
combtest = "fisher.method"
snp_pval = 1e-08
covarfile = NULL
ncores = 0
plot.jpeg = FALSE
MF.mc.cores = 1
genomewideline = 7.3
suggestiveline = 5
plotname = "GXwas.plot"
annotateTopSnp = FALSE
MF.na.rm = FALSE
B = 10000
MF.zero.sub = 0.00001
MF.p.corr = "none"
plotname = "GXwas.plot"
ResultGXwas <- GXwas(DataDir = DataDir,ResultDir = ResultDir, finput = finput, xmodel = xmodel, trait =
         trait, covarfile = covarfile, sex = sex, xsex = xsex, combtest = combtest, MF.p.corr = "none",
         snp_pval = snp_pval, plot.jpeg = plot.jpeg, suggestiveline = 5, genomewideline =
         7.3,MF.mc.cores = 1, ncores = ncores)
```

```
## [1] "Program is set up."
## [1] "Running FMstatrified model"
## [1] "Program is set up."
## [1] "Stratified test is running"
## [1] "Program is set up."

## [1] "Stratified test is running"
## [1] "If you want parallel computation, please provide non-zero value for argument ncores."
```

```
## [1] "If you want parallel computation, please provide non-zero value for argument ncores."
```

```
## [1] "Plots are initiated."
```

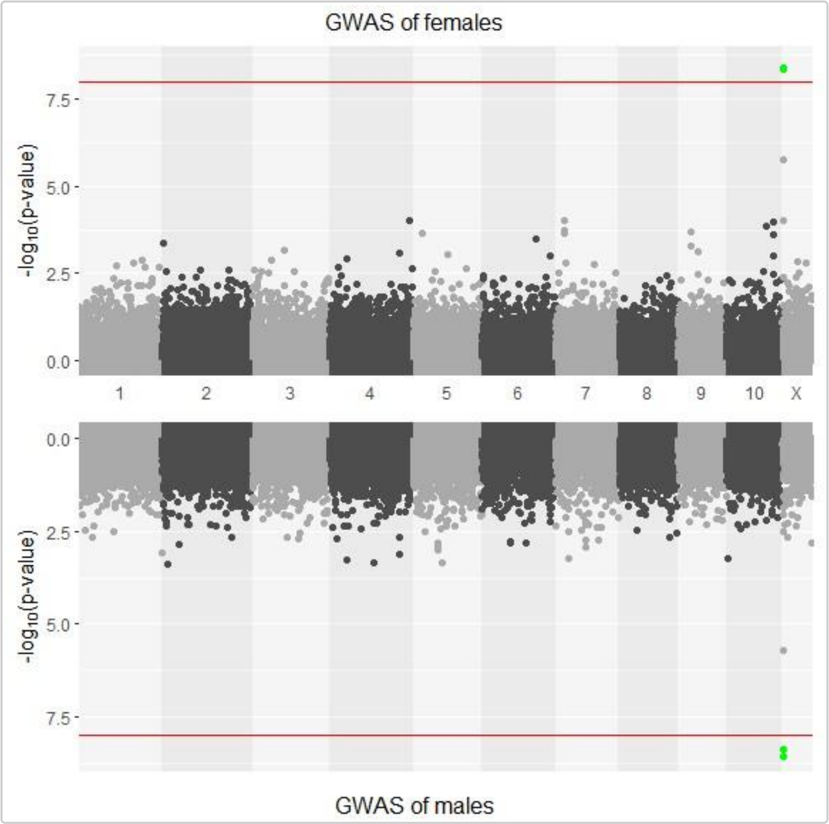

Figure 7 : Miami plot of male vs female GWAS.

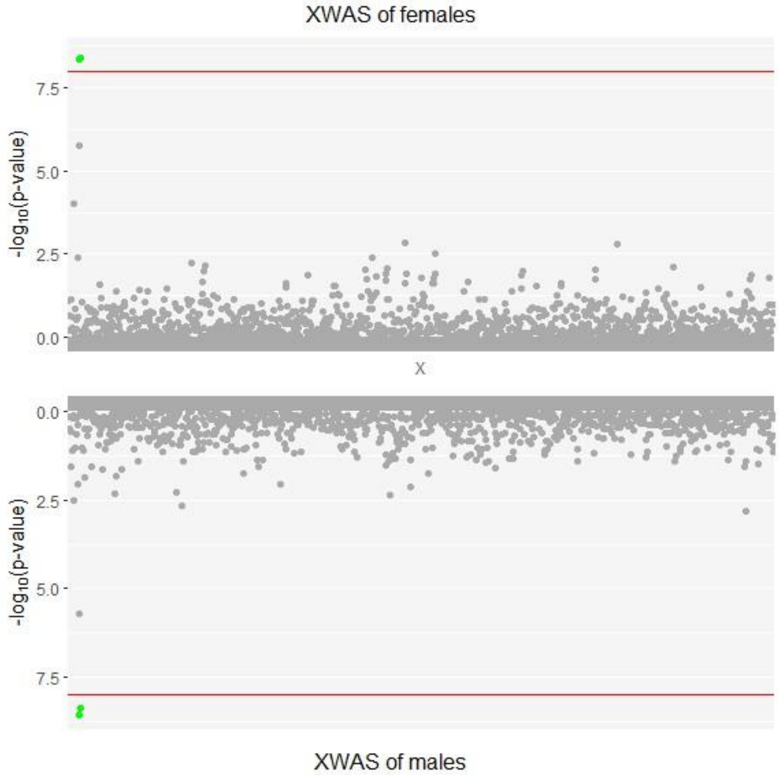

Figure 8: Miami plot of male vs female XWAS.

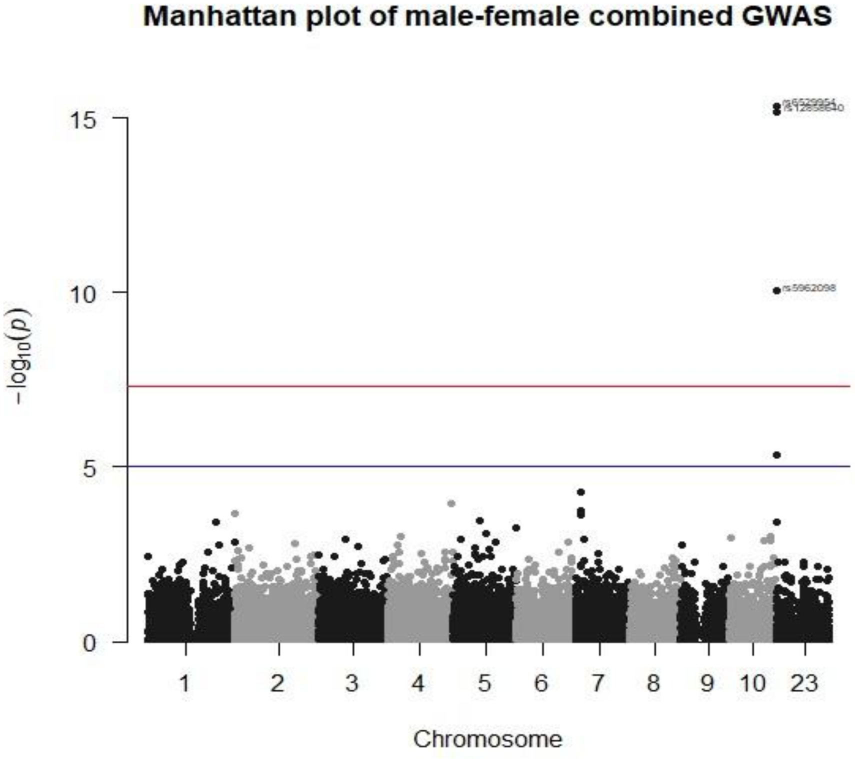

Figure 9: Manhattan plot of the combined-sex GWAS, generated by aggregating p-values across males and females from the sex-stratified tests for each SNP.

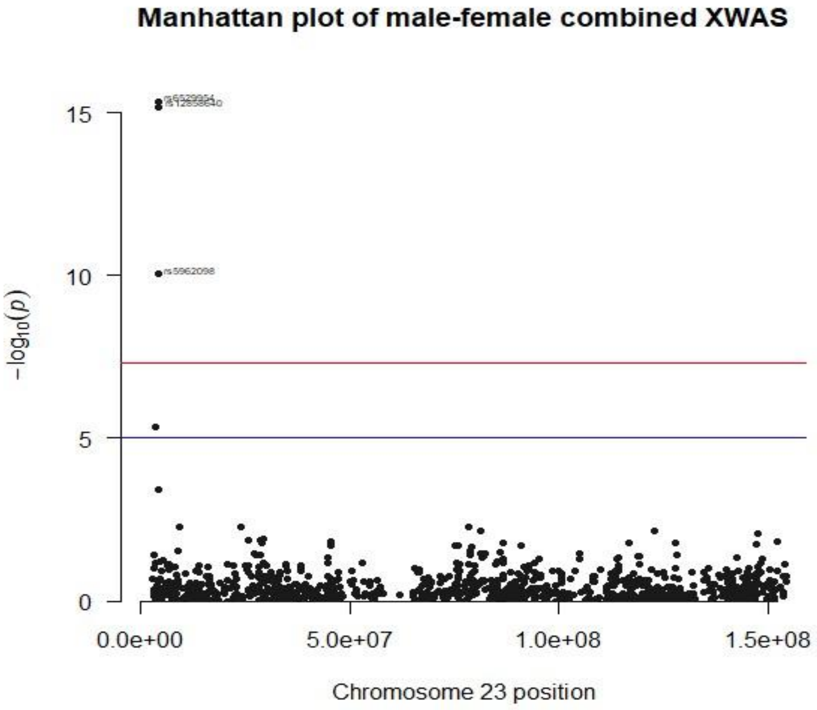

Figure 10: Manhattan plot of the combined-sex XWAS, generated by aggregating p-values across males and females from the sex-stratified tests for each SNP.

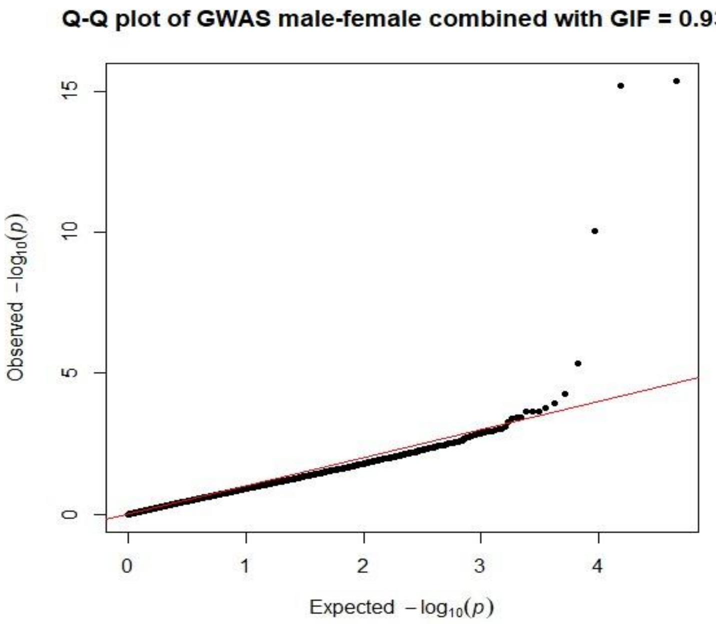

Figure 11: QQ plot of the combined-sex GWAS, generated by aggregating p-values across males and females from the sex-stratified tests for each SNP.

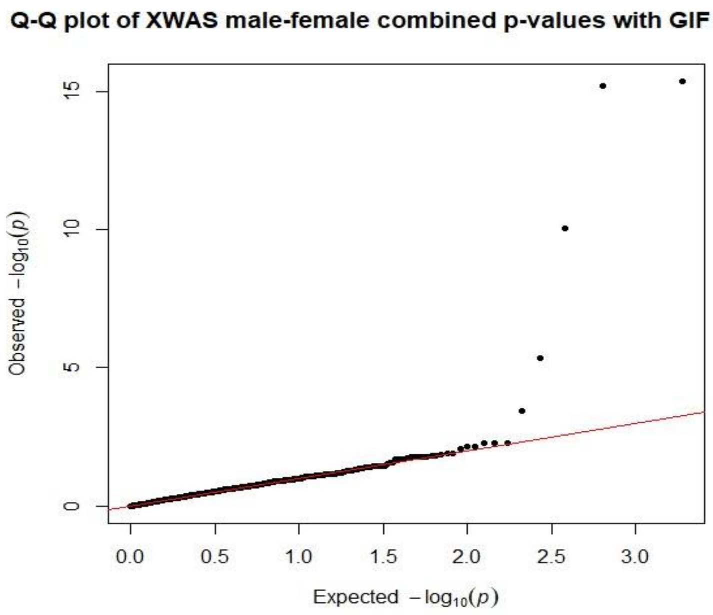

Figure 12: QQ plot of the combined-sex XWAS, generated by aggregating p-values across males and females from the sex-stratified tests for each SNP.

```
## [1] "Three dataframes such as. CombinedWAS. MaleWAS and FemaleWAS are produced
inC:\\Users\\AppData\\Local\\Temp\\RtmpUXS5LP"
```

Dataframe containing the result of the sex-stratified model

```
knitr::kable(ResultGXwas$MaleWAS[1:5,], caption = 'Example of stratified GWAS result with male cohort.')
```

Table 34: Sex-stratified GWAS result, male cohort, five SNPs.

| CHR | SNP | BP | A1 | TEST | NMISS | BETA | SE | L95 | U95 | STAT | P |
|---|---|---|---|---|---|---|---|---|---|---|---|
| 1 | rs143773730 | 73841 | T | ADD | 125 | -0.0789 | 0.2643 | -0.5968 | 0.4390 | -0.2986 | 0.76530 |
| 1 | rs147281566 | 775125 | T | ADD | 125 | -0.3959 | 1.2380 | -2.8230 | 2.0310 | -0.3197 | 0.74920 |
| 1 | rs35854196 | 863863 | A | ADD | 125 | 1.0500 | 0.8858 | -0.6864 | 2.7860 | 1.1850 | 0.23600 |
| 1 | rs12041521 | 1109154 | A | ADD | 125 | -0.4451 | 0.3387 | -1.1090 | 0.2188 | -1.3140 | 0.18890 |
| 1 | rs148527527 | 1127860 | G | ADD | 125 | 1.6770 | 0.6864 | 0.3317 | 3.0220 | 2.4430 | 0.01456 |

```
knitr::kable(ResultGXwas$FemaleWAS[1:5,], caption = 'Example of stratified GWAS result with female cohort.')
```

Table 35: Sex-stratified GWAS result, female cohort, five SNPs.

| CHR | SNP | BP | A1 | TEST | NMISS | BETA | SE | L95 | U95 | STAT | P |
|---|---|---|---|---|---|---|---|---|---|---|---|
| 1 | rs143773730 | 73841 | T | ADD | 151 | 0.3459 | 0.2818 | -0.2064 | 0.89820 | 1.2270 | 0.21970 |
| 1 | rs147281566 | 775125 | T | ADD | 151 | 0.1568 | 0.9290 | -1.6640 | 1.97800 | 0.1688 | 0.86590 |
| 1 | rs35854196 | 863863 | A | ADD | 151 | -0.1446 | 0.7282 | -1.5720 | 1.28300 | -0.1985 | 0.84260 |
| 1 | rs115490086 | 928969 | T | ADD | 151 | 0.5649 | 1.4240 | -2.2270 | 3.35700 | 0.3966 | 0.69170 |
| 1 | rs12041521 | 1109154 | A | ADD | 151 | -0.6697 | 0.3314 | -1.3190 | -0.02008 | -2.0210 | 0.04332 |

```r
## Top ten associations in female-specific study:
load(paste0(ResultDir,"/","FemaleWAS.Rda"))
x1 <- FemaleWAS
x2 <- x1[x1$TEST=="ADD",]
x3 <- x2[order(x2$P),]
x <- x3[1:10, -c(5:6)]
```

```r
## Top ten associations in male-specific study:
load(paste0(ResultDir,"/","MaleWAS.Rda"))
x1 <- MaleWAS
x2 <- x1[x1$TEST=="ADD",]
x3 <- x2[order(x2$P),]
x <- x3[1:10, -c(5:6)]
```

```r
## Top ten associations in FM02comb model:
load(paste0(ResultDir,"/","CombinedWAS.Rda"))
x1 <- CombinedWAS
x2 <- x1[order(x1$P),]
x <- x2[1:10,]
```

#### Testing for sex-differentiated genetic effects

##### Running SexDiff

```r
## Running
## Running the function
x1 <- MaleWAS
x1 <- x1[x1$TEST=="ADD",]
x1 <- x1[,c(1:4,7:8)]
colnames(x1)<- c("CHR","SNP","BP","A1","BETA_M","SE_M")
x2 <- FemaleWAS
x2 <- x2[x2$TEST=="ADD",]
x2 <- x2[,c(1:4,7:8)]
colnames(x2)<- c("CHR","SNP","BP","A1","BETA_F","SE_F")
Difftest <- SexDiff(Mfile=x1,Ffile=x2)
```

```r
## Significant SNPs with sex-differential effect
sig.snps <- Difftest[Difftest $adjP <0.05,]
colnames(sig.snps) <- c("SNP","CHR","BP","A1","T_statistic","Pvalue","Adjusted_Pvalue" )
knitr::kable(sig.snps, caption = 'SNPs with significant sex-differential effect.' )
```

Table 36: SNPs with sex-differences in effect size, according to t-test.

| SNP | CHR | BP | A1 | T_statistic | Pvalue | Adjusted_Pvalue |
|---|---|---|---|---|---|---|
| rs10448254 | 9 | 123107908 | A | 14.806192 | 0.0e+00 | 0.0000000 |
| rs17023642 | 4 | 96292396 | G | 8.651162 | 0.0e+00 | 0.0000000 |
| rs1437407 | 2 | 21060301 | C | 8.224730 | 0.0e+00 | 0.0000000 |

| | | | | | | |
|---|---|---|---|---|---|---|
| rs12013178 | 23 | 53292827 | A | 7.826765 | 0.0e+00 | 0.0000000 |
| rs16848716 | 2 | 164470044 | T | 6.994229 | 0.0e+00 | 0.0000000 |
| rs115673262 | 6 | 32572742 | G | 6.871507 | 0.0e+00 | 0.0000001 |
| rs34327427 | 3 | 178423302 | G | 6.663956 | 0.0e+00 | 0.0000003 |
| rs114724684 | 10 | 83565239 | A | 6.301273 | 0.0e+00 | 0.0000034 |
| rs5952262 | 23 | 44658959 | T | 6.179342 | 0.0e+00 | 0.0000075 |
| rs6520493 | 23 | 38403163 | C | 5.841292 | 0.0e+00 | 0.0000602 |
| rs13440910 | 23 | 38370234 | C | 5.404919 | 0.0e+00 | 0.0007523 |
| rs2214279 | 23 | 9277424 | G | 4.856426 | 6.0e-07 | 0.0138459 |
| rs113460214 | 23 | 154103785 | G | 4.826912 | 7.0e-07 | 0.0160622 |
| rs73560159 | 8 | 33911457 | C | 4.810213 | 8.0e-07 | 0.0174634 |
| rs6641395 | 23 | 147448153 | C | 4.638433 | 1.8e-06 | 0.0406462 |
| rs1766730 | 1 | 247742268 | A | 4.617747 | 1.9e-06 | 0.0449127 |

### Tutorial for computing polygenic risk score

#### Computing standard PRS using ComputePRS():

##### Running ComputePRS()

```
# Running
DataDir <- system.file("extdata", package = "GXwasR")
```

```r
ResultDir <- tempdir()
finput <- "GXwasR_example"
summarystat <- Summary_Stat_Ex1[,c(2,4,7,1,3,12)]
phenofile <- Example_phenofile #Cannot be NULL, the interested phenotype column should be labeled as
         "Pheno1".
covarfile <- Example_covarfile
clump_p1 = 0.0001
clump_p2 = 0.0001
clump_kb = 500
clump_r2 = 0.5
byCHR = TRUE
pthreshold <- Example_pthresoldfile$Threshold
ld_prunning <- TRUE
highLD_regions <- highLD_hg19
window_size <- 50
step_size <- 5
r2_threshold <- 0.02
nPC = 6 #We can incorporate PCs into our PRS analysis to account for population stratification.
pheno_type = "binary"
ldclump = FALSE
pheno_type = "binary"
PRSresult <-
         ComputePRS(DataDir,ResultDir,finput,summarystat,phenofile,covarfile,effectsize="BETA",LDreference
         = "GXwasR_example", ldclump = FALSE,clump_p1,clump_p2,clump_r2,clump_kb, byCHR = TRUE,pthreshold
         = pthreshold, highLD_regions = highLD_regions, ld_prunning = TRUE,window_size = 50, step_size =
         5,r2_threshold = 0.02, nPC = 6,pheno_type = "binary")
```

```
## [1] "Program is set up."
## [1] "Program is set up."
## [1] "plotPC = FALSE was chosen"
## [1] 0.001
## [1] "Computing PRS for threshold 0.001"
## [1] 0.05
## [1] "Computing PRS for threshold 0.05"
## [1] 0.1
## [1] "Computing PRS for threshold 0.1"
## [1] 0.2
## [1] "Computing PRS for threshold 0.2"
## [1] 0.3
## [1] "Computing PRS for threshold 0.3"
## [1] 0.4
## [1] "Computing PRS for threshold 0.4"
## [1] 0.5
```

```
## [1] "Computing PRS for threshold 0.5"

## [1] "Plots are initiated."

## [1] "Plots are printed."
```

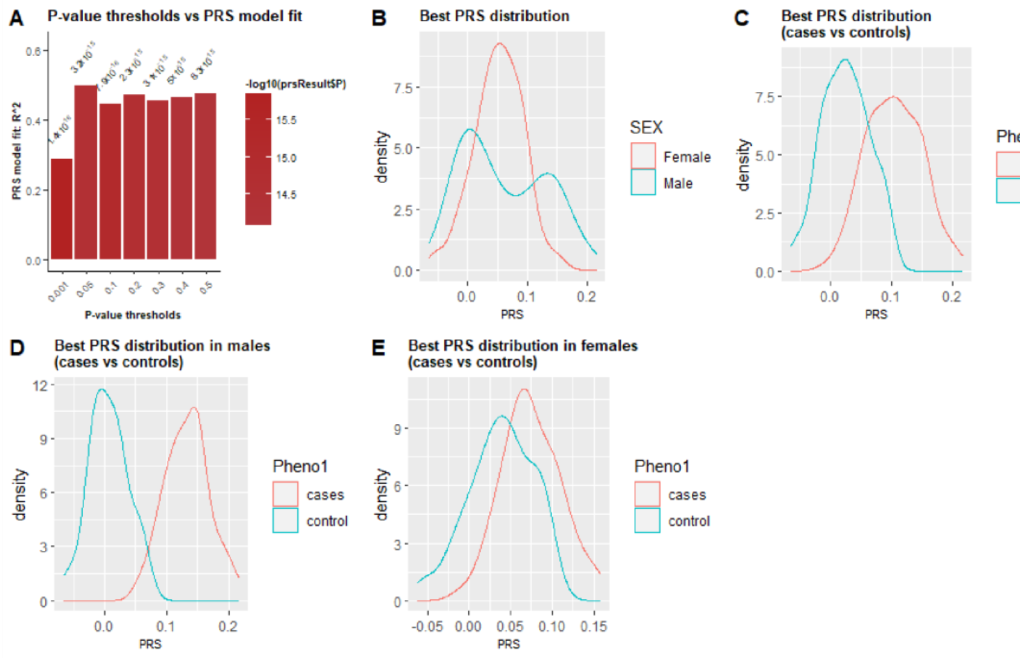

Figure 14 : Polygenic Risk Score (PRS) analysis showing: A) model fit across p-value thresholds; PRS distributions by: B) sex, C) case-control status in the full sample, D) stratified by case-control status within males and E) females.

```
## This table shows 10 samples with phenotype, covariates and a PRS column.
PRS <- PRSrsult$PRS
PRS[1:10,]
```

Table 37: A table showing ten samples with phenotypes, covariates and a PRS score column.

| FID | IID | Pheno1 | AGE | testcovar | PC1 | PC2 | PC3 | PC4 | PC5 | PC6 | SCORE |
|---|---|---|---|---|---|---|---|---|---|---|---|
| EUR_FIN | HG00171 | 1 | 36 | 1 | 0.0984270 | 0.0545964 | -0.0128387 | -0.0295421 | 0.0381968 | -0.0338884 | 0.0484212 |
| EUR_FIN | HG00173 | 1 | 81 | 1 | 0.0814620 | 0.0427825 | -0.0353089 | -0.0046323 | -0.0547033 | 0.0234367 | 0.0293742 |
| EUR_FIN | HG00174 | 1 | 83 | 1 | 0.0574856 | 0.0231881 | 0.0074347 | 0.0018039 | -0.0072442 | 0.0242692 | 0.0099674 |
| EUR_FIN | HG00176 | 2 | 75 | 0 | 0.0846743 | 0.0531902 | -0.0239229 | -0.0125275 | 0.0185402 | -0.0232204 | 0.1042060 |
| EUR_FIN | HG00177 | 1 | 88 | 1 | 0.0844408 | 0.0447869 | -0.0623942 | -0.0351411 | -0.0208355 | 0.0764725 | 0.0314303 |
| EUR_FIN | HG00178 | 1 | 24 | 1 | 0.0590789 | 0.0356180 | 0.0225026 | 0.0310004 | 0.0463228 | 0.0212882 | 0.0722689 |
| EUR_FIN | HG00179 | 2 | 78 | 1 | 0.0858355 | 0.0325418 | -0.0140910 | 0.0264206 | -0.0449786 | 0.0054808 | 0.1165240 |
| EUR_FIN | HG00180 | 1 | 39 | 1 | 0.0926786 | 0.0389852 | 0.0080128 | 0.0158442 | 0.0253621 | -0.0609059 | 0.0617159 |
| EUR_FIN | HG00182 | 1 | 50 | 1 | 0.0884415 | 0.0535143 | -0.0150184 | 0.0237222 | -0.0630822 | -0.0074837 | 0.0283703 |
| EUR_FIN | HG00183 | 1 | 58 | 0 | 0.0841885 | 0.0497837 | -0.0166280 | -0.0271598 | -0.0113098 | -0.0216147 | 0.0386297 |

```
## The best threshold
BestPvalue <- PRSresult$BestP$Threshold
BestPvalue
```

```
## [1] 0.05
```

### Computing sex-aware PRS

#### Datasets for computing PRS

**Discovery Data** i.e., GWAS summary statistics with mandatory columns: “SNP”(SNP names), “A1”(effect allele), “BETA”(effect size in beta value), and “P”(p-value).

```
#Example discovery data is included in GXwasR
library(GXwasR)
data("GXwasRData")
summarystat <- Summary_Stat_Ex1[,c(2,4,7,1,3,12)]
```

**Target Data** i.e., genotype dataset in plink .bed, .bim and .fam format.

```
# Example target data is included in GXwasR
DataDir <- system.file("extdata", package = "GXwasR")

finput <- "GXwasR_example"
```

##### Quality control of the datasets before computing sex-aware PRS.

First ensure that both discovery and target datasets have the **same genome build**. Next, perform the following quality control procedure:

**Discovery Data:** Remove multi-allelic, indels and ambiguous (A/T or C/G) SNPs. Then remove SNPs with minor allele frequency (MAF) < 0.05 and quality info score < 0.1. For this filtering, R packages like data.table, dplyr, tidyverse can be used.

**Target Data:** Remove multi-allelic, indels and ambiguous (A/T or C/G) SNPs. Then remove SNPs with MAF < 0.05 and Hardy Weinberg Equlibrium (hwe) > e-10. For this filtering, users can use FilterAllele() and QCsnp() functions in R.

Use FilterAllele () on target data to filter out any multi-allelic SNPs.

```
#Target data
DataDir <- system.file("extdata", package = "GXwasR")
ResultDir <- tempdir()
finput <- "GXwasR_example"
foutput <- "filtered_multiallelic"
x <- FilterAllele(DataDir, ResultDir, finput, foutput)
```

```
## [1] "Program is set up."
## [1] "There is no multi-allelic SNP present in the input dataset."
```

Use QCsnp() to remove ambiguous (A/T or C/G) SNPs and SNPs with MAF < 0.05 and Hardy Weinberg Equlibrium (hwe) > e-10.

```r
##Since there was no multiallelic SNPs, we will continue with original input data.
finput <- "GXwasR_example"
foutput <- "filtered_step1"
geno <- NULL
maf <- 0.05
casecontrol <- FALSE
caldiffmiss <- FALSE
diffmissFilter <- FALSE
dmissX <- FALSE
dmissAutoY <- FALSE
monomorphicSNPs <- TRUE
ld_prunning <- FALSE

casecontrol <- FALSE ## Since the filtering doesn't require us to run on cases and controls separately,
         we will make this parameter FALSE.
hweCase <- NULL
hweControl <- NULL
hwe <- 1e-10
monomorphicSNPs <- FALSE
ld_prunning <- FALSE

x <- QCsnp(DataDir = DataDir, ResultDir = ResultDir, finput = finput,foutput = foutput, geno = geno,
         maf = maf,hweCase = hweCase, hweControl = hweControl, hwe = hwe, ld_prunning = ld_prunning,
         casecontrol = casecontrol, monomorphicSNPs = monomorphicSNPs, caldiffmiss = caldiffmiss,
         dmissX = dmissX, dmissAutoY = dmissAutoY, diffmissFilter = diffmissFilter)
```

```
## [1] "Program is set up."
## [1] "4214 Ambiguous SNPs (A-T/G-C), indels etc. were removed."
## [1] "Thresholds for maf, geno and hwe worked."
## [1] "--hwe: 3 variants removed due to Hardy-Weinberg exact test."
## [2] "5467 variants removed due to minor allele threshold(s)"
## [1] "No filter based on differential missingness will be applied."
## [1] "Output plink files prefixed as ,filtered_step1, with passed SNPs are saved in ResultDir."
## [1] "Input file has 26527 SNPs."
## [1] "Output file has 16843 SNPs after filtering."
```

**Check whether SNPs present in discovery data are in target data.** Gather SNPs common to discovery and target datasets.

```r
SNP1 <- unique(summarystat$SNP)
targetbim <- read.table(paste0(ResultDir,"/filtered_step1.bim"))
SNP2 <- unique(targetbim$V2)
commonSNP <- intersect(SNP1,SNP2)## 991 SNPs are common between our discovery data and target data
```

#### Filter discovery and target data to contain only shared SNPs

```
commonSNP <- data.table::as.data.table(commonSNP)
colnames(commonSNP) <- "SNP"
NewDiscoveryData <- merge(commonSNP,summarystat,by = "SNP")
## For making target data with common SNPs, we will use FilterSNP().
SNPvec <-commonSNP
#Need to copy filtered_step1 file from ResultDir to DataDir
ftemp <- list.files(paste0(ResultDir,"/"),pattern = "filtered_step1")
invisible(file.copy(paste0(ResultDir,"/",ftemp),DataDir))
finput <- "filtered_step1"

foutput <- "NewtargetData"
FilterSNP(DataDir, ResultDir, finput, foutput, SNPvec,extract = TRUE)
```

```
## [1] "Program is set up."
## [1] "991 SNPs are extracted"
## [1] "Plink files with extracted SNPs are in C:\\Users\\Sarah\\AppData\\Local\\Temp\\RtmpuuME3B
prefixed as NewtargetData"
```

```
## NULL
```

Now, discovery and target datasets are ready for computing sex-combined and sex-stratified PRS.

##### Sex-combined PRS computation

A. Perform LD-clumping of discovery data using target data as reference.

B. Compute PRS at a variety of p-value thresholds.

C. Select the best threshold based on R-square (for a quantitative trait) or MacFadden R-square (for a binary trait) using a generalized linear model with covariates, e.g., genetic PCs.

Use ComputePRS() to perform steps (A), (B), and (C).

```
# Running
# Filtered target data needs to be copied from ResultDir to DataDir.
ftemp <- list.files(paste0(ResultDir,"/"),pattern = "NewtargetData")
invisible(file.copy(paste0(ResultDir,"/",ftemp),DataDir))
finput <- "NewtargetData"

# Filtered discovery data.
# Need to maintain the first three coulmn of this dataset as SNP ID, Effect Allele and Effect Size
summarystat <- NewDiscoveryData
phenofile <- Example_phenofile #Cannot be NULL, the interested phenotype column should be labeled as
         "Pheno1".
## Added "AGE" and "testcovar" as covariates.
covarfile <- Example_covarfile
clump_p1 = 0.0001
clump_p2 = 0.0001
clump_kb = 500
clump_r2 = 0.5
byCHR = TRUE
pthreshold <- Example_pthresoldfile$Threshold
ld_prunning <- TRUE
```

```r
highLD_regions <- highLD_hg19
window_size <- 50
step_size <- 5
r2_threshold <- 0.02
nPC = 6  # We can incorporate PCs into our PRS analysis to account for population stratification.
pheno_type = "binary"
effectsize = "BETA"
ldclump = FALSE
PRSresult <-
         ComputePRS(DataDir,ResultDir,finput,summarystat,phenofile,covarfile,effectsize="BETA",LDreference
         = "GXwasR_example", ldclump = FALSE,clump_p1,clump_p2,clump_r2,clump_kb, byCHR = TRUE,pthreshold
         = pthreshold,highLD_regions = highLD_regions, ld_prunning = TRUE,window_size = 50, step_size =
         5,r2_threshold = 0.02, nPC = 6,pheno_type = "binary")
```

```
## [1] "Program is set up."
## [1] "Program is set up."
## [1] "plotPC = FALSE was chosen"
## [1] 0.001
## [1] "Computing PRS for threshold 0.001"
## [1] 0.05
## [1] "Computing PRS for threshold 0.05"
## [1] 0.1
## [1] "Computing PRS for threshold 0.1"
## [1] 0.2
## [1] "Computing PRS for threshold 0.2"
## [1] 0.3
## [1] "Computing PRS for threshold 0.3"
## [1] 0.4
## [1] "Computing PRS for threshold 0.4"
## [1] 0.5
## [1] "Computing PRS for threshold 0.5"
## [1] "Plots are printed."
```

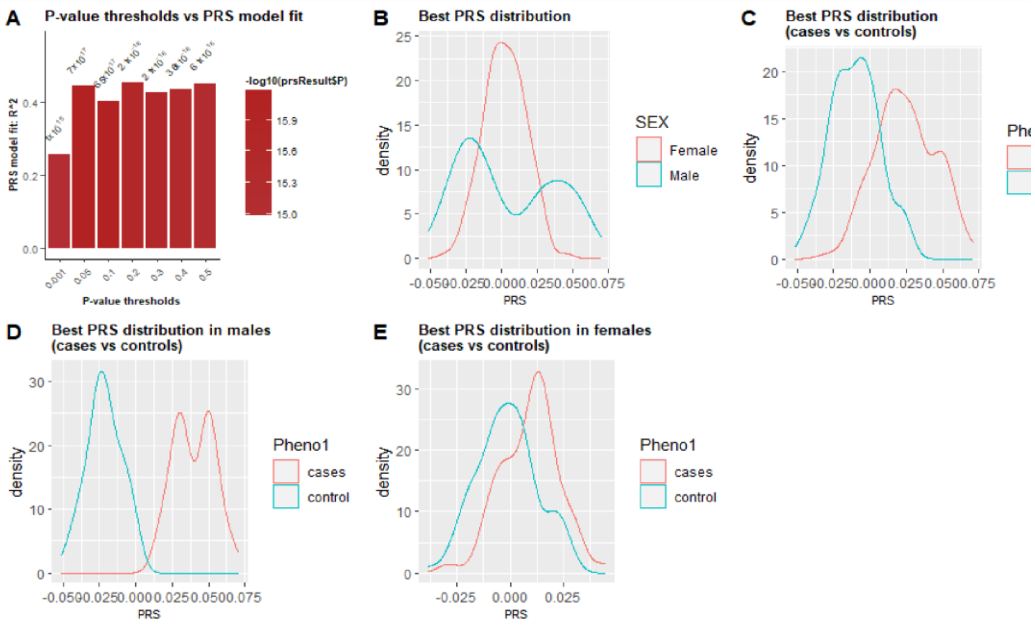

Figure 15 : PRS analysis. (A) Sex-combined PRS analysis showing model fit across p-value thresholds,  PRS distributions by: (B) sex, (C) case-control status , and PRS stratified by: case-control status within: (D) males, and (E) females.

```r
## This table shows 10 samples with phenotype, covariates and a PRS column.
PRS <- PRSresult$PRS
PRS[1:10,]
```

Table 38: A table showing 10 samples with phenotype, covariates, and sex-combined PRS scores.

| FID | IID | Pheno1 | AGE | testcovar | PC1 | PC2 | PC3 | PC4 | PC5 | PC6 | SCORE |
|---|---|---|---|---|---|---|---|---|---|---|---|
| EUR_FIN | HG00171 | 1 | 36 | 1 | 0.1152000 | 0.0845964 | -0.0528387 | -0.0225421 | 0.3381968 | -0.5338884 | 0.0184212 |
| EUR_FIN | HG00173 | 1 | 81 | 1 | 0.0685788 | 0.0827825 | -0.0395089 | -0.1246323 | -0.0547033 | 0.1234367 | 0.1293742 |
| EUR_FIN | HG00174 | 1 | 83 | 1 | 0.0322192 | 0.0237567 | -0.020812 | 0.0480590 | 0.0142846 | 0.0012434 | 0.0030211 |
| EUR_FIN | HG00176 | 2 | 75 | 0 | 0.0923099 | 0.0945430 | -0.0258776 | 0.0720050 | -0.061009 | -0.0227889 | 0.0297140 |
| EUR_FIN | HG00177 | 1 | 88 | 1 | 0.0894694 | 0.0507977 | -0.0201713 | -0.012165 | 0.0514540 | -0.009474 | -0.002032 |
| EUR_FIN | HG00178 | 1 | 24 | 1 | 0.0535609 | -0.026909 | 0.0531649 | 0.0230253 | 0.0040255 | 0.0438350 | 0.0115410 |
| EUR_FIN | HG00179 | 2 | 78 | 1 | 0.0010501 | 0.0279390 | -0.1512810 | -0.047068 | -0.0353966 | 0.1357360 | 0.0112993 |
| EUR_FIN | HG00180 | 1 | 39 | 1 | 0.0495020 | -0.123686 | -0.158575 | -0.007801 | -0.120689 | -0.0168742 | 0.0234045 |
| EUR_FIN | HG00182 | 1 | 50 | 1 | 0.0833507 | 0.0209397 | 0.0113593 | -0.033259 | -0.0321080 | -0.0223653 | -0.019635 |
| EUR_FIN | HG00183 | 1 | 58 | 0 | 0.0411108 | -0.041677 | 0.0282441 | 0.0208968 | -0.031037 | 0.1355450 | -0.008747 |

```r
## The best threshold
BestPvalue <- PRSresult$BestP$Threshold
BestPvalue
```

```
## [1] 0.2
```

D. Perform regression to test the association between PRS and sex.

E. Perform the regression using different thresholds of PRS to check for consistent evidence of sex difference.

In this regression, we can include genetic PCs and and other covariates.

**PRS ~ Sex + PCs + Other Covariates.**

For this test, we use SexRegress().

##### Run SexRegress()

```
# Running
# First, we need to make fdata object.
library(GXwasR)
famfile <- read.table(paste0(ResultDir,"/NewtargetData.fam"))[,c(1,2,5)]
colnames(famfile) <- c("FID","IID","Sex")
prefdata <- merge(famfile, PRS, by = c("IID","FID"))

fdata <- prefdata[,c(13,3:12)]
fdata$Sex <- as.factor(as.character(fdata$Sex))
response_index <- 1
regressor_index <- 2

x <- SexRegress(fdata, regressor_index, response_index)
x
```

```
##      Estimate    Std. Error       t value      Pr(>|t|)
## -0.0001303925  0.0021759687 -0.0599239012  0.9522614090
```

In this example, the association between PRS and sex is not significant. Users can test across different p-value thresholds of PRS as mentioned in step (E).

##### Sex-stratified PRS computation

The steps are:

(A) Generate separate male and female discovery data using sex-stratified GWAS.

Use SumstatMale and SumstatFemale for these.

(B) Prepare separate male and female target datasets.

Use GetMFPlink().

```
library(GXwasR)
## We will use NewtargetData as input for this function.
finput <- "NewtargetData"
foutput <- "maletarget"
sex <- "males"
x <- GetMFPlink(DataDir = DataDir, ResultDir = ResultDir, finput = finput, foutput = foutput,sex = sex,
         xplink = FALSE, autoplink = FALSE)
```

```
## [1] "Program is set up."
## [1] "Output plink files, prefixed as maletarget, are in
C:\\Users\\Sarah\\AppData\\Local\\Temp\\RtmpuuME3B"
```

```
foutput <- "femaletarget"
sex <- "females"
x <- GetMFPlink(DataDir = DataDir, ResultDir = ResultDir, finput = finput, foutput = foutput,sex = sex,
         xplink = FALSE, autoplink = FALSE)
```

```
## [1] "Program is set up."
## [1] "Output plink files, prefixed as femaletarget, are in
C:\\Users\\Sarah\\AppData\\Local\\Temp\\RtmpuuME3B"
```

(C) Perform LD clumping separately in male and female discovery datasets.

(D) Compute PRS across p-value thresholds by using male-only discovery data to compute scores for female-only target data and vice versa. Alternatively, same-sex comparisons can also be performed, such as using male discovery data for male targets and female discovery data for female targets.

(E) Select the best threshold based on R-square (for a quantitative trait) or MacFadden R-square (for a binary trait) using a generalized linear model with covariates, e.g., genetic PCs.

Use ComputePRS() to perform the steps (C), (D) and (E).

```
# Running
# Male and female target datasets need to be copied from ResultDir to DataDir.
ftemp <- list.files(paste0(ResultDir,"/"),pattern = "target")
invisible(file.copy(paste0(ResultDir,"/",ftemp),DataDir))
```

##### Compute PRS for female-only target data using male-only discovery data

Evaluate the transferability of genetic risk prediction across sexes and to test whether associations identified in one sex generalize to the other.

```
DataDir <- system.file("extdata", package = "GXwasR")
ResultDir <- tempdir()
finput <- "femaletarget"
# Filtered discovery data.
# Need to maintain the first three coulmn of this dataset as SNP ID, Effect Allele and Effect Size
summarystat <- Summary_Stat_Ex1[,c(2,4,7,1,3,12)]
## Making phenofile only with females
famfile <- read.table(paste0(ResultDir,"/femaletarget.fam"))[,c(1,2,5)]
colnames(famfile) <- c("FID","IID","Sex")
famfileF <- famfile[famfile$Sex== 2,]
phenofile <- merge(famfileF,Example_phenofile,by = c("FID","IID")) #Cannot be NULL, the interested
         phenotype column should be labeled as "Pheno1".
## Added "AGE" and "testcovar" as covariates.
## Making cpvarfile with females
covarfile <- merge(famfileF,Example_covarfile,by = c("FID","IID"))
```

```r
clump_p1 = 0.0001
clump_p2 = 0.0001
clump_kb = 500
clump_r2 = 0.5
byCHR = TRUE
pthreshold <- Example_pthresoldfile$Threshold
ld_prunning <- TRUE
highLD_regions <- highLD_hg19
window_size <- 50
step_size <- 5
r2_threshold <- 0.02
nPC = 6  # We can incorporate PCs into our PRS analysis to account for population stratification.
pheno_type = "binary"
ldclump = FALSE
effectsize="BETA"
PRSresultFemale <-
         ComputePRS(DataDir,ResultDir,finput,summarystat,phenofile,covarfile,effectsize="BETA",LDreference
         = "GXwasR_example", ldclump = FALSE,clump_p1,clump_p2,clump_r2,clump_kb, byCHR = TRUE,pthreshold
         = pthreshold,highLD_regions = highLD_regions, ld_prunning = TRUE,window_size = 50, step_size =
         5,r2_threshold = 0.02, nPC = 6,pheno_type = "binary")
```

```
## [1] "Program is set up."
## [1] "Program is set up."
## [1] "plotPC = FALSE was chosen"
## [1] 0.001
## [1] "Computing PRS for threshold 0.001"
## [1] 0.05
## [1] "Computing PRS for threshold 0.05"
## [1] 0.1
## [1] "Computing PRS for threshold 0.1"
## [1] 0.2
## [1] "Computing PRS for threshold 0.2"
## [1] 0.3
## [1] "Computing PRS for threshold 0.3"
## [1] 0.4
## [1] "Computing PRS for threshold 0.4"
## [1] 0.5
## [1] "Computing PRS for threshold 0.5"
## [1] "Plots are printed."
```

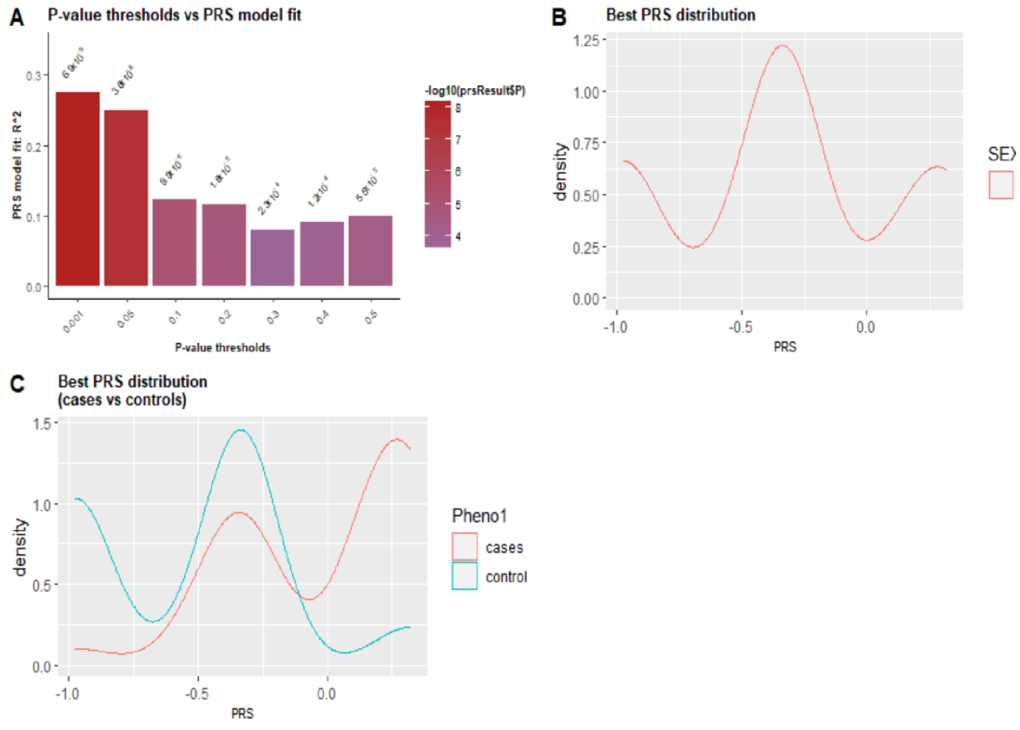

Figure 16 : Female-specific PRS analysis showing: (A) model fit across p-value thresholds, PRS distributions in: (B) females, and (C) stratified by case-control status within females.

```
## This table shows 10 samples with phenotype, covariates and a PRS column.
PRSfemale <- PRSresultFemale$PRS
PRSfemale[1:10,]
```

Table 39: A table showing eight male samples with phenotype, covariates, and with female-specific PRS scores.

| FID | IID | Pheno1 | AGE | testcovar | PC1 | PC2 | PC3 | PC4 | PC5 | PC6 | SCORE |
|---|---|---|---|---|---|---|---|---|---|---|---|
| EUR_FIN | HG00171 | 1 | 36 | 1 | 0.1152000 | 0.0845964 | -0.0528387 | -0.0225421 | 0.3381968 | -0.5338884 | 0.0184212 |
| EUR_FIN | HG00173 | 1 | 81 | 1 | 0.0685788 | 0.0827825 | -0.0395089 | -0.1246323 | -0.0547033 | 0.1234367 | 0.1293742 |
| EUR_FIN | HG00174 | 1 | 83 | 1 | -0.021171 | -0.032675 | 0.0101237 | 0.0106363 | 0.0042739 | 0.0071773 | -0.975333 |
| EUR_FIN | HG00176 | 2 | 75 | 0 | -0.106273 | -0.113970 | 0.0105312 | -0.0236382 | 0.0342991 | 0.0904657 | 0.320733 |
| EUR_FIN | HG00177 | 1 | 88 | 1 | -0.100336 | -0.080632 | -0.0054835 | 0.1248180 | 0.0485978 | -0.017674 | -0.975333 |
| EUR_FIN | HG00178 | 1 | 24 | 1 | -0.087763 | -0.052104 | -0.046601 | 0.0323986 | -0.039007 | -0.057816 | 0.320733 |
| EUR_FIN | HG00179 | 2 | 78 | 1 | -0.012028 | 0.0215276 | 0.2411880 | -0.141282 | -0.0107047 | 0.0137807 | -0.327300 |
| EUR_FIN | HG00180 | 1 | 39 | 1 | -0.141166 | 0.1431790 | 0.1060060 | -0.249856 | -0.131981 | 0.0020415 | -0.327300 |
| EUR_FIN | HG00182 | 1 | 50 | 1 | -0.105387 | -0.013805 | 0.0405550 | -0.030058 | 0.0165495 | 0.0197359 | 0.320733 |
| EUR_FIN | HG00183 | 1 | 58 | 0 | 0.0119449 | 0.1796970 | -0.1024400 | -0.0057617 | 0.0680961 | 0.0724916 | -0.327300 |

```r
## The best threshold
BestPvalue <- PRSresultFemale$BestP$Threshold
BestPvalue
```

```
## [1] 0.001
```

#### Compute PRS for male-only target data using female-only discovery data

```r
library(GXwasR)
data("GXwasRData")
DataDir <- system.file("extdata", package = "GXwasR")
ResultDir <- tempdir()
finput <- "maletarget"
# Filtered discovery data.
# Need to maintain the first three coulmn of this dataset as SNP ID, Effect Allele and Effect Size
summarystat <- Summary_Stat_Ex2[,c(2,4,7,1,3,12)]
## Making phenofile only with females
famfile <- read.table(paste0(ResultDir,"/maletarget.fam"))[,c(1,2,5)]
colnames(famfile) <- c("FID","IID","Sex")
famfileM <- famfile[famfile$Sex== 1,]
phenofile <- merge(famfileM,Example_phenofile,by = c("FID","IID")) #Cannot be NULL, the interested
        phenotype column should be labeled as "Pheno1".
## Added "AGE" and "testcovar" as covariates.
## Making cpvarfile with females
covarfile <- merge(famfileM,Example_covarfile,by = c("FID","IID"))
```

```r
clump_p1 = 0.0001
clump_p2 = 0.0001
clump_kb = 500
clump_r2 = 0.5
byCHR = TRUE
pthreshold <- Example_pthresoldfile$Threshold
ld_prunning <- TRUE
highLD_regions <- highLD_hg19
window_size <- 50
step_size <- 5
r2_threshold <- 0.02
nPC = 6  # We can incorporate PCs into our PRS analysis to account for population stratification.
pheno_type = "binary"
ldclump = FALSE
effectsize="BETA"
PRSresultmale <-
         ComputePRS(DataDir,ResultDir,finput,summarystat,phenofile,covarfile,effectsize="BETA",LDreference
         = "GXwasR_example", ldclump = FALSE,clump_p1 = clump_p1,clump_p2 = clump_p2,clump_r2 =
         clump_r2,clump_kb = clump_kb, byCHR = TRUE,pthreshold = pthreshold,highLD_regions =
         highLD_regions, ld_prunning = TRUE,window_size = 50, step_size = 5,r2_threshold = 0.02, nPC =
         6,pheno_type = "binary")
```

```
## [1] "Program is set up."
## [1] "Program is set up."
## [1] "plotPC = FALSE was chosen"
## [1] 0.001
## [1] "Computing PRS for threshold 0.001"
## [1] 0.05
## [1] "Computing PRS for threshold 0.05"
## [1] 0.1
## [1] "Computing PRS for threshold 0.1"
## [1] 0.2
## [1] "Computing PRS for threshold 0.2"
## [1] 0.3
## [1] "Computing PRS for threshold 0.3"
## [1] 0.4
## [1] "Computing PRS for threshold 0.4"
## [1] 0.5
## [1] "Computing PRS for threshold 0.5"
## [1] "Plots are printed."
```

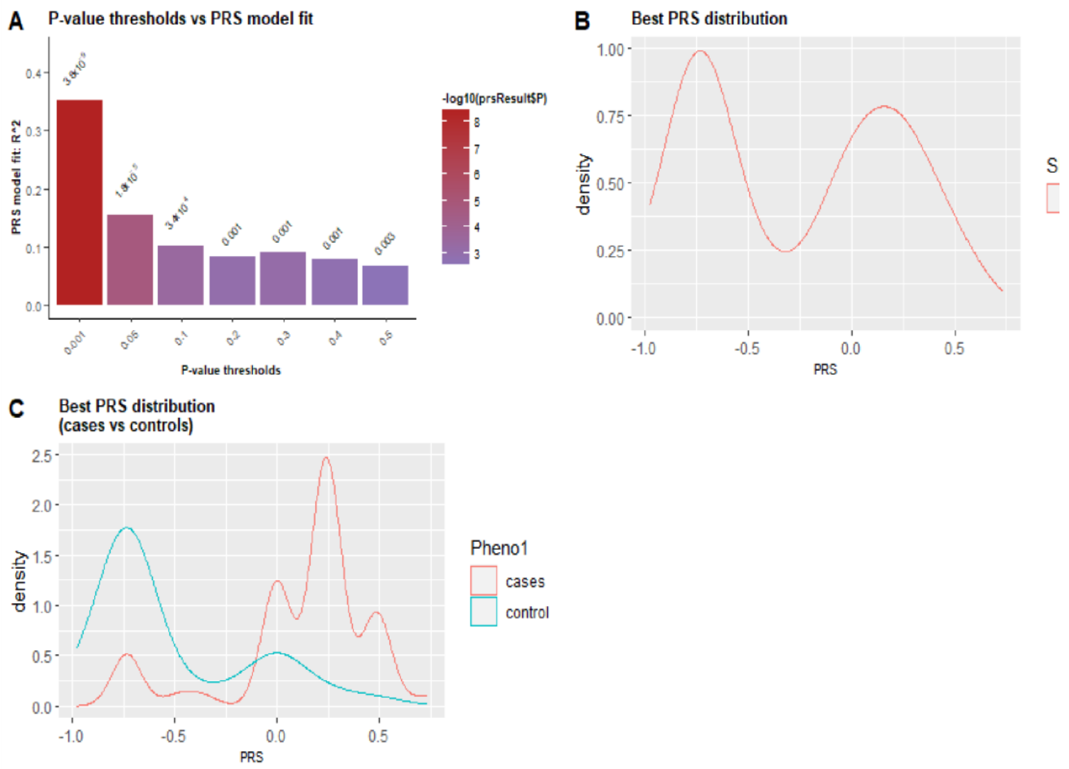

Figure 17 : Male-specific PRS analysis showing: (A) model fit across p-value thresholds, PRS distributions in: (B) males, and (C) stratified by case-control status within males.

```
## The best threshold
BestPvalue <- PRSresultmale$BestP$Threshold
BestPvalue
```

```
## [1] 0.001
```

Table 40: A table showing eight female samples with phenotype, covariates and with male-specific PRS scores.

| FID | IID | Pheno1 | AGE | testcovar | PC1 | PC2 | PC3 | PC4 | PC5 | PC6 | SCORE |
|---|---|---|---|---|---|---|---|---|---|---|---|
| EUR_FIN | HG00185 | 2 | 22 | 1 | -0.057393 | 0.0855428 | -0.046644 | -0.039166 | -0.228309 | -0.152104 | -0.001833 |
| EUR_FIN | HG00186 | 2 | 52 | 0 | -0.158784 | 0.0155813 | -0.017859 | -0.023012 | -0.124404 | 0.0063170 | 0.0000000 |
| EUR_FIN | HG00187 | 2 | 49 | 1 | -0.066489 | 0.1702040 | 0.0382637 | -0.055301 | 0.0094718 | -0.062734 | 0.4873330 |
| EUR_FIN | HG00188 | 1 | 47 | 1 | -0.044187 | 0.0753788 | -0.034038 | 0.1284960 | -0.013723 | 0.0091291 | -0.488333 |
| EUR_FIN | HG00189 | 1 | 27 | 1 | -0.132578 | 0.1367130 | -0.093038 | 0.0484504 | 0.1212130 | -0.197444 | -0.734833 |
| EUR_FIN | HG00190 | 2 | 42 | 0 | 0.1815650 | 0.0989251 | 0.1148980 | 0.0278529 | 0.0288093 | -0.068524 | -0.734833 |
| EUR_FIN | HG00267 | 2 | 36 | 1 | 0.0732148 | 0.1039340 | -0.000889 | -0.135554 | 0.0746865 | 0.0322867 | -0.734833 |
| EUR_FIN | HG00271 | 2 | 51 | 1 | -0.161405 | 0.1023490 | 0.0131086 | 0.0469537 | 0.1181770 | -0.100801 | 0.2408330 |

(F) Perform regression to test the association between the PRS for a trait derived in one sex and case status in the other sex. This approach helps evaluate the cross-sex predictive validity of the PRS and can reveal whether genetic risk factors identified in one sex are relevant or transferable to the other, potentially uncovering sex-specific or shared genetic architecture.

PRS ~ trait + PCs + other covariates

Let's check the association between PRS and trait in female.

```
# Running
# First, we need to make fdata object.
famfile <- read.table(paste0(ResultDir,"/femaletarget.fam"))[,c(1,2,6)]
colnames(famfile) <- c("FID","IID","Trait")
prefdata <- merge(famfile, PRS, by = c("IID","FID"))

fdata <- prefdata[,c(13,3:12)]
fdata$Sex <- as.factor(as.character(fdata$Trait))
response_index <- 1
regressor_index <- 2

x <- SexRegress(fdata, regressor_index, response_index)
x
```

```
##     Estimate   Std. Error      t value     Pr(>|t|)
## 1.059080e-02 2.315619e-03 4.573636e+00 1.040039e-05
```

```
# In this case, there is a significant association between PRS and trait value.
```

### Tutorial for estimating heritability using *EstimateHerit*

#### Run EstimateHerit() with model = 'GREML' separately for each chromosome

Currently, model = 'GREML' is only supported in Unix.

Users can run this model by computing the Genetic Relationship Matrice (GRM) per chromosome or they can use pre-computed GRMs by setting the parameter ComputeGRM <- TRUE or FALSE.

In this example, we set this to 'TRUE'.

```
# Running

DataDir = system.file("extdata", package = "GXwasR")

ResultDir = tempdir()

finput <- "GXwasR_example"

test.sumstats <- na.omit(Summary_Stat_Ex1[Summary_Stat_Ex1$TEST=="ADD",c(1:4,6:8)])

colnames(test.sumstats) <- c("chr","rsid","pos","a1","n_eff","beta","beta_se")

summarystat = test.sumstats

ncores = 3

model = "GREML"

byCHR = FALSE

r2_LD = 0

LDSC_blocks = 20

REMLalgo = 0

nitr = 100

cat_covarfile = NULL

quant_covarfile = NULL
```

```
  prevalance = 0.01
  partGRM = FALSE
  autosome = TRUE
  Xsome = TRUE
  nGRM = 3
  cripticut = 0.025
  minMAF = NULL
  maxMAF = NULL
  hg = "hg19"
  byCHR = TRUE
  PlotIndepSNP = TRUE
  IndepSNP_window_size = 50
  IndepSNP_step_size = 5
  IndepSNP_r2_threshold = 0.02
  highLD_regions = highLD_hg19
  H2 <- EstimateHerit(DataDir =
  DataDir, ResultDir = ResultDir,
  finput = finput, summarystat =
  NULL, ncores, model = model,
  byCHR = TRUE, r2_LD = 0,
  LDSC_blocks = 20,REMLalgo = 0,
  nitr = nitr, cat_covarfile =
  NULL, quant_covarfile =
  NULL,prevalance = 0.01, partGRM =
  FALSE, autosome = TRUE, Xsome =
  TRUE, nGRM = 3,cripticut = 0.025,
  minMAF = NULL, maxMAF = NULL,hg =
              "hg19",PlotIndepSNP = TRUE, IndepSNP_window_size = 50,IndepSNP_step_size = 5,IndepSNP_r2_threshold =
              0.02,highLD_regions = highLD_hg19)
```

```
## [1] "Program is set up."
## [1] "GCTA setup complete. This program is currently only supported in Linux."
## [1] "Processing chromosome 1"
## [1] "Processing chromosome 2"
## [1] "Processing chromosome 3"
## [1] "Processing chromosome 4"
## [1] "Processing chromosome 5"
## [1] "Processing chromosome 6"
## [1] "Processing chromosome 7"
## [1] "Processing chromosome 8"
## [1] "Convergence issue occurs, please check the models, use byCHR = TRUE, check different options, SNP partitioning or quality of the data"
## Reading IDs of the GRM from [/projects/b1137/BBose/Herit/Chr8_GXwasR.grm.id].
## 276 IDs are read from [/projects/b1137/BBose/Herit/Chr8_GXwasR.grm.id].
## Reading the GRM from [/projects/b1137/BBose/Herit/Chr8_GXwasR.grm.bin].
## GRM for 276 individuals are included from [/projects/b1137/BBose/Herit/Chr8_GXwasR.grm.bin].
```

```
## Reading phenotypes from [/projects/b1137/BBose/Herit/phenofile.phen].
## Non-missing phenotypes of 276 individuals are included from [/projects/b1137/BBose/Herit/phenofile.phen].
## Assuming a disease phenotype for a case-control study: 108 cases and 168 controls
## 276 individuals are in common in these files.
##
## Performing  REML analysis ... (Note: may take hours depending on sample size).
## 276 observations, 1 fixed effect(s), and 2 variance component(s)(including residual variance).
## Calculating prior values of variance components by EM-REML ...
## Updated prior values: 0.119115 0.129833
## logL: 48.1674
## Running AI-REML algorithm ...
## Iter.    logL    V(G)    V(e)
## 1    49.00   0.05048 0.18556
## 2    53.68   -0.09416    0.31783
## 3    67.73   -0.09671    0.32598
## 4    66.03   -0.09871    0.33207
## 5    64.63   -0.10388    0.34661
## 6    62.27   -0.10733    0.35109
## 7    60.80   -0.12071    0.36824
## 8    60.65   -0.12892    0.37665
## 9    58.81   7.33365 -9.18968
## 10   -117.27 761.08102   -950.87811
## 11   -764.61 632972.57848    -792333.84535
## 12   -1689.41    1188815819784.33789 -1486907005424.49951
## 13   -3674.51    2763428134209696342999040.00000 -3456651210872768390234112.00000
## Error: the information matrix is not invertible.
## An error occurs, please check the options or data
## [1] "Processing chromosome 9"
## [1] "Processing chromosome 10"
## [1] "Convergence issue occurs, please check the models, use byCHR = TRUE, check different options, SNP partitioning or
quality of the data"
## Reading IDs of the GRM from [/projects/b1137/BBose/Herit/Chr10_GXwasR.grm.id].
## 276 IDs are read from [/projects/b1137/BBose/Herit/Chr10_GXwasR.grm.id].
## Reading the GRM from [/projects/b1137/BBose/Herit/Chr10_GXwasR.grm.bin].
## GRM for 276 individuals are included from [/projects/b1137/BBose/Herit/Chr10_GXwasR.grm.bin].
## Reading phenotypes from [/projects/b1137/BBose/Herit/phenofile.phen].
## Non-missing phenotypes of 276 individuals are included from [/projects/b1137/BBose/Herit/phenofile.phen].
## Assuming a disease phenotype for a case-control study: 108 cases and 168 controls
## 276 individuals are in common in these files.
##
## Performing  REML analysis ... (Note: may take hours depending on sample size).
## 276 observations, 1 fixed effect(s), and 2 variance component(s)(including residual variance).
```

```
## Calculating prior values of variance components by EM-REML ...
## Updated prior values: 0.119126 0.129587
## logL: 48.6719
## Running AI-REML algorithm ...
## Iter.    logL    V(G)    V(e)
## 1    49.47   0.05480 0.18180
## 2    53.72   -0.07424    0.30004
## 3    57.94   -0.08006    0.31082
## 4    58.16   -0.08691    0.32162
## 5    58.39   -0.12212    0.36891
## 6    56.39   -0.07579    0.31133
## 7    58.18   -0.09180    0.33392
## 8    58.57   -0.10523    0.34959
## 9    57.99   -0.00394    0.22819
## 10   56.34   -0.06657    0.30149
## 11   58.07   -0.08325    0.32417
## 12   58.30   -0.08939    0.33122
## 13   58.45   -0.12347    0.37065
## 14   56.26   -0.07816    0.31400
## 15   58.21   -0.09521    0.33782
## 16   58.91   -0.10254    0.34670
## 17   58.34   0.18680 0.00091
## 18   10.88   0.10726 0.09786
## 19   44.41   -0.01797    0.23132
## 20   56.26   -0.03232    0.25246
## 21   57.09   -0.04386    0.26938
## 22   57.56   -0.07299    0.31165
## 23   58.16   -0.08795    0.32961
## 24   58.40   -0.11717    0.36328
## 25   56.87   -0.06529    0.29947
## 26   58.05   -0.08245    0.32324
## 27   58.28   -0.08830    0.32995
## 28   58.41   -0.11865    0.36501
## 29   56.73   -0.06852    0.30311
## 30   58.09   -0.08465    0.32575
## 31   58.32   -0.09146    0.33359
## 32   58.54   -0.13298    0.38191
## 33   55.11   -0.09666    0.33526
## 34   59.31   -0.10646    0.35222
## 35   57.92   -0.07680    0.31625
## 36   58.21   -0.09199    0.33422
```

```
## 37   58.58   -0.13487    0.38420
## 38   54.83   -0.10046    0.33974
## 39   58.31   0.18782 -0.00038
## 40   10.01   0.16322 0.02986
## 41   25.93   0.05597 0.14831
## 42   50.61   0.02575 0.18433
## 43   53.81   0.00090 0.21512
## 44   55.69   -0.01877    0.24024
## 45   56.76   -0.03398    0.26010
## 46   57.36   -0.04567    0.27555
## 47   57.70   -0.07434    0.31350
## 48   58.18   -0.08921    0.33104
## 49   58.44   -0.12259    0.36963
## 50   56.35   -0.07647    0.31209
## 51   58.19   -0.09270    0.33495
## 52   58.63   -0.10647    0.35108
## 53   57.87   -0.01984    0.24694
## 54   57.00   -0.06943    0.30629
## 55   58.12   -0.08514    0.32638
## 56   58.33   -0.09222    0.33451
## 57   58.59   -0.13548    0.38495
## 58   54.73   -0.10170    0.34121
## 59   58.15   0.03593 0.18081
## 60   53.77   0.00673 0.21369
## 61   55.69   -0.06323    0.29640
## 62   58.01   -0.06892    0.30443
## 63   58.10   -0.07396    0.31126
## 64   58.17   -0.08919    0.33100
## 65   58.44   -0.12253    0.36955
## 66   56.36   -0.07634    0.31195
## 67   58.19   -0.09253    0.33475
## 68   58.61   -0.10630    0.35087
## 69   57.88   -0.01814    0.24494
## 70   56.93   -0.06913    0.30580
## 71   58.11   -0.08493    0.32614
## 72   58.33   -0.09189    0.33413
## 73   58.57   -0.13453    0.38378
## 74   54.88   -0.09978    0.33893
## 75   58.42   2.65560 -2.94901
## 76   37.01   98.93694    -109.44542
## 77   -472.12 -8101.89143 8941.35613
```

```
## 78   -1078.91    547968215.40932 -605601840.21001
## 79   -2606.68    -27098667354966118400.00000 29946205646968172544.00000
## Error: the information matrix is not invertible.
## An error occurs, please check the options or data
## [1] "Processing chromosome 23"
## [1] "Convergence issue occurs, please check the models, use byCHR = TRUE, check different options, SNP partitioning or quality of the data"
## An error occurs, please check the options or data
## [1] "All GRM related files are in /projects/b1137/BBose/Herit"
```

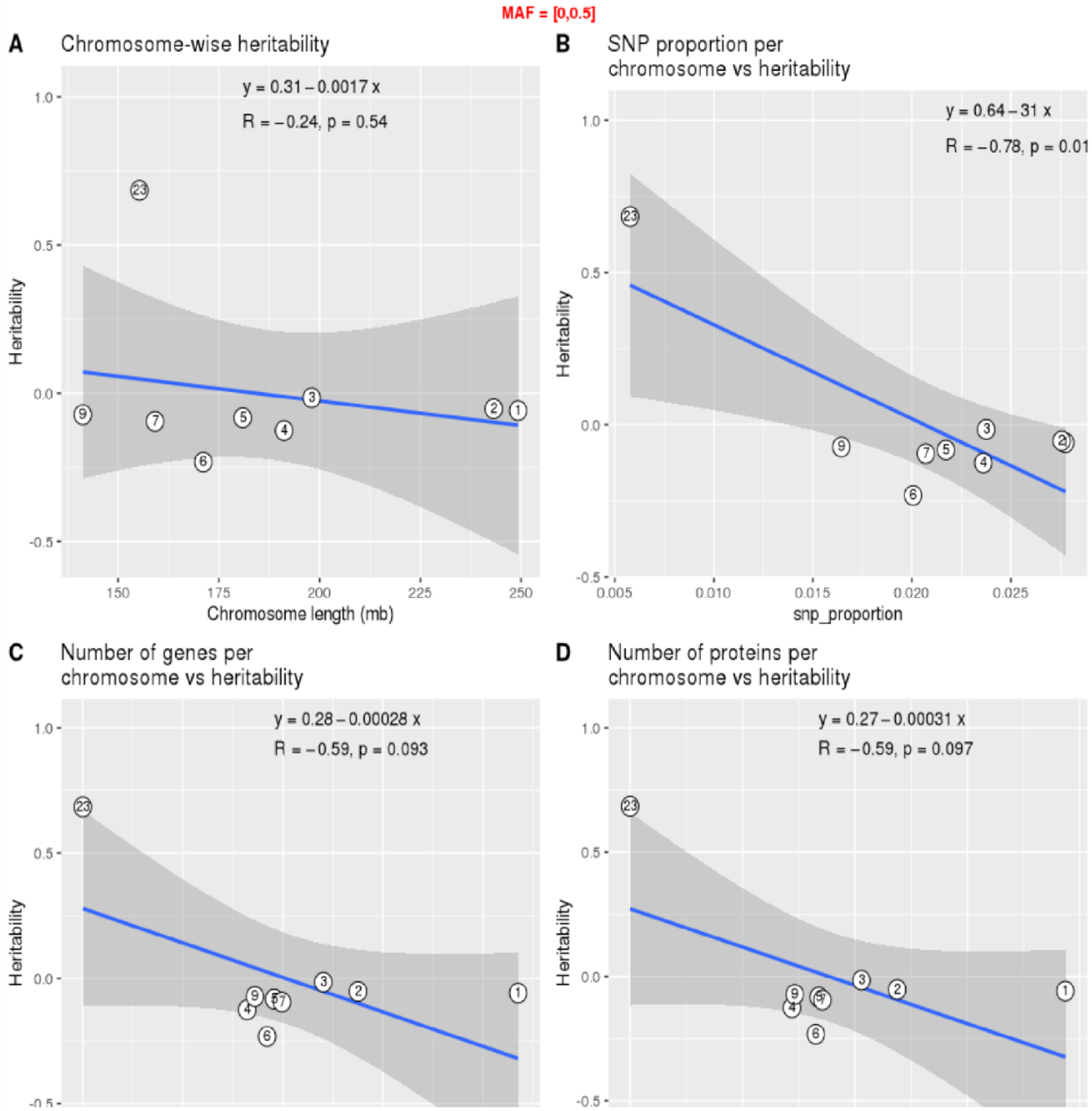

Figure 18: Per chromosome heritability analysis, showing: (A) heritability increases with chromosome length; (B) SNP proportion per chromosome correlates positively with heritability; (C) the number of genes per chromosome shows a moderate positive correlation with heritability; (D) number of proteins per chromosome is positively correlated with heritability.

Table 41: Per chromosome heritability extimates for example genotype datasets using GREML model.

| Chromosome | SNP_proportion | No.of.genes | No.of.proteins | Size_mb | Source | Variance | SE |
|---|---|---|---|---|---|---|---|
| 1 | 0.0277453 | 2175 | 1936 | 249.25062 | V(G) | -0.014132 | 0.053414 |
| 1 | 0.0277453 | 2175 | 1936 | 249.25062 | V(e) | 0.253334 | 0.058554 |
| 1 | 0.0277453 | 2175 | 1936 | 249.25062 | Vp | 0.239202 | 0.020425 |
| 1 | 0.0277453 | 2175 | 1936 | 249.25062 | V(G)/Vp | -0.059081 | 0.222989 |
| 1 | 0.0277453 | 2175 | 1936 | 249.25062 | V(G)/Vp_L | -0.034224 | 0.129174 |
| 2 | 0.0275568 | 1375 | 1188 | 243.19937 | V(G) | -0.012654 | 0.049342 |
| 2 | 0.0275568 | 1375 | 1188 | 243.19937 | V(e) | 0.251833 | 0.054662 |
| 2 | 0.0275568 | 1375 | 1188 | 243.19937 | Vp | 0.239179 | 0.020419 |
| 2 | 0.0275568 | 1375 | 1188 | 243.19937 | V(G)/Vp | -0.052906 | 0.206040 |
| 2 | 0.0275568 | 1375 | 1188 | 243.19937 | V(G)/Vp_L | -0.030647 | 0.119355 |
| 3 | 0.0237494 | 1202 | 1029 | 198.02243 | V(G) | -0.003691 | 0.049497 |
| 3 | 0.0237494 | 1202 | 1029 | 198.02243 | V(e) | 0.242773 | 0.054155 |
| 3 | 0.0237494 | 1202 | 1029 | 198.02243 | Vp | 0.239082 | 0.020396 |
| 3 | 0.0237494 | 1202 | 1029 | 198.02243 | V(G)/Vp | -0.015438 | 0.206990 |
| 3 | 0.0237494 | 1202 | 1029 | 198.02243 | V(G)/Vp_L | -0.008943 | 0.119906 |
| 4 | 0.0235986 | 821 | 720 | 191.15428 | V(G) | -0.030153 | 0.046321 |
| 4 | 0.0235986 | 821 | 720 | 191.15428 | V(e) | 0.269533 | 0.053054 |
| 4 | 0.0235986 | 821 | 720 | 191.15428 | Vp | 0.239380 | 0.020474 |
| 4 | 0.0235986 | 821 | 720 | 191.15428 | V(G)/Vp | -0.125964 | 0.192379 |
| 4 | 0.0235986 | 821 | 720 | 191.15428 | V(G)/Vp_L | -0.072969 | 0.111442 |
| 5 | 0.0217137 | 956 | 839 | 180.91526 | V(G) | -0.019899 | 0.046183 |

#### Run EstimateHerit() with model = 'GREML' genome-wide

In this example, we set byCHR to 'FALSE'.

```
## [1] "Program is set up."

## [1] "GCTA setup complete. This program is currently only supported in Linux."

## [1] "GRM has been saved in the file [/projects/b1137/BBose/Herit/GXwasR.grm.bin]"

## [1] "Number of SNPs in each pair of individuals has been saved in the file
[/projects/b1137/BBose/Herit/GXwasR.grm.N.bin]"

## [1] "GRM has been saved in the file [/projects/b1137/BBose/Herit/xGXwasR.grm.bin]"

## [1] "Number of SNPs in each pair of individuals has been saved in the file
[/projects/b1137/BBose/Herit/xGXwasR.grm.N.bin]"

## [1] "computeGRM is set to: TRUE"

## [1] "Creating multi_GRMs.txt because computeGRM is TRUE"

## [1] "Convergence issue occurs, please check the models, use byCHR = TRUE, check different options, SNP partitioning or
quality of the data"

## Reading IDs of the GRM from [/projects/b1137/BBose/Herit/GXwasR.grm.id].

## 276 IDs are read from [/projects/b1137/BBose/Herit/GXwasR.grm.id].

## Reading the GRM from [/projects/b1137/BBose/Herit/GXwasR.grm.bin].

## GRM for 276 individuals are included from [/projects/b1137/BBose/Herit/GXwasR.grm.bin].

## Reading the GRM from the 2th file ...
```

```
## Reading IDs of the GRM from [/projects/b1137/BBose/Herit/xGXwasR.grm.id].
## 276 IDs are read from [/projects/b1137/BBose/Herit/xGXwasR.grm.id].
## Reading the GRM from [/projects/b1137/BBose/Herit/xGXwasR.grm.bin].
## GRM for 276 individuals are included from [/projects/b1137/BBose/Herit/xGXwasR.grm.bin].
## 276 individuals are in common in these files.
##
## Performing  REML analysis ... (Note: may take hours depending on sample size).
## 276 observations, 1 fixed effect(s), and 3 variance component(s)(including residual variance).
## Calculating prior values of variance components by EM-REML ...
## Updated prior values: 0.0766102 0.0812353 0.0774914
## logL: 65.5415
## Running AI-REML algorithm ...
## Iter.    logL    V(G1)   V(G2)   V(e)
## 1    65.89   -0.06529    0.10883 0.18382
## 2    70.71   -0.25864    0.16268 0.32471
## 3    72.94   -0.23673    0.16287 0.30397
## 4    73.45   -0.20353    0.16373 0.27055
## 5    74.08   -0.45543    0.15421 0.54362
## 6    55.70   -0.42677    0.15010 0.51465
## 7    65.58   -0.36549    0.14547 0.45062
## 8    72.90   -0.40226    0.14409 0.49583
## 9    70.18   -0.33960    0.14395 0.42475
## 10   83.68   -0.35796    0.14304 0.45264
## 11   77.96   -0.36246    0.14063 0.45974
## 12   76.23   -0.38598    0.13463 0.49122
## 13   72.66   -0.55884    0.13430 0.68739
## 14   85.29   -0.55748    0.12346 0.68031
## 15   76.27   -0.61248    0.12069 0.74611
## 16   71.14   0.35778 0.17516 -0.44139
## 17   273.66  -0.00866    0.09609 -0.04373
## 18   -1136.55    -0.00593    0.08610 -0.04096
## 19   -770.22 0.00914 0.02422 -0.02204
## 20   2880.94 0.01149 0.03049 -0.02772
## 21   2290.67 0.01428 0.03797 -0.03449
## 22   1835.73 0.02449 0.06561 -0.05943
## 23   1103.72 0.03753 0.10234 -0.09207
## 24   709.55  0.05006 0.14288 -0.12629
## 25   491.18  0.06037 0.19147 -0.16279
## 26   361.37  0.07242 0.28266 -0.22422
## 27   266.18  0.09459 0.62551 -0.43261
## 28   140.77  -1.21853    -5.67494    4.29047
```

```
## 29   -99.30   -40.58338   -196.81962  147.06877
## 30   -582.74 -54257.98773    -263508.86898   196820.55656
## 31   -1564.04    -3628841.93652  -17623814.47841 13163610.81180
## 32   -2137.22    -4864916.19679  -23626832.30372 17647414.70936
## 33   -2176.86    -5378237.38139  -26119687.47055 19509410.65450
## 34   -2189.91    58950571.90792  286299353.71159 -213843156.25175
## Error: the information matrix is not invertible.
## An error occurs, please check the options or data
## [1] "All GRM related files are in /projects/b1137/BBose/Herit"
```

The output of running the genome-wide heritability model with GREML using example genotype datasets. Since the information matrix is not invertible, it will have NA result.

##### Run EstimateHerit() with model = 'LDSC' per chromosome

Users can run this model by computing the GRM per chromosome or they can use pre-computed GRMs by setting the parameter ComputeGRM <- TRUE or FALSE.

In this example, we set this to 'TRUE'.

```
model <- "LDSC"
data("GXwasRData")
  DataDir = system.file("extdata", package =
  "GXwasR") ResultDir = tempdir() finput <-
  "GXwasR_example"
  test.sumstats <- na.omit(Summary_Stat_Ex1[Summary_Stat_Ex1$TEST=="ADD",c(1:4,6:8)])
  colnames(test.sumstats) <- c("chr","rsid","pos","a1","n_eff","beta","beta_se") summarystat
  = test.sumstats ncores = 1 byCHR = TRUE r2_LD = 0 LDSC_blocks = 20 REMLalgo = 0 nitr = 100
  cat_covarfile = NULL quant_covarfile = NULL prevalance = 0.01 partGRM = FALSE autosome =
  TRUE Xsome = TRUE nGRM = 3 cripticut = 0.025 minMAF = NULL maxMAF = NULL hg = "hg19"
  PlotIndepSNP = TRUE
  IndepSNP_window_size = 50
  IndepSNP_step_size = 5

  IndepSNP_r2_threshold = 0.02
  highLD_regions = highLD_hg19

  H2 <- EstimateHerit(DataDir = DataDir, ResultDir = ResultDir, finput = finput, summarystat = summarystat, ncores =
             ncores, model = model, byCHR = byCHR, r2_LD = r2_LD, LDSC_blocks =
             LDSC_blocks,REMLalgo = REMLalgo, nitr = nitr, cat_covarfile = cat_covarfile, quant_covarfile =
             quant_covarfile,prevalance = prevalance, partGRM = partGRM, autosome = autosome, Xsome = Xsome, nGRM =
             nGRM,cripticut = cripticut, minMAF = minMAF, maxMAF = maxMAF, hg = hg,
             PlotIndepSNP = PlotIndepSNP, IndepSNP_window_size = IndepSNP_window_size, IndepSNP_step_size =
             IndepSNP_step_size, IndepSNP_r2_threshold = IndepSNP_r2_threshold, highLD_regions = highLD_hg19)
```

#### [1] "Program is set up."
#### 1,478 variants to be matched.

#### 237 ambiguous SNPs have been removed.
#### 1,241 variants have been matched; 0 were flipped and 0 were reversed.
#### [1] "Processing chromosome 1"
#### [1] "Processing chromosome 2"
#### [1] "Processing chromosome 3"
#### [1] "Summary statistics doesn't contain all the chromosomes."
#### [1] "Processing chromosome 4"
#### [1] "Summary statistics doesn't contain all the chromosomes."
#### [1] "Processing chromosome 5"
#### [1] "Summary statistics doesn't contain all the chromosomes."
#### [1] "Processing chromosome 6"
#### [1] "Summary statistics doesn't contain all the chromosomes."
#### [1] "Processing chromosome 7"
#### [1] "Summary statistics doesn't contain all the chromosomes."
#### [1] "Processing chromosome 8"
#### [1] "Summary statistics doesn't contain all the chromosomes."
#### [1] "Processing chromosome 9"
#### [1] "Summary statistics doesn't contain all the chromosomes."
#### [1] "Processing chromosome 10"
#### [1] "Summary statistics doesn't contain all the chromosomes."
#### [1] "Processing chromosome 23"
#### [1] "Summary statistics doesn't contain all the chromosomes."
#### [1] "Processing chromosome 24"
#### [1] "Summary statistics doesn't contain all the chromosomes."
#### [1] "Not enough data points for plots."

Table 42: Per chromosome heritability estimates for the example genotype datasets using the LDSC model. Chromosome 2 showing a higher proportion of phenotypic variance explained (V(G)/Vp_L = 0.88) compared to chromosome 1 (V(G)/Vp_L = 0.58), despite having fewer genes and proteins.

| Chromosome | snp_proportion | no.of.genes | no.of.proteins | Intercept | Int_SE | Varianc | SE | Source |
|---|---|---|---|---|---|---|---|---|
| 1 | 0.0277 | 2175 | 1936 | 1.02263 | 0.05950 | NA | 0.58 | V(G)/Vp_L |
| 2 | 0.0275 | 1375 | 1188 | 0.9388 | 0.05845 | NA | 0.88 | V(G)/Vp_L |

#### Run EstimateHerit() with model = 'LDSC' genome-wide

Users can run this model by computing the GRM per chromosome or they can use pre-computed GRMs by setting the parameter ComputeGRM <- TRUE or FALSE.

In this example, we set this to 'TRUE'.

```
model <- "LDSC"
data("GXwasRData")
DataDir = system.file("extdata", package = "GXwasR")
ResultDir = tempdir()
finput <- "GXwasR_example"
```

```
test.sumstats <- na.omit(Summary_Stat_Ex1[Summary_Stat_Ex1$TEST=="ADD",c(1:4,6:8)])
colnames(test.sumstats) <- c("chr","rsid","pos","a1","n_eff","beta","beta_se")
summarystat = test.sumstats
highLD_regions = highLD_hg19
H2 <- EstimateHerit(DataDir = DataDir, ResultDir = ResultDir, finput = finput, summarystat = summarystat, ncores =
            ncores, model = model, byCHR = byCHR, r2_LD = r2_LD, LDSC_blocks =
            LDSC_blocks,REMLalgo = REMLalgo, nitr = nitr, cat_covarfile = cat_covarfile, quant_covarfile =
            quant_covarfile,prevalance = prevalance, partGRM = partGRM, autosome = autosome, Xsome = Xsome, nGRM =
            nGRM,cripticut = cripticut, minMAF = minMAF, maxMAF = maxMAF, hg = hg,
            PlotIndepSNP = PlotIndepSNP, IndepSNP_window_size = IndepSNP_window_size, IndepSNP_step_size =
            IndepSNP_step_size, IndepSNP_r2_threshold = IndepSNP_r2_threshold, highLD_regions = highLD_hg19
```

Table 43: Genome-wide heritability estimate for the example genotype dataset using the LDSC model.

| Source | Variance | SE | Intercept | Int_SE |
|---|---|---|---|---|
| V(G)/Vp | -1.3083494 | 0.6461225 | 0.965281 | 0.0345606 |
| V(G)/Vp_L | -0.7220876 | 0.6461225 | 0.965281 | 0.0345606 |

Tutorial for computing genetic correlation between two traits

Users can run this model by computing the GRM per chromosome or they can use pre-computed GRMs by setting the parameter ComputeGRM <- TRUE or FALSE.

In this example, we set this to ‘TRUE.

```
# Running’

DataDir <- system.file("extdata", package = "GXwasR")

ResultDir <- tempdir()

finput <- "GXwasR_example"

byCHR = FALSE

REMLalgo = 0

nitr <- 3

ncores <- 3

phenofile <- Example_phenofile #Cannot be NULL, the interested phenotype column should be labeled as

cat_covarfile <- NULL

quant_covarfile <- NULL

partGRM <- FALSE #Partition the GRM into m parts (by row)

autosome = TRUE

Xsome <- TRUE

cripticut = 0.025

minMAF <- 0.01 # if MAF filter apply

maxMAF <- 0.04

excludeResidual = TRUE

GC <- GeneticCorrBT(DataDir = DataDir, ResultDir = ResultDir, finput = finput, byCHR = byCHR,
```

```
REMLalgo = 0, nitr = nitr, phenofile = phenofile, cat_covarfile = NULL, quant_covarfile = NULL,partGRM =FALSE,
        autosome = TRUE, Xsome = TRUE, nGRM = 3,cripticut =
        0.025, minMAF = NULL, maxMAF = NULL,excludeResidual = TRUE, ncores = ncores)
```

```
## [1] "GCTA setup complete. This program is currently only supported in Linux."
## [1] "Error: Log-likelihood not converged (stop after 3 iteractions). "
## [1] "Note: to constrain the correlation being from -1 to 1, a genetic (or residual) variancecovariance matrix is
bended to be positive definite. In this case, the SE is unreliable."
## [1] "Convergence problem occurs, please try byCHR = TRUE, check different options, SNP partitioning or ensure the
quality of the input data."
## [1] "The result will be provided for the last iteration."
##Genetic correlation from the last iteration in this example there was no rg value since the program stopped after nitr-1
= 2 iteration due to convergence issue.
```

Table 44: Result from the last iteration of genetic correlation estimate between two traits using example datasets provided in with the tool GXwasR.

| Source | Variance |
|---|---|
| logL | 1504.20 |
| V(G1)_tr1 | -0.08685 |
| V(G1)_tr2 | -0.07878 |
| C(G1)_tr12 | 0.02083 |
| V(G2)_tr1 | 0.12472 |
| V(G2)_tr2 | 0.01038 |
| C(G2)_tr12 | 0.00332 |
| V(e)_tr1 | 0.21145 |
| V(e)_tr2 | 0.04887 |

### Tutorial for performing GWAS meta-analysis Example Datasets

Example datasets are provided with the package and can be accessed by calling data(“GXwasRData”).

Load the example datasets to perform this tutorial.

```
## Load Library
library(GXwasR)
data("GXwasRData")
```

1. Two sets of summary GWAS summary statistics in .Rda files will be required to perform this tutorial. **Summary_Stat_Ex1.Rda Summary_Stat_Ex2.Rda**

Check one set of the summary statistics

```r
## Visualize three rows and all the columns
Summary_Stat_Ex1[1:3,]
```

```
##   CHR         SNP     BP A1 TEST NMISS    BETA     SE     L95   U95    STAT
## 1   1 rs143773730  73841  T  ADD   125 -0.0789 0.2643 -0.5968 0.439 -0.2986
## 4   1 rs147281566 775125  T  ADD   125 -0.3959 1.2380 -2.8230 2.031 -0.3197
## 6   1  rs35854196 863863  A  ADD   125  1.0500 0.8858 -0.6864 2.786  1.1850
##        P
## 1 0.7653
## 4 0.7492
## 6 0.2360
```

Of the 12 columns, several are mandatory for this tutorial: 'SNP' (i.e., SNP identifier), 'BETA' (i.e., effect-size or logarithm of odds ratio), 'SE' (i.e., standard error of BETA), 'P' (i.e., p-values) and 'NMISS' (i.e., effective sample size). The other columns are, 'CHR' (i.e., chromosome), 'BP' (i.e., basepair position), A1 (i.e., risk allele), TEST (i.e., association test type), L95 (i.e., lower limit of 95 percentile confidence interval), U95 (i.e., upper limit of 95 percentile confidence interval) and STAT (i.e., test statistic).

2. **UniqueLoci.Rda:** .Rda file with a single column containing SNP names. These could be LD clumped SNPs or any other list of SNPs selected for meta-analysis.
3. **SNPsPlot.Rda:** .Rda file with a single column containing SNP names for the forest plots.

#### Run MetaGWAS

```r
DataDir = system.file("extdata", package = "GXwasR")
ResultDir = tempdir()
SummData <- list(Summary_Stat_Ex1,Summary_Stat_Ex2)
SNPfile = "UniqueLoci"
useSNPposition = FALSE
UseA1 = TRUE
GCse = TRUE
byCHR = FALSE
pval_filter = "R"
top_snp_pval = 1e-08
max_top_snps = 10
chosen_snps_file = NULL
pval_threshold_manplot = 1e-05
```

```r
plotname = "Meta_Analysis.plot"

x <- MetaGWAS(DataDir = DataDir, SummData = SummData,ResultDir=ResultDir, SNPfile = NULL,
         useSNPposition = TRUE, UseA1 = UseA1,GCse = GCse, plotname = "Meta_Analysis.plot",
         pval_filter, top_snp_pval, max_top_snps, chosen_snps_file = NULL, byCHR,
         pval_threshold_manplot)
```

```
## [1] "Processing file number 1"
## [1] "Processing file number 2"
## [1] "Applying study-specific genomic control."
## [1] "Applying study-specific genomic control."
## [1] "Program is set up."
## [1] "Processing chromosome "
```

```
## [1] "Meta_Analysis.plot files containing the forest plots of the SNPs are produced in the directory
C:\\Users\\AppData\\Local\\Temp\\RtmpucHg2F."
```

```
## [1] "Meta_Analysis.plot files containing the forest plots of the SNPs are produced in the directory
C:\\Users\\AppData\\Local\\Temp\\RtmpucHg2F."
```

#### Random effect meta-analysis result

```r
# Dataframe with the fixed effect result
x1 <- x$Resultrandom
x2 <- x1[order(x1$P),]
knitr::kable(x2[1:10,], caption = 'Top ten associations from random effect model.')
```

Table 45: Top ten associations from the random effects meta-analysis model.

| CHR | BP | SNP | A1 | A2 | Q | I | P | ES | SE | CI_L | CI_U |
|---|---|---|---|---|---|---|---|---|---|---|---|
| 23 | 4184349 | rs6529954 | A | ? | 0.1162 | 59.48 | 0.0000002 | -1.7980 | 0.3441459 | -2.472526 | -1.1234740 |
| 23 | 4137114 | rs5962098 | A | ? | 0.1753 | 45.56 | 0.0000010 | 1.1488 | 0.2351018 | 0.6880004 | 1.6095996 |
| 23 | 3376304 | rs6420571 | A | ? | 0.3068 | 4.26 | 0.0000015 | -1.1667 | 0.2426946 | -1.642381 | -0.6910186 |
| 23 | 4214861 | rs12858640 | C | ? | 0.0725 | 69.00 | 0.0000062 | -1.7572 | 0.3887229 | -2.519096 | -0.9953032 |
| 2 | 2579014 | rs10186455 | G | ? | 0.4490 | 0.00 | 0.0000476 | 1.2694 | 0.3121061 | 0.6576721 | 1.8811279 |
| 23 | 4119808 | rs10521557 | A | ? | 0.8021 | 0.00 | 0.0000631 | 1.4884 | 0.3720121 | 0.7592563 | 2.2175437 |
| 1 | 1127860 | rs148527527 | G | ? | 0.5778 | 0.00 | 0.0008237 | 1.3978 | 0.4179142 | 0.5786881 | 2.2169119 |
| 2 | 2535670 | rs13430614 | C | ? | 0.6681 | 0.00 | 0.0010230 | 0.6358 | 0.1935981 | 0.2563478 | 1.0152522 |
| 23 | 45563626 | rs1207312 | T | ? | 0.5388 | 0.00 | 0.0046670 | 0.6138 | 0.2169547 | 0.1885689 | 1.0390311 |
| 23 | 45565805 | rs1780835 | A | ? | 0.5206 | 0.00 | 0.0064620 | 0.5404 | 0.1984308 | 0.1514757 | 0.9293243 |

#### Fixed effect meta-analysis result

```r
# Dataframe with the fixed effect result
x1 <- x$Resultfixed
x2 <- x1[order(x1$P),]
knitr::kable(x2[1:10,], caption = 'Top ten associations from fixed effect model.')
```

Table 46: Top ten associations from the fixed effect meta-analysis model.

| CHR | BP | SNP | A1 | A2 | Q | I | P | ES | SE | CI_L | CI_U |
|---|---|---|---|---|---|---|---|---|---|---|---|
| 23 | 4184349 | rs6529954 | A | ? | 0.1162 | 59.48 | 0.0000000 | -1.712 | 0.2025697 | -2.109736 | -1.3156635 |
| 23 | 4214861 | rs12858640 | C | ? | 0.0725 | 69.00 | 0.0000000 | -1.636 | 0.1955735 | -2.019724 | -1.2530760 |
| 23 | 4137114 | rs5962098 | A | ? | 0.1753 | 45.56 | 0.0000000 | 1.1030 | 0.1618941 | 0.7856876 | 1.4203124 |
| 23 | 3376304 | rs6420571 | A | ? | 0.3068 | 4.26 | 0.0000009 | -1.164 | 0.2371161 | -1.629447 | -0.6999524 |
| 2 | 2579014 | rs10186455 | G | ? | 0.4490 | 0.00 | 0.0000476 | 1.2694 | 0.3121061 | 0.6576721 | 1.8811279 |
| 23 | 4119808 | rs10521557 | A | ? | 0.8021 | 0.00 | 0.0000631 | 1.4884 | 0.3720121 | 0.7592563 | 2.2175437 |
| 1 | 1127860 | rs148527527 | G | ? | 0.5778 | 0.00 | 0.0008237 | 1.3978 | 0.4179142 | 0.5786881 | 2.2169119 |
| 2 | 2535670 | rs13430614 | C | ? | 0.6681 | 0.00 | 0.0010230 | 0.6358 | 0.1935981 | 0.2563478 | 1.0152522 |
| 2 | 8181194 | rs7370955 | A | ? | 0.2403 | 27.48 | 0.0024130 | 0.7752 | 0.2555007 | 0.2744187 | 1.2759813 |

#### Weighted effect meta-analysis result

```
# Dataframe with the fixed effect result
x1 <- x$Resultweighted
x2 <- x1[order(x1$P),]
knitr::kable(x2[1:10,], caption = 'Top ten associations from weighted effect model.')
```

Table 47: Top ten associations from the weighted effect model.

| CHR | BP | SNP | A1 | A2 | Q | I | P | ES | SE | CI_L | CI_U | CHR |
|---|---|---|---|---|---|---|---|---|---|---|---|---|
| 1054 | 23 | 4184349 | rs6529954 | A | ? | 0.1162 | 59.48 | 0.00000 | -8.340 | 0.0000000 | -8.3400000 | -8.340000 |
| 1055 | 23 | 4214861 | rs12858640 | C | ? | 0.0725 | 69.00 | 0.00000 | -8.306 | 0.0000000 | -8.3060000 | -8.306000 |
| 1053 | 23 | 4137114 | rs5962098 | A | ? | 0.1753 | 45.56 | 0.00000 | 6.741 | 1.0000820 | 4.7808392 | 8.701161 |
| 1043 | 23 | 3376304 | rs6420571 | A | ? | 0.3068 | 4.26 | 0.00001 | -4.883 | 1.0000062 | -6.8430122 | -2.922988 |
| 559 | 2 | 2579014 | rs10186455 | G | ? | 0.4490 | 0.00 | 0.00007 | 3.969 | 1.0000874 | 2.0088287 | 5.929171 |
| 1052 | 23 | 4119808 | rs10521557 | A | ? | 0.8021 | 0.00 | 0.00009 | 3.902 | 1.0000837 | 1.9418359 | 5.862164 |
| 6 | 1 | 1127860 | rs148527527 | G | ? | 0.5778 | 0.00 | 0.00107 | 3.270 | 0.9999618 | 1.3100749 | 5.229925 |
| 558 | 2 | 2535670 | rs13430614 | C | ? | 0.6681 | 0.00 | 0.00142 | 3.189 | 0.9998946 | 1.2292065 | 5.148794 |
| 638 | 2 | 8181194 | rs7370955 | A | ? | 0.2403 | 27.48 | 0.00324 | 2.944 | 1.0000649 | 0.9838728 | 4.904127 |
| 1382 | 23 | 45563626 | rs1207312 | T | ? | 0.5388 | 0.00 | 0.00483 | 2.818 | 1.0000617 | 0.8578790 | 4.778121 |

#### Summary of the meta-analysis

```
# Dataframe with the metadata
x1 <- x$Metadata
x2 <- x1[order(x1$Q),]
knitr::kable(x2[1:10,], caption = 'Metadata of the top ten associations based on Cochrane’s Q
         statistics.')
```

Table 48: Metadata of the top ten heterogeneous associations based on Cochrane’s Q statistic. Higher Q values indicate greater heterogeneity in effect sizes across subgroups or strata.

| CHR | BP | SNP | A1 | A2 | N | Q | I | F0 | F1 |
|---|---|---|---|---|---|---|---|---|---|
| 2 | 21385538 | rs575905 | T | ? | 2 | 0.0010 | 90.73 | -0.5218 | 0.6472 |
| 23 | 9277424 | rs2214279 | G | ? | 2 | 0.0015 | 90.09 | 0.5001 | -0.5365 |
| 2 | 9913645 | rs1106144 | A | ? | 2 | 0.0020 | 89.48 | 0.5909 | -0.8972 |
| 2 | 10210922 | rs11678624 | T | ? | 2 | 0.0021 | 89.45 | -0.3659 | 0.7374 |
| 23 | 23909933 | rs2428144 | A | ? | 2 | 0.0031 | 88.55 | 0.5344 | -0.4081 |
| 23 | 29189630 | rs225456 | C | ? | 2 | 0.0031 | 88.56 | 0.3208 | -1.1170 |
| 23 | 28763567 | rs12008039 | G | ? | 2 | 0.0032 | 88.48 | -0.3187 | 0.5565 |
| 2 | 21275825 | rs12468735 | A | ? | 2 | 0.0036 | 88.19 | -0.5095 | 0.5175 |
| 2 | 10343419 | rs759347 | C | ? | 2 | 0.0041 | 87.88 | 0.9790 | -0.3894 |
| 23 | 29104124 | rs16988439 | C | ? | 2 | 0.0042 | 87.81 | 0.3208 | -1.0730 |

```
# Dataframe with the problematic SNPs
x1 <- x$ProblemSNP
knitr::kable(x1[1:10,], caption = 'Top ten probematic SNPs.')
```

Table 49: Ten problematic SNPs identified during meta-analysis, ranked based on the frequency or severity of allele mismatches across datasets. These SNPs exhibit inconsistencies between reported alleles across contributing studies, flagged as ALLELE_MISMATCH.

| File | SNP | Problem |
|---|---|---|
| C:2F/SNPdata_2 | rs2803333 | ALLELE_MISMATCH |
| C:2F/SNPdata_2 | rs672606 | ALLELE_MISMATCH |
| C:2F/SNPdata_2 | rs7519955 | ALLELE_MISMATCH |
| C:2F/SNPdata_2 | rs1538466 | ALLELE_MISMATCH |
| C:2F/SNPdata_2 | rs61777960 | ALLELE_MISMATCH |
| C:2F/SNPdata_2 | rs2865211 | ALLELE_MISMATCH |
| C:2F/SNPdata_2 | rs10917217 | ALLELE_MISMATCH |
| C:2F/SNPdata_2 | rs196402 | ALLELE_MISMATCH |
| C:2F/SNPdata_2 | rs6662038 | ALLELE_MISMATCH |
| C:2F/SNPdata_2 | rs3845484 | ALLELE_MISMATCH |

Users should review these variants carefully and consider excluding them or resolving the mismatches (e.g., through all harmonization) before re-running the meta-analysis to avoid inflated heterogeneity or spurious associations.

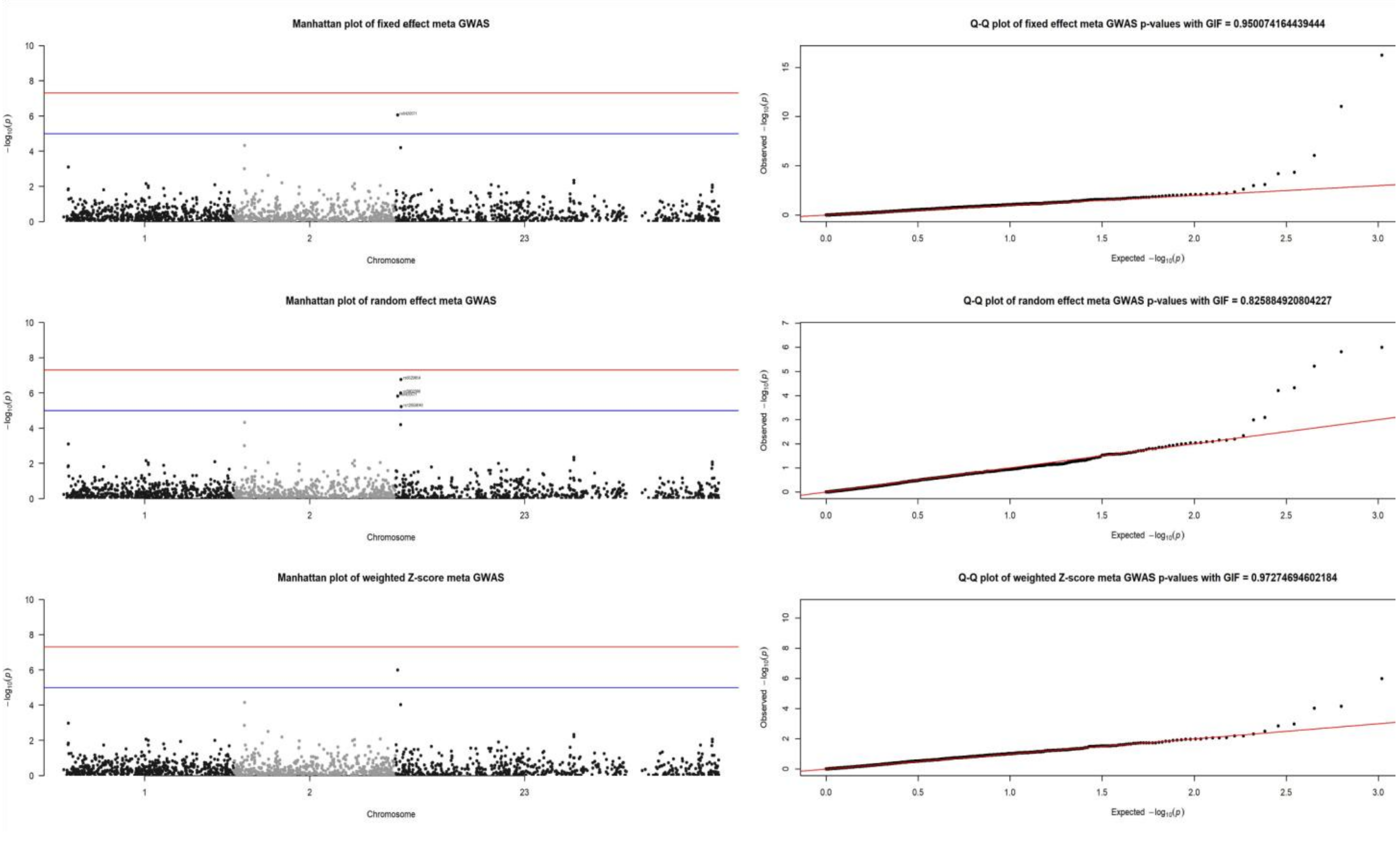

Figure 19 : Manhattan and QQ plots from the meta-analysis from fixed, random and weighted-effect meta-analysis models.

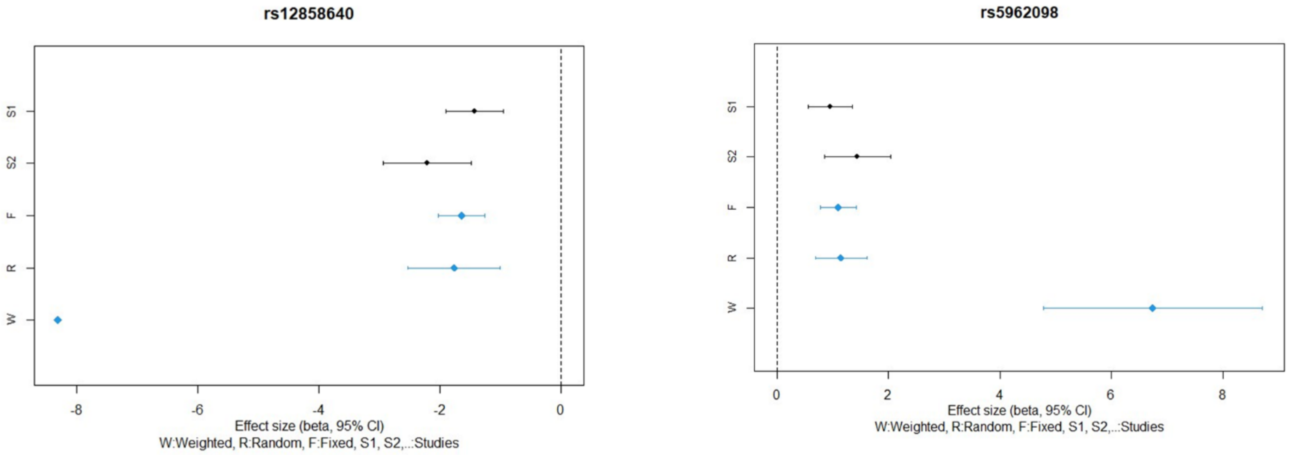

Figure 20 : Forest plot of the two SNPs from the meta-analysis from fixed, random and weighted-effect models.

### Supplementary Method

#### Preparation of phenotype data for BioVu samples

Clinical laboratory values and routine clinical measurements were extracted from the SD using the established pipeline developed for this purpose, QualityLabs[34], with some updates. First, clinical measurements were standardized into the most common unit of measurement using the Unified Codes of Units of Measurement software library from the National Library of Medicine (UCUM-LHC). Clinical measurements were restricted to numeric, non-infinite values from adult (taken at age$\geq$18 years) males and females. Stratified by sex, we filtered observations outside of 4 standard deviations from the sample mean, calculated the median lab value for each patient and extracted the patient's age at median lab value. Separately for males and females, median values were residualized for 3 splines of median age, and then inverse normal quantile transformed to account for skewness and non-normality.

#### Preparation of genotype data using GXwasR

We used genotype data from 72,824 BioVU individuals with genetic data which clustered with European reference populations, and who were genotyped on the Illumina MEGAEX array. Imputation was completed using the Michigan Imputation Server using the Haplotype Reference Consortium (HRC) reference panel separately for the autosomes and the X chromosome. The Michigan Imputation Sever does not provide imputation of the pseudoautosomal regions of the X chromosomes. SNPs were then filtered for SNP imputation quality ($R^2 > 0.3$) and converted to hard calls. We performed sex-aware quality control using GXwasR (**Supplementary Figure 21**), separately in the autosomes and the X chromosome and in males and females. Namely, we filtered biallelic variants with MAF < 0.05, or missing call rate > 0.05. We performed a sex-stratified sample filter, removing individuals with missingness > 0.10 or outside of > 3 SD heterozygosity. This yielded a dataset of 72,131 individuals and 3,359,461 variants.

For genome-wide and X-chromosome association analyses, we also removed related individuals, using a pi hat filter of 0.2 (11,754 individuals removed, 60,377 remaining). After restricting to those who had adult height measurements, there were a total of 50,302 individuals. Principal components were calculated within the GXwas R package using the function *ComputeGeneticPC()*, and options to remove regions of high LD and perform LD pruning (window size=80, step size=8, r2 threhsold=0.15).

Heritability was estimated with *EstimateHerit()*, using the GREML model with computed genetic relationship matrices (GRM) derived from BioVU genotype data. We also use the LDSC model implemented in *EstimateHerit()*, using the summary statistics from the GWAS and XWAS with precomputed LD scores derived from UK Biobank genotype data[16].

To assess genetic correlation, we used the reference UKB_imputed_SVD_eigen99_extraction with default parameters of the *SumstatGenCorr()* function.

#### Application of GXwasR function with BioVU data

When running the *GXwas()* function in the package, we modified the original code from the default settings to tailor the analysis pipeline to our dataset. Specifically, we made the following changes in the input arguments:

1. *trait* = "quantitative"
2. *standard_beta* = "FALSE"
3. *xmodel* = "FMstratified"
4. *sex* = "TRUE"
5. *xsex* = "TRUE"
6. *MF.p.core* = "bonferroni"

When running heritability using , *EstimateHerit()*, we modified the default setting as:

1. ncores = 1
2. byCHR = TRUE for GREML, also ran with it set to FALSE
3. REMLalgo = 0
4. cripticut = 0.05
5. hg = hg19
6. PlotIndepSNP = FALSE except for the chromosome-level heritability

We clumped the sex-stratified GWAS results using the *ClumpLD()* function and the following parameters: clump_kb=250, clump_p1=5e-8, clump_p2=0.01, clump_r2=0.5. We restricted the *SexDiff*() function to SNPs that were only nominally significant in males or females ($p < 0.05$), resulting in 474,680 variants that underwent sex-differential testing. We then ran the *SexDiff*() function without altering the default settings.

#### Supplementary Figures from BioVU data analysis

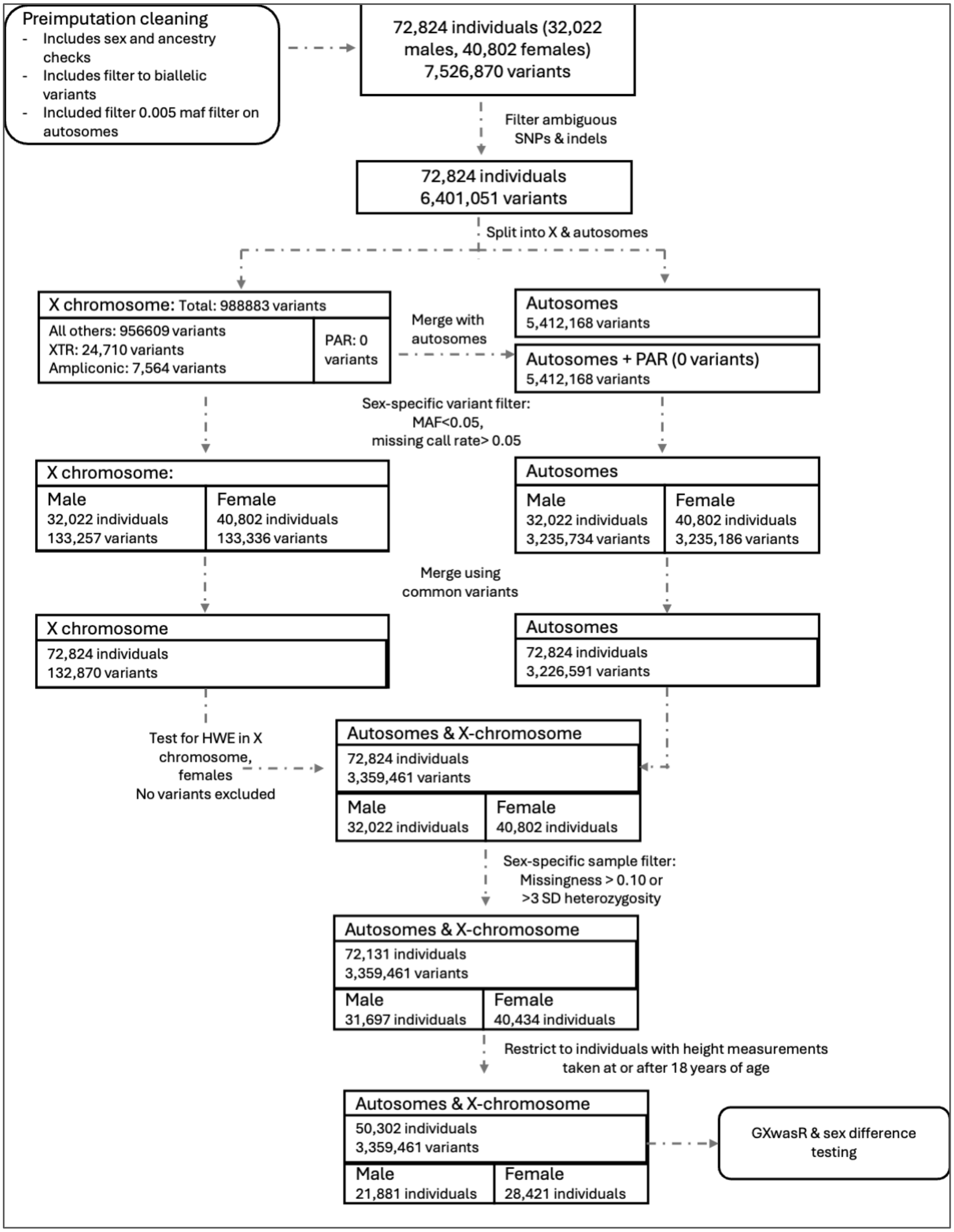

Figure 21: Post-imputation QC pipeline of BioVU, the VUMC genotype dataset using GXwasR.

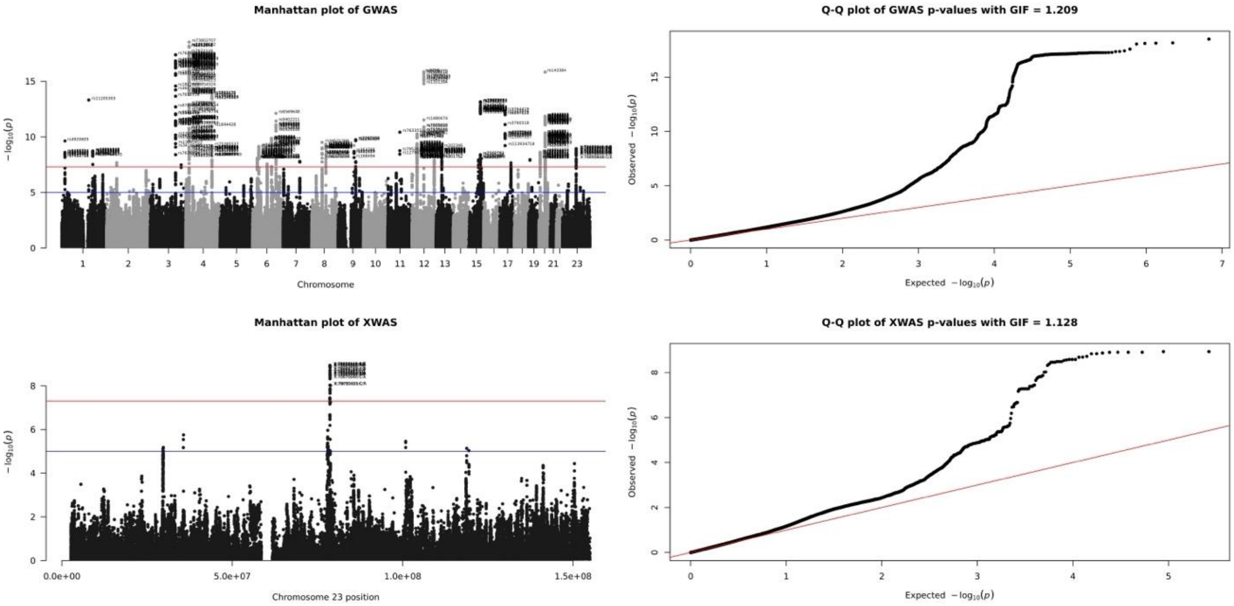

Figure 22. Manhattan and Q-Q plots of GWAS and XWAS results for height in females using the *GXwas*() function in BioVU data

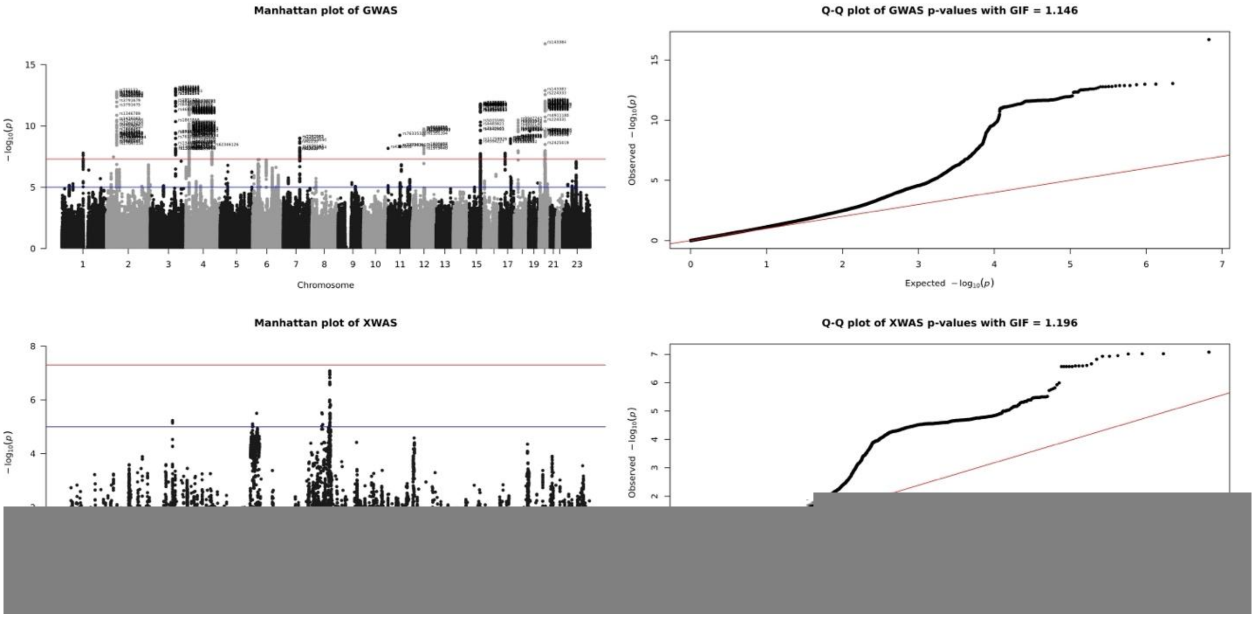

Figure 23. Manhattan and Q-Q plots of GWAS and XWAS results for height in males using the *GXwas*() function in BioVU data.

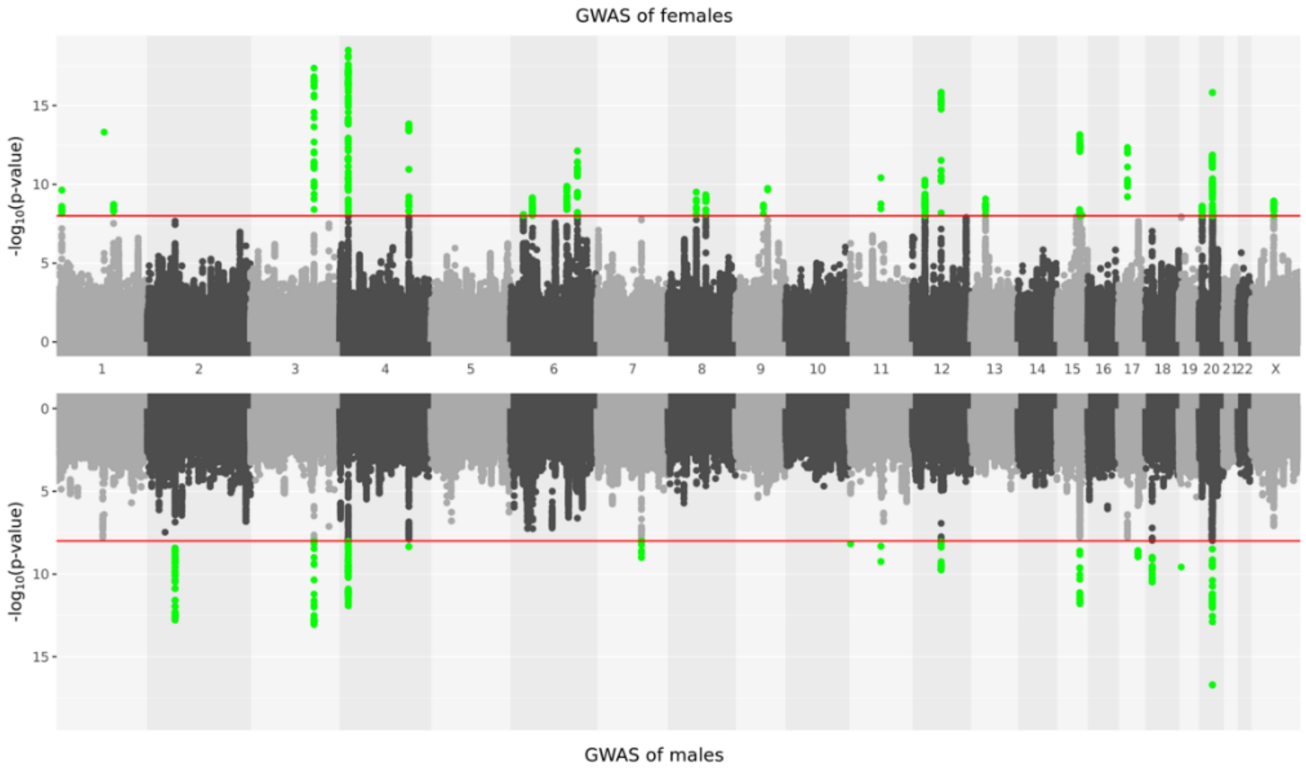

Figure 24. Miami plot of sex-stratified GWAS results for height in males and females using the GXwas() function in BioVU data.

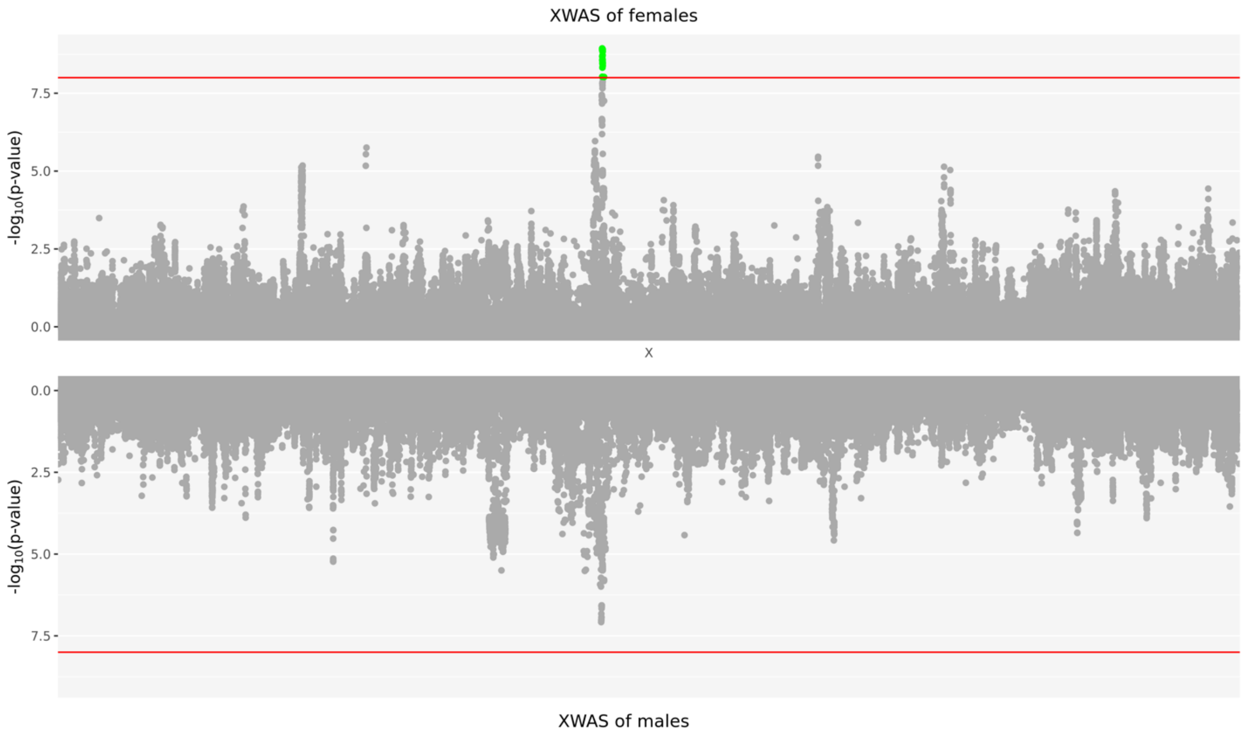

Figure 25. Miami plot of sex-stratified XWAS results for height in males and females generated by using the GXwas() package with BioVU data.

Table 50: Table X. Genome-wide significant loci associated with height from sex-stratified analysis. This table lists 71 independent loci identified using the ClumpLD() function, each represented by the lead SNP with genome-wide significance ($p < 5\times10^{-8}$) in males, females, or both. Columns include chromosome (CHR), SNP ID, base-pair position (BP), gene symbol, sex in which the variant was clumped, sex-specific effect sizes (beta and standard error), corresponding p-values, and overlap with lead or clumped variants in the EMBL dataset.

| CHR | SNP | BP | Gene_Symbol | Clumped variant from sex | Female beta (SE) | female_P | Male beta (SE) | male_P | Lead variant in EMBL | Any clumped variant in EMBL |
|---|---|---|---|---|---|---|---|---|---|---|
| 1 | rs4920605 | 17315425 | ATP13A2 | Both | -0.05197 (0.008197) | 2.34E-10 | -0.0284 (0.009372) | 0.002447 | FALSE | TRUE |
| 1 | rs59051938 | 118850501 | RNA5SP56 | Both | -0.02587 (0.009454) | 0.00621 | -0.06054 (0.01072) | 1.644E-08 | FALSE | TRUE |
| 1 | rs11205303 | 149906413 | MTMR11 | Both | 0.06232 (0.008266) | 4.86E-14 | 0.04778 (0.00942) | 3.966E-07 | TRUE | TRUE |
| 1 | rs10911338 | 172197896 | DNM3 | Both | 0.05961 (0.009917) | 1.87E-09 | 0.04537 (0.01124) | 0.00005459 | FALSE | TRUE |
| 2 | rs12615742 | 37995727 | AC006369.3 | Male only | 0.0165 (0.008205) | 0.04437 | 0.05133 (0.009292) | 3.359E-08 | TRUE | TRUE |
| 2 | rs732132 | 56011881 | EFEMP1 | Both | -0.03858 (0.009173) | 2.61E-05 | -0.07768 (0.01053) | 1.659E-13 | FALSE | TRUE |
| 2 | rs1346786 | 56108333 | EFEMP1 | Both | -0.03492 (0.008788) | 7.08E-05 | -0.06289 (0.01008) | 4.512E-10 | TRUE | TRUE |
| 3 | rs13100711 | 141082990 | ZBTB38 | Both | 0.05785 (0.008864) | 6.85E-11 | 0.05129 (0.01004) | 3.274E-07 | FALSE | TRUE |
| 3 | rs7624084 | 141093285 | ZBTB38 | Both | 0.07138 (0.008225) | 4.23E-18 | 0.06963 (0.00937) | 1.117E-13 | FALSE | TRUE |
| 3 | rs7636914 | 141158614 | ZBTB38 | Both | 0.05129 (0.008714) | 4E-09 | 0.03717 (0.009895) | 0.0001725 | FALSE | TRUE |
| 3 | rs13094143 | 171973787 | FNDC3B | Both | 0.04807 (0.008698) | 3.31E-08 | 0.05308 (0.009869) | 7.586E-08 | FALSE | TRUE |
| 4 | rs740672 | 17782264 | FAM184B | Both | -0.05537 (0.009691) | 1.12E-08 | -0.04761 (0.01084) | 0.00001126 | TRUE | TRUE |
| 4 | rs73802707 | 17932319 | LCORL | Both | -0.0998 (0.01112) | 3.07E-19 | -0.08623 (0.01271) | 1.187E-11 | FALSE | TRUE |
| 4 | rs2061456 | 17998426 | LCORL | Both | 0.06543 (0.009312) | 2.16E-12 | 0.06151 (0.01061) | 6.926E-09 | TRUE | TRUE |
| 4 | rs2169033 | 18044357 | LCORL | Both | 0.05259 (0.008825) | 2.57E-09 | 0.05147 (0.01005) | 3.033E-07 | FALSE | FALSE |
| 4 | rs17776795 | 145544836 | HHIP-AS1 | Both | 0.04968 (0.008156) | 1.13E-09 | 0.05024 (0.009309) | 6.854E-08 | FALSE | TRUE |
| 4 | rs1812175 | 145574844 | HHIP | Both | -0.08415 (0.01094) | 1.47E-14 | -0.06677 (0.01256) | 1.074E-07 | TRUE | TRUE |
| 4 | rs7666450 | 145662842 | HHIP | Both | 0.04514 (0.00815) | 3.07E-08 | 0.0288 (0.009239) | 0.001827 | FALSE | TRUE |
| 4 | rs75385537 | 145706070 | HHIP | Both | -0.08499 (0.01374) | 6.27E-10 | -0.05805 (0.01558) | 0.0001958 | FALSE | FALSE |
| 6 | rs806794 | 26200677 | HIST1H2BF | Both | -0.05094 (0.008993) | 1.49E-08 | -0.04292 (0.01022) | 0.00002701 | TRUE | TRUE |
| 6 | rs12660432 | 26317384 | HIST1H3PS1 | Both | -0.04931 (0.008555) | 8.29E-09 | -0.02148 (0.009684) | 0.02653 | FALSE | TRUE |
| 6 | rs112540634 | 34623905 | C6orf106 | Both | 0.06994 (0.01135) | 7.25E-10 | 0.05624 (0.01291) | 0.00001337 | TRUE | TRUE |
| 6 | rs1821780 | 81100747 | RPL17P25 | Both | 0.09972 (0.01797) | 2.89E-08 | 0.03858 (0.02075) | 0.06294 | FALSE | TRUE |
| 6 | rs78892181 | 81305223 | RP11-486E2.1 | Both | 0.07352 (0.01322) | 2.71E-08 | 0.03932 (0.01523) | 0.00982 | FALSE | TRUE |
| 6 | rs9391254 | 105377347 | LINC00577 | Both | 0.05582 (0.008686) | 1.33E-10 | 0.03046 (0.009904) | 0.002105 | FALSE | TRUE |
| 6 | rs6569648 | 130349119 | L3MBTL3 | Both | 0.06883 (0.0096) | 7.67E-13 | 0.05643 (0.01092) | 2.393E-07 | TRUE | TRUE |
| 7 | rs4272 | 92236829 | CDK6 | Both | 0.05605 (0.009921) | 1.63E-08 | 0.05353 (0.01121) | 0.000001815 | FALSE | FALSE |
| 7 | rs2282983 | 92279363 | CDK6 | Both | 0.03141 (0.008495) | 0.000219 | 0.05901 (0.009652) | 9.851E-10 | FALSE | TRUE |
| 8 | rs62515430 | 57109358 | PLAG1 | Both | 0.06189 (0.01012) | 9.64E-10 | 0.03356 (0.01139) | 0.003217 | TRUE | TRUE |
| 8 | rs34571768 | 57112046 | PLAG1 | Both | -0.07589 (0.01206) | 3.16E-10 | -0.04179 (0.01379) | 0.002452 | TRUE | TRUE |

| | | | | | | | | | | |
|---|---|---|---|---|---|---|---|---|---|---|
| 8 | rs7846385 | 78160179 | AC105242.1 | Both | 0.05611 (0.009003) | 4.67E-10 | 0.04057 (0.01024) | 0.00007418 | TRUE | TRUE |
| 9 | rs353785 | 89099362 | RNU2-36P | Both | -0.04873 (0.008127) | 2.04E-09 | -0.03813 (0.009265) | 0.00003873 | FALSE | FALSE |
| 9 | rs4385527 | 97648587 | C9orf3 | Female only | 0.04518 (0.008271) | 4.74E-08 | 0.02232 (0.009386) | 0.0174 | FALSE | FALSE |
| 9 | rs2297086 | 98240120 | PTCH1 | Female only | 0.05414 (0.008486) | 1.8E-10 | 0.01057 (0.009667) | 0.2744 | FALSE | TRUE |
| 11 | rs4320932 | 2171601 | INS-IGF2 | Both | -0.05137 (0.01027) | 5.67E-07 | -0.06773 (0.01168) | 6.709E-09 | TRUE | TRUE |
| 11 | rs76335321 | 67097778 | SSH3 | Both | -0.09857 (0.0149) | 3.81E-11 | -0.1053 (0.01699) | 5.722E-10 | FALSE | TRUE |
| 12 | rs10843114 | 28303296 | CCDC91 | Both | -0.05704 (0.008695) | 5.49E-11 | -0.02111 (0.009881) | 0.03268 | FALSE | TRUE |
| 12 | rs2272361 | 28602970 | CCDC91 | Both | -0.0546 (0.008735) | 4.14E-10 | -0.02495 (0.009983) | 0.01244 | FALSE | TRUE |
| 12 | rs8756 | 66359752 | HMGA2 | Both | 0.06756 (0.008175) | 1.46E-16 | 0.0577 (0.009209) | 3.789E-10 | TRUE | TRUE |
| 12 | rs61921611 | 66367726 | HMGA2 | Both | -0.05096 (0.008782) | 6.6E-09 | -0.04381 (0.00998) | 0.00001143 | TRUE | TRUE |
| 12 | rs1790120 | 123613565 | PITPNM2 | Both | 0.05756 (0.0102) | 1.68E-08 | 0.02517 (0.01152) | 0.02889 | FALSE | TRUE |
| 12 | rs28372579 | 123881176 | SETD8 | Both | 0.05766 (0.01013) | 1.29E-08 | 0.03402 (0.01143) | 0.002912 | TRUE | TRUE |
| 13 | rs202346 | 51087443 | DLEU1 | Both | -0.0572 (0.009321) | 8.54E-10 | -0.03781 (0.01062) | 0.0003702 | FALSE | TRUE |
| 15 | rs16869 | 74322431 | PML | Female only | -0.05507 (0.009661) | 1.21E-08 | -0.02661 (0.01081) | 0.01381 | FALSE | FALSE |
| 15 | rs2585058 | 84284552 | SH3GL3 | Both | 0.04477 (0.008172) | 4.31E-08 | 0.03885 (0.009281) | 0.00002845 | FALSE | FALSE |
| 15 | rs2562781 | 84312568 | ADAMTSL3 | Both | -0.05845 (0.009934) | 4.04E-09 | -0.0366 (0.01128) | 0.001178 | FALSE | TRUE |
| 15 | rs4843154 | 84419524 | ADAMTSL3 | Female only | 0.04468 (0.008186) | 4.87E-08 | 0.02123 (0.009253) | 0.02176 | FALSE | FALSE |
| 15 | rs5015595 | 84493651 | ADAMTSL3 | Both | 0.04053 (0.008204) | 7.85E-07 | 0.06097 (0.009267) | 4.857E-11 | FALSE | TRUE |
| 15 | rs4594227 | 84497207 | ADAMTSL3 | Both | 0.03295 (0.00822) | 6.11E-05 | 0.05539 (0.009282) | 2.452E-09 | FALSE | FALSE |
| 15 | rs28801104 | 84552843 | ADAMTSL3 | Both | -0.06108 (0.008155) | 7.12E-14 | -0.06315 (0.009201) | 6.891E-12 | FALSE | TRUE |
| 15 | rs4414460 | 84562740 | ADAMTSL3 | Female only | 0.05056 (0.008739) | 7.29E-09 | 0.02388 (0.009905) | 0.0159 | FALSE | FALSE |
| 15 | rs11259926 | 84566248 | ADAMTSL3 | Both | -0.03881 (0.008863) | 1.2E-05 | -0.06064 (0.01004) | 1.537E-09 | FALSE | TRUE |
| 15 | rs11637264 | 84580024 | ADAMTSL3 | Both | 0.02708 (0.01045) | 0.009558 | 0.06592 (0.01171) | 1.816E-08 | FALSE | FALSE |
| 15 | rs2280470 | 89395626 | ACAN | Both | 0.04982 (0.008663) | 8.95E-09 | 0.03392 (0.009789) | 0.000531 | TRUE | TRUE |
| 17 | rs3764419 | 29164023 | ATAD5 | Both | -0.06026 (0.008323) | 4.58E-13 | -0.05007 (0.009466) | 0.000000124 | TRUE | TRUE |
| 17 | rs2079795 | 59496649 | C17orf82 | Both | 0.04121 (0.00862) | 1.75E-06 | 0.0594 (0.009742) | 1.097E-09 | TRUE | TRUE |
| 17 | rs3020619 | 61993137 | CSHL1 | Both | 0.05118 (0.009141) | 2.18E-08 | 0.04911 (0.01036) | 0.000002156 | TRUE | TRUE |
| 18 | rs9947743 | 20708321 | CABLES1 | Both | -0.04752 (0.01006) | 2.3E-06 | -0.0759 (0.01144) | 3.31E-11 | FALSE | TRUE |
| 18 | rs11082304 | 20720973 | CABLES1 | Both | -0.03483 (0.008122) | 1.81E-05 | -0.05287 (0.009275) | 1.217E-08 | TRUE | TRUE |
| 19 | rs4542783 | 8642160 | MYO1F | Both | -0.04631 (0.008141) | 1.3E-08 | -0.02741 (0.009322) | 0.00328 | FALSE | FALSE |
| 19 | rs11670030 | 8669867 | ADAMTS10 | Both | -0.09169 (0.01605) | 1.13E-08 | -0.1156 (0.01829) | 2.656E-10 | FALSE | FALSE |
| 20 | rs1884902 | 6608502 | CASC20 | Both | 0.05031 (0.00843) | 2.44E-09 | 0.03544 (0.009538) | 0.0002028 | FALSE | TRUE |
| 20 | rs6038566 | 6615197 | RP5-859D4.3 | Both | -0.04705 (0.008243) | 1.15E-08 | -0.03105 (0.009362) | 0.0009127 | FALSE | TRUE |
| 20 | rs291700 | 31981849 | CDK5RAP1 | Both | -0.02887 (0.008826) | 0.001073 | -0.05677 (0.009986) | 1.323E-08 | FALSE | TRUE |
| 20 | rs55990870 | 32302908 | PXMP4 | Both | -0.0367 (0.00943) | 9.99E-05 | -0.05946 (0.01056) | 1.832E-08 | FALSE | TRUE |
| 20 | rs7280 | 33864484 | EDEM2 | Both | 0.04442 (0.00825) | 7.35E-08 | 0.05901 (0.009355) | 2.882E-10 | FALSE | FALSE |
| 20 | rs143384 | 34025756 | GDF5 | Both | 0.0686 (0.008303) | 1.49E-16 | 0.08004 (0.009415) | 1.995E-17 | TRUE | TRUE |

| | | | | | | | | | | |
|---|---|---|---|---|---|---|---|---|---|---|
| 20 | rs224371 | 34074831 | CEP250 | Both | 0.06323 (0.009597) | 4.52E-11 | 0.0572 (0.01083) | 1.298E-07 | FALSE | TRUE |
| 20 | rs6060529 | 34227500 | CPNE1 | Both | 0.05901 (0.01058) | 2.5E-08 | 0.05923 (0.01206) | 9.139E-07 | FALSE | TRUE |
| 20 | rs6058376 | 34543834 | SCAND1 | Both | 0.06294 (0.01078) | 5.38E-09 | 0.05133 (0.01232) | 0.00003104 | FALSE | TRUE |
| 23 | X:78653645:A:G | 78653645 | RP1-164L12.1/CORO1CP1 | Both | -0.05126 (0.008419) | 1.16E-09 | -0.06067 (0.01349) | 0.000006931 | FALSE | FALSE |
